# Generational gains in memory capacity and stability may account for declining dementia incidence rates in Europe and the United States

**DOI:** 10.64898/2026.04.14.26350835

**Authors:** Anders M. Fjell, Edvard O.S. Grødem, Gabriela Lunansky, Didac Vidal-Piñeiro, Ole Rogeberg, the Alzheimer’s Disease Neuroimaging Initiative, the Vietnam Era Twin Study of Aging, Mayo Clinic Study of Aging, the Australian Imaging Biomarkers and Lifestyle flagship study of ageing, The Health and Aging Brain Study (HABS-HD) Study Team, Kristine B. Walhovd

**Affiliations:** Center for Lifespan Changes in Brain and Cognition, University of Oslo, Norway; Computational Radiology and Artificial Intelligence, Department of Radiology and Nuclear Medicine, Oslo University Hospital, Norway; Department of Epidemiology and Data Science, Amsterdam University Medical Centers (VUmc); Ragnar Frisch Centre for Economic Research, Oslo, Norway

## Abstract

Dementia incidence has been declining in Western societies for decades, but whether this reflects higher cognitive capacity entering old age, slower cognitive decline, or both remains unresolved. Analysing ∼783,000 episodic memory assessments from ∼219,000 individuals across five longitudinal cohorts, we find that later-born cohorts benefit from a double dividend: higher memory levels entering old age and slower rates of decline. The projected 20-year cohort advantage at age 80 is of sufficient magnitude to plausibly account for the observed 13% per-decade decline in dementia incidence reported in meta-analyses. Generational gains are disproportionately concentrated among the fastest-declining individuals, and are reflected in lower hippocampal atrophy rates in an independent sample. A formal bounding analysis shows that the double dividend is robust across a range of plausible period assumptions, consistent with environmental conditions operating across the lifespan having reshaped the architecture of human cognitive aging.

## Introduction

Western societies have seen persistent declines in dementia incidence for decades [1-4], offering hope that the projected global burden due to the aging of the population may be modifiable [5, 6]. The Lancet Commission estimates that nearly half of dementia cases could be prevented or delayed through environmental and lifestyle interventions [7]. Yet a fundamental question remains unresolved: do these declining incidence rates reflect older adults today having higher cognitive capacity developed early in life [8, 9], slower rates of cognitive decline in aging, or both? The answer matters because these two mechanisms likely have partially distinct origins. Cognitive performance in adulthood correlates highly with childhood function [10], and individual differences in level of function predominantly reflect early-life conditions that build cognitive capacity. In contrast, slope differences - differences in rate of change in aging - may reflect a broader mixture of influences [11], including additional life-course exposures and generational changes in how the aging brain sustains cognitive function.

Prior studies have produced conflicting answers [12, 13]. Some find that today’s older adults perform better than previous generations but follow similar decline trajectories [14], while others suggest the rate of decline has also improved [12, 15-17]. These inconsistencies may partly reflect how cognitive aging is conceptualized. The prevailing view treats it as smooth and monotonic, with individuals differing mainly in starting level in young adulthood, age of onset, and rate of decline [18-20]. Recent large-scale evidence challenges this picture: at the individual level, memory aging is often punctuated, characterized by extended plateaus of stability interrupted by abrupt transitions to lower function, and the smooth decline visible in population averages can largely be a statistical artefact of aggregation[21]. Cohort comparisons that treat all decline as equivalent may therefore miss important generational differences in how and when cognitive loss occurs.

To address this, we assembled a large-scale dataset of ∼783,000 memory assessments from ∼219,000 individuals across five premier longitudinal cohorts spanning Europe and the United States, followed for up to 30 years. We target verbal episodic memory, the primary clinical indicator of Alzheimer’s-related cognitive decline [22], included in all major dementia screening batteries [23-25] and critical for daily function [26], allowing direct calibration against established clinical thresholds for mild cognitive impairment and dementia. In addition to asking whether average performance differs between generations, we characterise the full distribution of individual trajectories, translate the observed advantages into MCI and dementia thresholds, and validate the clinical relevance of the trajectory measures at two levels: clinically, by linking individual decline rates to independently assessed cognitive impairment and dementia status, and neurobiologically, by showing that the same trajectory categories are associated with differential hippocampal atrophy rates in an independent sample, indicating that the memory slope differences we study reflect genuine variation in the pace of neurodegeneration in a memory-critical brain structure.

We show that later-born cohorts benefit from a double dividend of higher memory capacity and slower cognitive decline - differences that are associated with both lower clinical impairment rates and slower hippocampal atrophy at the individual level - and that the combined projected generational advantage is of sufficient magnitude to plausibly account for the observed decline in dementia incidence in Western societies [3]. The question of whether these differences reflect genuine generational change or calendar-year period effects is addressed through a bounding analysis that characterises the full range of cohort effects compatible with the data under transparent assumptions, showing that a genuine generational contribution is the most plausible interpretation across a range of empirically defensible period specifications. The core findings are further replicated under an independent Bayesian hierarchical framework, confirming robustness to analytical choices.

## Results

Sample characteristics are presented in Table 1 and data harmonization in SI Section 1. For each participant, two person-level summary statistics were derived from their longitudinal practice-corrected recall record: a memory level estimate and a memory change estimate. Both were computed using Theil–Sen regression - the median of all pairwise slopes across a participant’s observations - which provides resistance to outlying observations and irregular assessment spacing without distributional assumptions (SI Sections 2–4). Uncertainty in each estimate was quantified using leave-one-out jackknife resampling, enabling precision-weighted downstream analyses. These single person-level estimates of level and slope are the units of analysis in the subsequent analyses. Because episodic memory scores show substantial within-person variability across occasions, obtaining reliable estimation of individual trajectories is challenging. To ensure precise person-level estimates, analyses were restricted to participants with at least four observations. This threshold was motivated empirically: precision improved sharply from three to four observations per participant, as median jackknife standard error dropped from 1.05 to 0.54 words/year with slope direction becoming fully stable at four observations in most participants (SI Section 4 for a full validation).

**Table 1.** Sample descriptives. ELSA - English Longitudinal Study of Ageing; HRS - Health and Retirement Study (US); SHARE - Survey of Health, Ageing and Retirement in Europe; TILDA - The Irish Longitudinal Study on Ageing; LASA - Longitudinal Aging Study Amsterdam.

| Cohort | Sample | n | Obs | Age | Waves | Interval |
| --- | --- | --- | --- | --- | --- | --- |
| ELSA | Full | 20,886 | 93,727 | 67.2 (50–102) | 4.5 (1–11) | 7.9 (0–22) |
|  | ≥4 obs | 10,231 | 75,647 | 68.0 (50–102) | 7.4 (4–11) | 14.2 (5–22) |
| HRS | Full | 35,420 | 210,172 | 67.9 (50–107) | 5.9 (1–13) | 10.4 (0–26) |
|  | ≥4 obs | 22,905 | 183,716 | 68.5 (50–107) | 8.0 (4–13) | 14.7 (5–26) |
| SHARE | Full | 150,358 | 428,896 | 67.9 (50–104) | 2.9 (1–8) | 5.1 (0–18) |
|  | ≥4 obs | 47,954 | 249,373 | 68.5 (50–104) | 5.2 (4–8) | 11.1 (6–18) |
| TILDA | Full | 7,850 | 33,654 | 66.4 (50–84) | 4.3 (1–6) | 7.9 (0–12) |
|  | ≥4 obs | 5,196 | 28,626 | 66.2 (50–84) | 5.5 (4–6) | 10.6 (5–12) |
| LASA | Full | 4,641 | 16,718 | 71.7 (54–101) | 3.6 (1–10) | 9.1 (0–29) |
|  | ≥4 obs | 2,258 | 12,387 | 71.8 (54–101) | 5.5 (4–10) | 15.4 (9–29) |
| Total | Full | 219,155 | 783,167 | 67.8 (50–107) | 3.6 (1–13) | 6.4 (0–29) |
|  | ≥4 obs | 88,544 | 549,749 | 68.4 (50–107) | 6.2 (4–13) | 12.5 (5–29) |

Adjusting for age and study, memory level and slope were plotted over age for consecutive 5-year birth cohorts from the 1910-14 to the 1960-64 cohort (Figure 1), revealing a consistent generational advantage: on average, one decade of later birth was associated with 0.54 additional words at memory level (95% CI: 0.49–0.59, p < 0.001) and 0.070 words/year shallower decline rate (95% CI: 0.063–0.077, p < 0.001). These initial estimates do not separate possible influence from period drift and therefore represent an upper bound on the cohort-specific effects (see below for analyses addressing this). The cohort ordering in both level and slope was preserved in sex-stratified analyses (SI Section 5), confirming that pooling across sex does not obscure divergent patterns. A full sex × cohort interaction analysis, requiring country-stratified modelling across the 30+ countries represented, remains a direction for future work [27-29]. All subsequent analyses therefore pool across sex. The cohort ordering was preserved when relaxing the ≥4-wave requirement to ≥2 or ≥3 observations (SI Section 8). Inverse-probability reweighting of the ≥4-wave sample back to the baseline-eligible population yielded nearly identical cohort differences (SI Section 8), indicating that selection into longer follow-up on the basis of observed baseline characteristics does not account for the cohort findings. However, selective attrition driven by cognitive decline itself, whereby faster decliners are less likely to complete four waves, cannot be fully addressed by reweighting and represents a residual limitation. The preserved cohort ordering when including participants with fewer waves still provides partial reassurance on this point. We also replicated the findings under a Bayesian hierarchical framework, including all datapoints and with inference performed jointly over memory level, rate of change, and retest effects (see details below).

**Figure 1.**
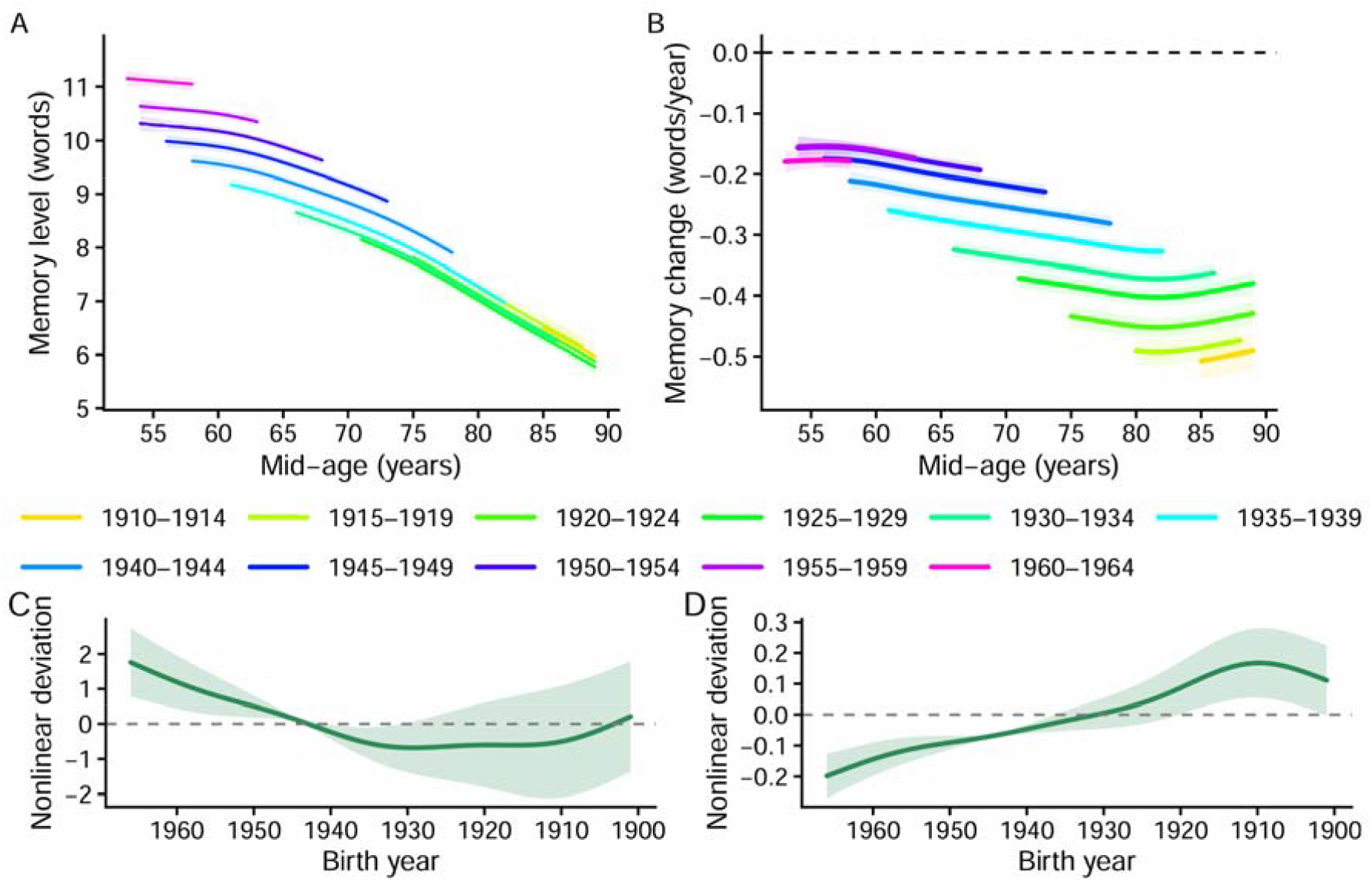
Episodic memory level and change by birth cohort across overlapping age ranges. Top row: memory level (A) and rate of change (Theil–Sen slope, B). Curves show GAM predictions with a shared age smooth and birth-cohort intercept offsets, adjusted for study. Shaded bands indicate 95% CI. Curves are shown only where the cohort × age-bin cell contains ≥50 participants; predictions are referenced to ELSA at baseline. Bottom row: nonlinear cohort deviations for memory level (C) and slope (D), estimated from penalised regression splines fitted to residuals after removing linear age and cohort trends, without assumptions about age–period–cohort separation (SI Section 18; all components p < 2×10^−1^□). Values are centred to zero mean; positive indicates that the cohort performs better than the linear cohort trend predicts, negative indicates worse. The birth cohort scale is reversed to match the age scale in the top row.

The cohort trajectories also suggest that each generational gain is not uniform. The spacing between adjacent level curves is wider for more recently born cohorts, while the slope advantage appears largest for the oldest cohorts. Formal analysis of the identifiable nonlinear cohort components confirms both patterns (Figure 1 bottom row; SI Section 19). These components capture curvature in the cohort trend - deviations from a simple linear generational improvement - that can be estimated from the data without requiring assumptions about the separation of age, period, and cohort effects. Generational gains in memory level have been growing larger with successive cohorts rather than accumulating at a steady rate, while the slope advantage is front-loaded toward older cohorts and diminishes for those born after approximately 1950.

### Slope distribution structure across cohorts and age

Having established that later-born cohorts show both higher memory levels and shallower decline rates, we next ask whether these generational differences reflect a uniform shift of the slope distribution or a disproportionate reduction in the most severe trajectories (Figure 2). We estimated cohort effects across the full slope distribution using quantile regression, adjusting for age and study. The cohort improvement was clearly not uniform: the effect at the 10th percentile, representing the most rapidly declining individuals, was approximately twice the size of the median effect (0.104 vs. 0.053 words/year per decade of birth, both p < 0.001, Fig 2, right panel; SI Section 10 for details). Generational progress has therefore disproportionately reduced the prevalence of the worst decline trajectories, precisely those most likely to cross clinical thresholds, while also improving typical trajectories throughout the distribution. The left-tail concentration was not explained by proximity-to-death artefacts. When repeating the analyses with participants who died within 2 years of their last observation in SHARE and HRS (88% of the analytic sample) excluded, the left-tail concentration was preserved. The cohort effect at the 10th percentile remained approximately 1.5 times the median effect (restricted sample 10th/50th ratio = 1.52 versus 1.71 in the full sample), with fully overlapping confidence intervals at every quantile, and cohort differences were also evident at ages 55–70 where terminal decline effects are negligible (SI Section 13).

**Figure 2.**
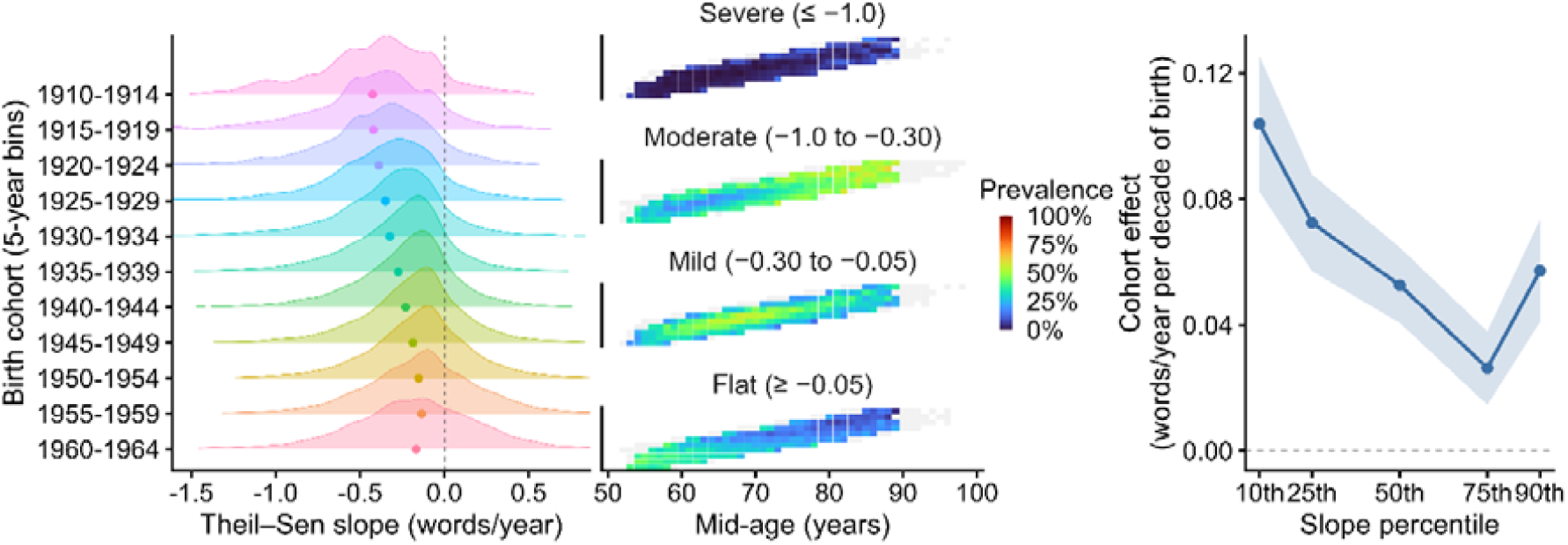
Distributional structure of episodic memory change slopes across birth cohort and age. Left panel: kernel density ridgelines of person-level Theil–Sen slopes (words/year) by 5-year birth cohort (n ≥ 200 per cohort shown); dots indicate cohort mean slopes; dashed vertical line marks zero. Middle panel: slopes discretized into four categories; heatmaps show within-cohort prevalence across mid-age (1-year bins; cells masked when n < 50), warmer colors indicate higher prevalence, illustrating that severe decline becomes increasingly common with advancing age regardless of birth cohort. Right panel: quantile regression cohort coefficients (words/year per decade of birth) across slope percentiles, adjusted for age and study (bootstrap 95% CI, R = 500).

These results motivate a new set of analyses. The generational improvement is concentrated in the left tail, populated by those with steepest decline, yet severe decline is predominantly age-driven within each cohort (Figure 2 heatmap). Understanding the generational advantage therefore requires examining the temporal structure of individual trajectories in addition to their average rate.

### From distributional structure to trajectory architecture: bouts of stability and decline

Recent work has shown that individual memory trajectories in aging are often punctuated rather than smoothly declining, characterised by extended plateaus of relative stability interrupted by episodes of accelerated loss [21]. We therefore asked whether the cohort differences documented above are expressed as a difference in the prevalence of stable vs. steep-decline trajectory states in addition to the distributional shift (SI Section 11). Consistent with prior work, we found that stable bouts became progressively less common with advancing age across every birth cohort [21]. At every age, however, later-born cohorts showed a systematically higher probability of being in a stable period (slope ≥ −0.05 words/year), with each decade of later birth associated with 64% higher odds of stable bout membership at any given age (OR = 1.64, 95% CI: 1.59–1.68, p < 0.001, Figure 3, left panel). This cohort ordering was robust across six operationalisations of varying bout length and stability threshold (Figure S10 & S11), supporting a genuine generational difference in the probability of sustained cognitive stability.

**Figure 3.**
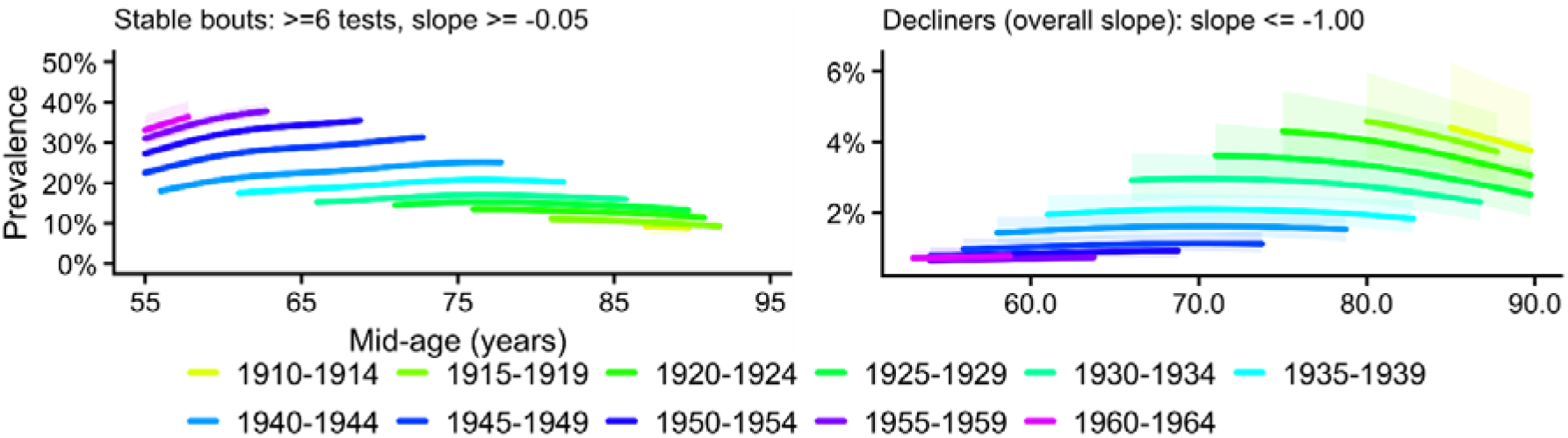
Prevalence of sustained stability and steep decline across age and birth cohort. Left panel: proportion of participants exhibiting at least one qualifying stable bout (≥6 consecutive assessments with slope ≥ −0.05 words/year), estimated from a binomial GAM with a shared smooth of age and cohort intercepts. Right panel: proportion classified as steep decliners based on overall person-level Theil–Sen slope ≤ −1.0 words/year, consistent with the Severe slope category used throughout. Curves shown where cohort × age-bin n ≥ 50. Cohorts are compared at equivalent mid-ages, so differences between curves reflect cohort rather than age.

The mirror image was seen for steep decline, defined as ≤ -1 word/year: earlier-born cohorts were consistently more likely to meet steep-decline criteria at the same age, with each decade of later birth associated with 38% lower odds (OR = 0.62, 95% CI: 0.55–0.70, p < 0.001, Figure 3, right panel). This was evident both when varying the required duration of a decline period or using the total follow-up interval for each participant, showing convergence across two different operationalisations. The cohort separation was most pronounced at older ages, where earlier-born cohorts reached steep-decline prevalence of ≈5% that later-born cohorts had not yet approached. Together, these analyses converge on a consistent account of how the generational improvement is expressed. The distributional analyses showed that the cohort improvement is largest in the left tail, while the bout analyses reveal a higher prevalence of stable plateaus and a lower prevalence of decline episodes that, when aggregated across people, produce the smoother population curves observed in later-born cohorts. Since this generational shift in trajectory architecture is most pronounced among the individuals otherwise most likely to cross thresholds for cognitive impairment and dementia, it raises the question of whether the generational advantage is large enough to account for the observed decline in dementia incidence.

### Memory dividend and declining Alzheimer’s incidence

To assess whether the magnitude of the generational advantage is clinically meaningful, we decomposed the total estimated advantage into level and slope components and anchored the result to established memory benchmarks for mild cognitive impairment and dementia. The level component represents the cohort difference in memory already present at age 60, derived from the estimated linear cohort drift in memory level from the preceding analyses. The slope component represents the additional advantage that accumulates through slower decline between ages 60 and 80, derived from the corresponding drift in rate of change. Figure 4A illustrates this as an empirically grounded projection: under the observed cohort drifts, two cohorts born 20 years apart would be expected to differ by approximately 2.5 words at age 60, widening to approximately 5.0 words at age 80 as the slower decline of the later-born cohort accumulates over time. These are model-derived estimates rather than directly observed differences, which means that they characterise the expected generational gap under current trends, assuming the estimated cohort drifts hold linearly across cohorts and ages. Because level is estimated at mid-observation age and slope from the full trajectory, the two components are not fully independent, and their simple sum should be understood as a projection of the expected total advantage rather than a precise decomposition into separable contributions. With that in mind, level accounts for approximately 47% and slope for 53% of the estimated ∼5.0-word 20-year cohort advantage at age 80.

**Figure 4.**
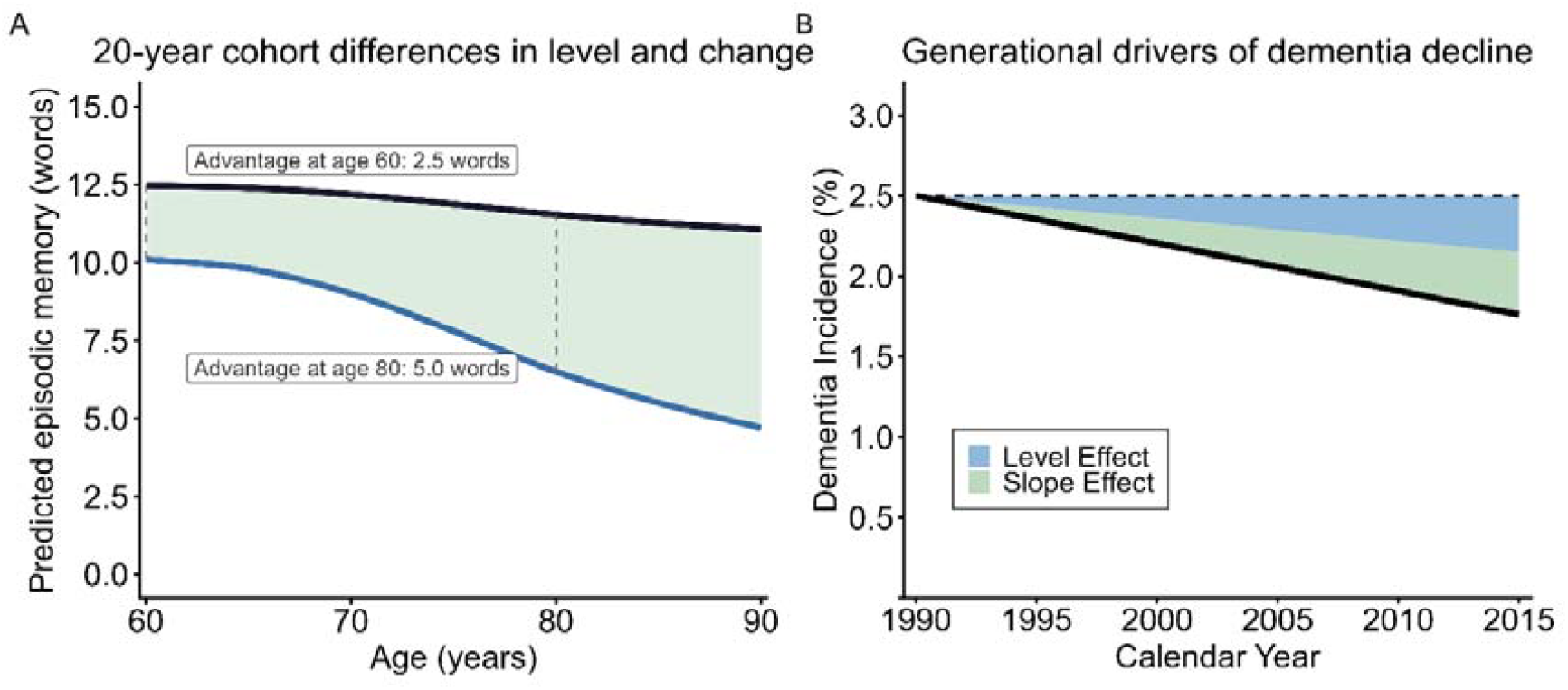
Decomposing cohort differences into level and change components. Panel A. Predicted episodic memory trajectories from ages 60 to 90 for two cohorts born 20 years apart. The earlier-born trajectory (blue) is the population-level prediction from a growth-curve model (SI Section 6); the reference study is ELSA at baseline. The later-born trajectory (dark) adds the estimated cohort offsets: a constant upward shift at age 60 equal to the level advantage, plus proportionally slower decline over age. Panel B. Generational memory dividend mapped against the empirical decline in dementia incidence (1990–2015; [3]). The solid line shows observed age-standardized incidence for the 75–79 age group; the dashed line shows the counterfactual if memory capacity and aging architecture had remained at 1990 levels. Shaded regions decompose the full observed decline into level (blue) and slope (green) contributions, proportional to their respective shares of the total cohort advantage at age 80.

To assess the clinical magnitude of this advantage, we anchored our memory measure to benchmarks from the Alzheimer’s Disease Neuroimaging Initiative (ADNI), scaled to our 0– 20 metric (SI Section 9). Mean scores were 14.2 words (cognitively normal, CN), 10.8 (mild cognitive impairment, MCI), and 7.8 (Alzheimer’s disease, AD), with excellent diagnostic discrimination (AUC = 0.959 for CN vs. AD; AUC = 0.821 for CN vs. MCI). The 20-year cohort advantage of 5.0 words represents 78% of the CN–AD gap and exceeds the gap between CN and the pooled MCI+AD group. These gains represent a generational improvement of sufficient magnitude to plausibly account for the full observed decline in dementia incidence. The documented 13% per-decade decline in age-standardized incidence (1990–2015) corresponds to roughly 7 fewer dementia diagnoses per 1,000 people per year among 75–79 year-olds [3]. This means that a person born in 1940 who scored at the MCI boundary in their late 70s would, if born instead in 1960, be expected to score in the cognitively normal range.

Although we cannot calculate exact explained variance without additional assumptions about the memory–dementia relationship across the full population distribution, the generational advantage matches the clinical gap at which dementia ascertainment begins and is sufficient in magnitude to account for the observed incidence decline. The slope component contributes approximately half of this advantage, and the quantile regression and bout analyses show this improvement is disproportionately concentrated among those declining most rapidly. Together, these findings suggest that the generational shift may reduce the prevalence of the trajectories that would otherwise carry individuals from normal cognition into the at-risk range (Figure 4B).

### Clinical and neurobiological validation

To validate the clinical relevance of our memory slope measure within the primary data, we linked person-level Theil–Sen slopes to the Langa–Weir cognitive classification in HRS [30, 31]. Langa–Weir assigns respondents to normal cognition, cognitive impairment no dementia (CIND), or dementia based on a 27-point battery encompassing orientation, arithmetic, and word-list recall, a classification that is largely independent of the recall-based slope estimates, which capture rate of change rather than level at a single occasion (SI Section 18). Slopes were estimated across each participant’s full observation window, and Langa– Weir status was assessed at the final wave of that window. The correspondence between memory slopes and clinical status was monotone across four decline categories (Figure 5, left panel). Impairment rates rose from 3.0% among flat decliners to 34.5% among severe decliners. After adjusting for mid-age of the observation window, each additional word/year of steeper decline was independently associated with an 86% reduction in the odds of remaining cognitively normal (OR = 0.14, 95% CI: 0.12–0.17, p < 0.001).

**Figure 5.**
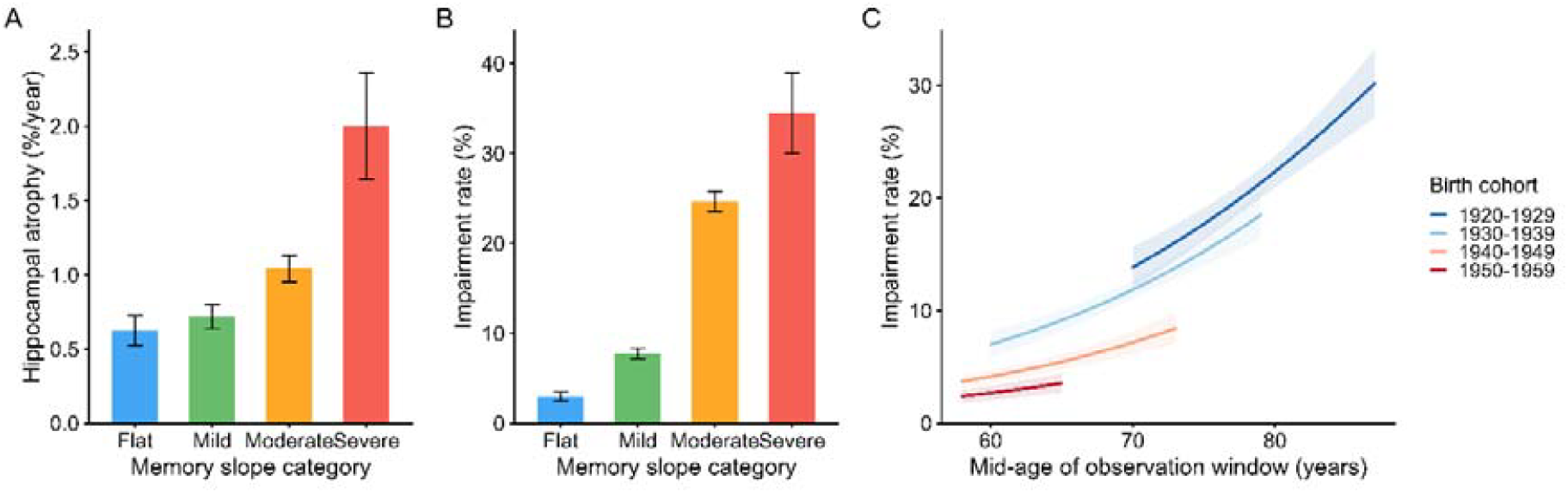
Cognitive impairment by memory slope category in HRS. A: Bilateral hippocampal volume change rate (%/year) by memory slope category (n = 1,925; median follow-up 4.1 years). Model-adjusted means (age, sex, intracranial volume, interval-weighted). B: Langa-Weir cognitive impairment at last observed wave by Theil-Sen memory slope category in HRS (n = 17,870). Categories: Flat (≥ −0.05 words/year, n=4,066, 3.0%); Mild (−0.05 to −0.30, n=7,842, 7.8%); Moderate (−0.30 to −1.0, n=5,530, 24.7%); Severe (≤ −1.0, n=432, 34.5%). Slope estimated from word-list recall. C: Langa-Weir impairment rate by birth cohort and age. Curves from binomial GAM with shared age smooth and cohort intercepts; shown where cohort x 2-year age bin n >= 30. Error bars/ shaded bands: 95% CI.

The cohort differences in clinical impairment mirrored the memory trajectory findings (Figure 5, right panel). At any given age, later-born cohorts showed substantially lower Langa–Weir impairment rates than earlier-born cohorts. The cohort separation was most pronounced at older ages, where impairment rates diverge steeply. For example, the 1940–1949 cohort reached age 70 with an impairment rate of 7.2%, comparable to the 7.0% rate observed in the 1930–1939 cohort at age 60, a roughly decade-long postponement of clinical risk. Quantifying this cohort advantage at matched ages, a logistic regression model showed that each decade of later birth was associated with 40% lower odds of impairment after adjusting for age (OR = 0.59, 95% CI: 0.52–0.68, p < 0.001), an estimate that reflects cohort differences within the observed range and should not be interpreted as a continuing secular projection. These cohort differences most likely reflect a postponement of cognitive impairment rather than its prevention. The generational advantage shifts the decline trajectory to higher ages, reducing risk at any specific age, but likely does not eliminate the eventual accumulation of cognitive loss.

Critically, this cohort protection was entirely mediated by the cohort difference in memory slopes. After adjusting for memory slope, the cohort effect on impairment was effectively eliminated (slope-adjusted OR = 1.04, 95% CI: 0.91–1.20, p = 0.55). Unlike the ADNI-based decomposition, in which level and slope contribute comparably to the total generational advantage, level enters directly into the Langa-Weir classification itself, precluding an equivalent decomposition. Moreover, the two calibrations capture fundamentally different constructs: ADNI benchmarks derive from clinically diagnosed patients, while Langa-Weir infers impairment in a population-representative sample. Hence, the ADNI comparison establishes that the combined advantage is large enough to explain the decline in dementia incidence, while the Langa-Weir mediation shows that the slope component is a proximal driver of clinical threshold crossing at any given age.

Finally, to anchor the memory slope categories in neurobiology, we examined bilateral hippocampal volume change rates in an independent sample of 1,925 participants with longitudinal MRI (median follow-up 4.1 years) and at least 4 timepoints of memory tests, adjusting for age, sex, and intracranial volume. Hippocampal atrophy rate followed a monotone gradient across slope categories: flat decliners showed −0.63%/year (95% CI: −0.73 to −0.53), moderate decliners −1.04% (−1.13 to −0.95), and severe decliners −2.00% (−2.36 to −1.64), approximately three times faster than flat decliners (Figure 5). These findings suggests that the memory slope categories used throughout this paper could reflect genuine differences in the rate of neurodegeneration in memory-critical brain structures.

### Replication under an independent Bayesian hierarchical framework

To assess robustness to model specification, we replicated the core findings using a Bayesian hierarchical model fitted jointly to the three largest cohorts (SHARE, HRS, and ELSA, with identical recall measures). The model represents a substantially different analytical approach from the main analysis in several respects. Inference was performed jointly over memory level, rate of change, and retest effects, allowing uncertainty in each component to propagate into the others. The rate of change was defined as the derivative of the level trajectory, ensuring internal consistency. Cohort was modelled as a continuous smooth function rather than categorical 5-year bins, and participants with fewer than four sessions were included, reducing the potential healthy-participant bias further. Age and cohort effects were estimated using flexible cubic splines and observed memory scores were modelled with a Student-t likelihood to provide robustness to outliers. Full model specification and posterior inference details are provided in Figure 6 and SI Section 20.

**Figure 6.**
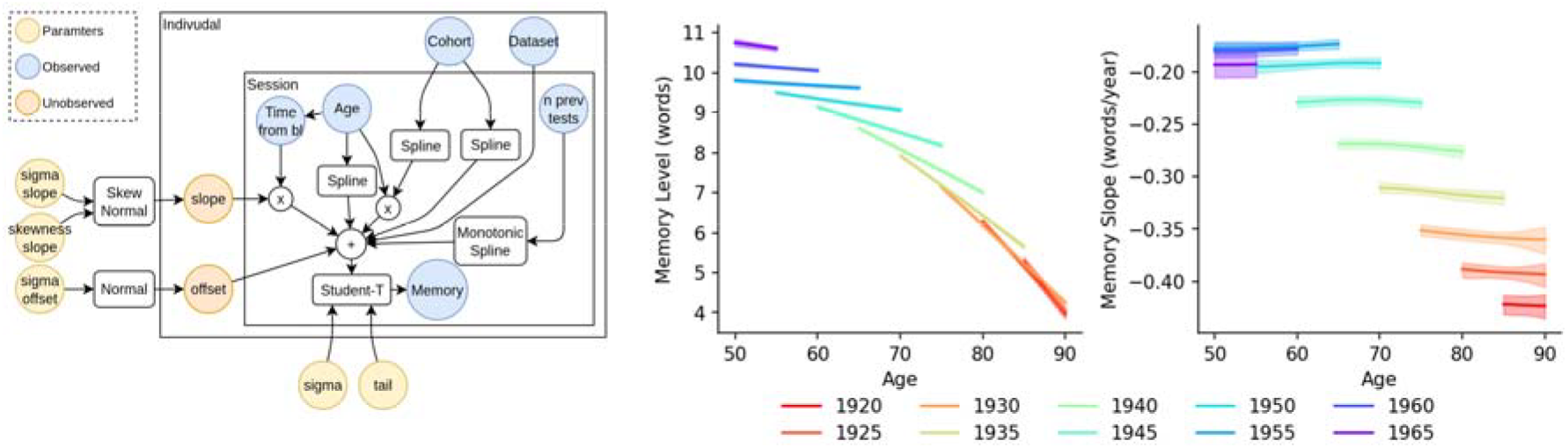
Bayesian hierarchical modelling of memory level and change. Left panel: schematic of the Bayesian hierarchical model. Transformations and distributions are shown as rounded boxes; observations, latent states, and parameters as circles. Spline parameters are omitted for simplicity. “Time from bl” indicates time in years from the middle session; “n prev tests” indicates the number of previous recall tests. Middle and right panels: episodic memory level and rate of change by birth cohort. Curves show regression splines from the hierarchical Bayesian model, adjusted for study, with 95% credible interval bands. Curves are shown only for ages and cohorts within the period 2000–2025. Analysis based on SHARE, ELSA, and HRS.

Results were consistent with the main analysis, with both the level and slope cohort ordering preserved, and the nonlinear cohort pattern closely replicated. Accelerating generational gains in memory level for more recently born cohorts were detected, as was a front-loaded slope advantage largest for cohorts born approximately 1930–1955 which diminished for more recently born cohorts. This qualitative replication across two independent analytical frameworks demonstrates robustness of the core findings.

### Disentangling cohort from period effects

The preceding analyses establish that later-born cohorts show higher memory levels, shallower decline rates, and lower clinical impairment at matched ages. The question of attribution, whether these differences reflect genuine generational change or also calendar-year period trends, matters for mechanistic interpretation and for projecting future trends. We address it here through a bounding analysis [32, 33]. The core challenge is that calendar year equals age plus birth year exactly, making the linear contributions of age, period, and cohort mathematically inseparable from observational data alone. Any combination of age, period, and cohort trends that sums correctly is equally consistent with the data. This is the APC identification problem.

To narrow the admissible range of solutions, we make two non-controversial assumptions. Memory should not improve with age in the 50–90 year window studied [34-36], and later-born cohorts should not be systematically disadvantaged relative to earlier-born cohorts across the 1910–1964 birth range examined, consistent with the well-documented secular gains in cognitive performance across this period [14, 37, 38]. These constraints define the set of combinations that are both mathematically consistent with the data and substantively plausible, shown as shaded regions in Figure 6. Within this admissible range, which by construction excludes negative cohort effects, the question is how far the data place the cohort effect from zero. The tipping point, the period drift at which the cohort effect would reach exactly zero, lies at C = 1.0 word per decade of calendar-year improvement, roughly equal to the average annual decline rate in the full sample. Crucially, this tipping point falls within the admissible range and is not ruled out by the sign constraints alone, meaning that a zero cohort effect on level, while requiring an implausibly large period trend, cannot be excluded on logical grounds alone. The full canonical solution lines are provided in SI Section 12.

As a further sensitivity check, we fitted flexible period splines of varying smoothness alongside the linear age and cohort terms, asking whether any empirically plausible period specification could substantially reduce or eliminate the cohort effect. This approach does not identify the true period drift, as nonlinear period and cohort components cannot be cleanly separated without additional assumptions, but it tests whether the cohort finding is sensitive to the assumed shape of the period trend. For levels, all specifications satisfying the sign constraints implied period drifts at or below zero, meaning no plausible smooth period trend worked in the direction needed to explain away the cohort effect. For slopes, valid specifications implied period drifts slightly above and below zero, meaning the direction of any calendar-year trend on decline rates is uncertain, but none approached the tipping point at which the cohort finding would be eliminated. The cohort findings are therefore robust not only across the linear grid but also to varying assumptions about the shape of the period trend, within the constraints imposed by the sign restrictions (SI Section 6). The identifiable nonlinear APC components, estimated without these limitations, are reported in SI Section 18 and are consistent with the linear bounding results.

Figure 7 displays the results of this bounding analysis from the Bayesian framework. The shaded regions show the range of age, period, and cohort effects that remain consistent with the data under the two sign constraints (memory does not improve with age; later-born cohorts are not disadvantaged). The wider the shaded band, the greater the residual uncertainty; a narrow band indicates the data constrain the effect tightly even within the admissible range. It is important to note that the positivity of the cohort effects visible in these panels is partly a consequence of the constraints themselves: within the admissible range, the cohort effect on level is bounded away from zero by assumption, and the data further narrow the admissible range to values that are clearly positive. For memory slope, the cohort effect is positive under most plausible specifications, but the lower boundary of the admissible range approaches zero under the most conservative assumptions, meaning a near-zero generational slope effect, though unlikely given the empirical period specifications, cannot be entirely excluded. Notably, the period effect on slope appears likely to be slightly positive under the most empirically supported specifications, suggesting that the observed flat slope trajectories across cohorts may partly reflect favourable calendar-year trends, which, if true, would mean the apparent cohort advantage in slope is a conservative estimate of the true generational improvement.

**Figure 7.**
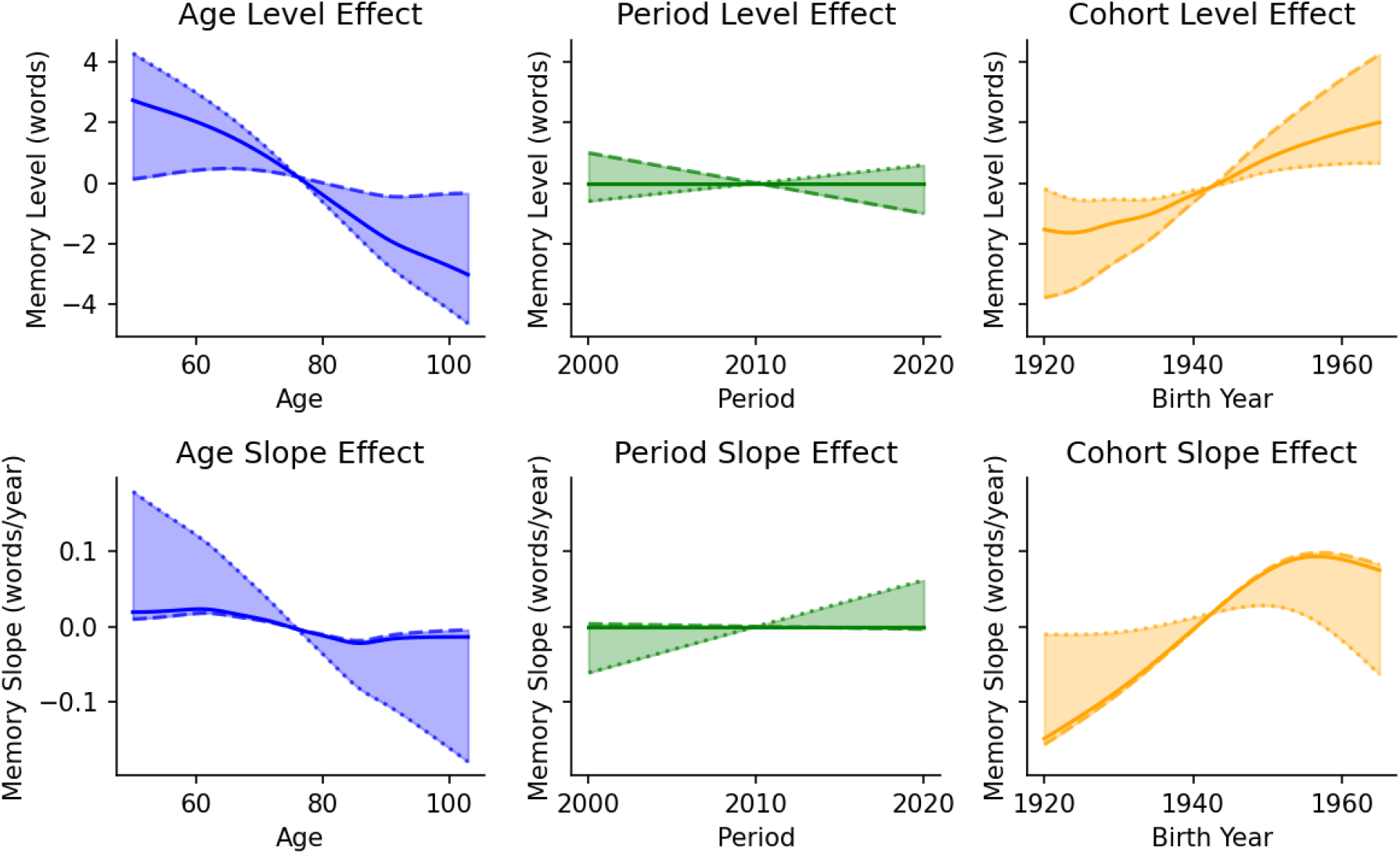
Age–period–cohort bounding analysis for memory level (top) and memory slope (bottom) from the hierarchical Bayesian model. Each panel shows the plausible range of combined linear and non-linear effects of age, birth cohort, and period consistent with the data under the sign constraints, shown as shaded regions. Solid lines show effects under the assumption of no period trend, dotted lines show effects assuming no linear cohort effect, and dashed lines show effects assuming no linear age effect.

## Discussion

We find that later-born cohorts in Europe and the United States benefit from a double dividend of higher memory capacity and slower cognitive decline that appears to have reshaped population-level cognitive aging. The combined generational advantage could be of sufficient magnitude to plausibly account for the observed 13% per decade decline in clinical dementia incidence [3] without requiring any additional mechanism. This conclusion is supported by four converging lines of evidence: the cohort protection against clinical impairment in a population sample is mediated by the cohort improvement in memory slopes, the core findings replicate under an independent Bayesian hierarchical framework using a substantially different modelling approach, a formal bounding analysis shows the level advantage is robust across all plausible period assumptions and that a genuine generational contribution to the slope advantage is the most parsimonious interpretation of the data, and the trajectory differences were associated with differential hippocampal atrophy rates in an independent MRI sample, implying they may reflect genuine variation in neurodegeneration rather than test-score artefact. Understanding what drives this advantage requires going beyond average decline rates to examine how generational gains are expressed within the punctuated architecture of individual cognitive aging, in the balance between stability and decline that population averages often obscure.

### The architecture of change: stability vs. decline

Being born 10 years later is associated with approximately 10% higher memory score at age 60, corresponding to d = 0.32 per decade, which is consistent with meta-analytic estimates from the level literature (d ≈ 0.20–0.48 per decade [13, 14]). The slope finding is where the present study most clearly advances prior work. The cohort slope advantage of approximately 0.065 words/year per decade (d = 0.18) may appear small by conventional standards, and prior studies reporting null slope effects typically had confidence intervals fully encompassing this effect size, which reflect the limits of statistical resolution in smaller samples (SI Section 14). The present sample of 88,550 participants with at least four memory tests over a median follow-up of 12.5 years provides the resolution to estimate this effect with higher precision.

Beyond the magnitude of the average advantage, we find that generational improvements in cognitive aging are disproportionately concentrated among those declining most rapidly. Quantile regression shows that the cohort effect at the 10th percentile of the slope distribution, the fastest decliners, is approximately twice the size of the median effect, indicating that later-born cohorts have selectively reduced the prevalence of the worst trajectories rather than shifting all trajectories uniformly. Larger Flynn-effects for the lower end of the score distribution have previously been reported for cognitive level [39, 40], but whether the same applies to longitudinal rates of change has, to our knowledge, not been examined. Yet the improvement is not confined to the extremes: the median and upper quantiles also show significant cohort advantages, reflecting a genuine broad redistribution rather than a narrowing of the left tail alone.

Further, our bout analyses demonstrate that later-born cohorts are also more likely to remain in sustained periods of stability and less likely to enter episodes of rapid loss. This goes beyond regarding the slope advantage as a slower pace of smooth, monotonic age-reductions [18-20] by revealing how individual advantage is instantiated through longer and more frequent plateaus of cognitive stability. Consistent with models of biological resilience [41, 42], generational progress appears to have produced a cognitive system less susceptible to the transitions that precipitate functional loss, whether through reduced neuropathological burden, greater cognitive resilience, or both. Across all cohorts, the probability of sustained stability falls sharply in the eighth and ninth decades, but generational factors appear to extend the plateaus of cognitive health rather than eliminate the vulnerability of the aging brain [43]. These patterns are consistent with a punctuated model of cognitive aging in which trajectories alternate between stable plateaus and episodes of decline [21] and suggest that generational progress has shifted the probability of residing in each state rather than simply slowing a uniform decline.

The neurobiological reality of these trajectory differences is supported by the hippocampal atrophy analyses. Participants classified as severe decliners in an independent sample showed approximately three times faster bilateral hippocampal atrophy than flat decliners (−2.00 vs. −0.63%/year), with a monotone gradient across all four categories. This finding means that the memory slope categories may not be psychometric abstractions but rather map onto the pace of neurodegeneration in the brain’s primary memory structure [43]. More broadly, it suggests that generational improvements in memory slopes could reflect genuine differences in the rate at which the hippocampal system accumulates neurobiological damage, providing a possible mechanistic bridge between the environmental drivers of cognitive aging and the clinical outcomes they ultimately determine.

### Drivers of the Double Dividend and Implications for Public Health

The speed of the generational changes we document rules out genetic drift as an explanation. Allele frequencies cannot have shifted meaningfully across this time, and within-family analyses of the Flynn effect confirm that level gains reflect environmental rather than genetic change [44]. The present findings extend this logic to the slope component: cohort differences in the rate and architecture of memory decline are too rapid and too systematic to reflect anything other than environmental change. To dismiss these findings as a period artefact would require a sustained calendar-year improvement in memory performance roughly equal to the average annual decline rate, which is a magnitude not supported by the empirical period specifications in our bounding analysis. This provides grounds for attributing both components of the double dividend causally to modifiable conditions.

The two components, however, implicate different environmental windows and therefore different policy levers. The level advantage is anchored in early life. Multiple factors such as education, nutrition, and childhood health may contribute to build the memory capacity that individuals carry into old age [8, 10, 13, 44-48]. Crucially, these same factors do not typically explain the slope advantage, for instance, education predicts memory level but not the rate of longitudinal change [45, 46, 49], suggesting that the window for intervention may differ fundamentally between the two components. Level advantages require early-life investment, while slope advantages may remain tractable across adulthood. Plausible candidates include broad environmental and cardiovascular improvements that have accumulated across these cohorts’ adult lives, such as management of vascular risk factors by antihypertensives [50], reduced smoking, and lower pollution burden [7, 11, 51-53].

This decomposition reveals a systematic gap in current dementia prevention frameworks. The Lancet Commission’s influential model [7] and the broader risk-factor literature it represents focus on identifying modifiable factors, but do not decompose dementia risk into level versus slope components, and therefore cannot distinguish whether a given factor operates by raising cognitive starting points, slowing the rate of decline, or both. Without this decomposition, the timing of intervention cannot be correctly specified [8]. A factor that primarily affects level requires early-life investment to have population-level impact [54, 55], while one that affects slope can potentially be targeted across adulthood [11, 56]. Our findings suggest that early-life conditions account for approximately half of the total generational advantage up to 80 years under reasonable assumptions about period drift, a contribution that is systematically underweighted in the Lancet framework that treats education as the sole early-life entry point and assigns the remainder of preventable risk to midlife and late-life factors. The practical consequence is a prevention agenda skewed toward later intervention windows, when a substantial portion of the generational dividend has already been determined.

These findings bear directly on the debate about whether the decline in dementia incidence [1-4] has reached its limit. Recent data suggest the Flynn effect has plateaued or reversed in several countries [44, 48], and academic disruptions following COVID-19 may further compress early-life cognitive gains in younger cohorts [57, 58]. While education has positive impacts on cognitive function lasting to old age [59], the potential for cognitive improvements from educational expansion has plateaued in many countries (Bratsberg et al., in review). Furthermore, improved medical management of risk factors such as hypertension, may not suffice to uphold health gains across cohorts into older age in a population that has entered midlife with a much heavier burden of e.g. obesity [60] and physical inactivity [61] than current older adults, with foreseeable negative consequences for brain health. These emerging threats do not bear equally on the two components of the double dividend: a reversal of early-life gains threatens the level advantage and may be difficult to correct once a cohort has passed through childhood, while erosion of adult environmental conditions threatens the slope and remains more tractable across the lifespan. Monitoring both components in younger cohorts, not overall cognitive performance alone, is therefore essential for anticipating future shifts in dementia incidence before they become visible in clinical statistics. If the double dividend is a primary mechanism currently contributing to holding back rising dementia burden, its two components require different early-warning indicators and different policy responses.

### Conclusion

Our findings reveal that the architecture of human cognitive aging is not a fixed biological inevitability, but a malleable process shaped by decades of societal progress. However, the double dividend currently protecting older generations is not a guaranteed constant. Recent plateaus and reversals of cognitive gains [44, 48] suggest that this progress may be fragile. If the environmental and social drivers of cognitive stability are allowed to erode, the current decline in dementia incidence could face a reversal. Our results demonstrate that the primary lever for population-level brain health lies in the continued and active enhancement of the lifelong conditions that build and sustain cognitive capacity and resilience, starting in early life.

## Methods

### Data assembly and harmonization

Episodic memory data were pooled from five large longitudinal studies: the Survey of Health, Ageing and Retirement in Europe (SHARE, release 9.0.0) [62, 63], the Health and Retirement Study (HRS, obtained from the University of Michigan, RAND longitudinal file 1992–2022) [64], the English Longitudinal Study of Ageing (ELSA, waves 1–11) [40, 52], the Irish Longitudinal Study on Ageing (TILDA, waves 1–6, accessed via the Irish Social Science Data Archive (www.ucd.ie/issda) [65-67], and the Longitudinal Aging Study Amsterdam (LASA, cycles B–K plus 2B and 3B, 1992–2021) [68]. All studies administered a word-list episodic memory task with immediate and delayed recall components. Scores were harmonized to a common 0–20 metric: ELSA, HRS, and SHARE use a 10-word single-trial paradigm already on the 0–20 scale; TILDA uses the same scale (second immediate trial discarded for comparability); LASA uses a 15-word three-trial paradigm rescaled by a factor of 2/3 to the common metric. Study membership was retained as a fixed covariate in all models (SI Section 1).

### Practice correction and inclusion criteria

To reduce retest inflation, practice effects were estimated separately for each study by regressing total recall on a flexible age function (natural spline, 8 df) and a log-transformed prior-test count, and then subtracting the estimated practice-related gain from each observation. Baseline age was restricted to 50–95 years. Person-level analyses required a minimum of four eligible assessments; precision diagnostics motivating this threshold are shown in SI Figure S3 (SI Sections 2–4).

### Person-level memory level and change estimates

For each participant meeting the inclusion criteria we estimated two summary statistics from their longitudinal practice-corrected recall record. Memory level was defined as the robust fitted value at the individual’s mid-observed age, derived from a Theil–Sen regression (median of all pairwise slopes) of recalled words on age. Memory change was the Theil–Sen slope itself (words/year). Slope uncertainty was quantified by a leave-one-out jackknife standard deviation (SI Section 4).

### Cohort trajectory estimation (Figure 1)

Cohort-stratified trajectories of level and change across overlapping age ranges were estimated by generalised additive models (GAMs) fitted to the person-level summary statistics, using a shared smooth of age and cohort intercepts, adjusted for study. Predictions are referenced to ELSA at baseline; curves are masked where any cohort × age-bin cell contains fewer than 50 participants. Sex-stratified versions using 10-year cohort bins are shown in SI Section 5.

Identifiable nonlinear cohort components (Figure, 2^nd^ row) were estimated in two steps. First, residuals from the canonical solution model (weighted linear regression of the outcome on age, birth year, and study) were computed, removing the unidentified linear APC drifts. Penalised thin-plate regression splines with null spaces removed (m=c(2,0), k=8) were then fitted to these residuals for age, birth year, and period (= age + birth year) using mgcv::gam() with REML estimation. The m=c(2,0) specification ensures each smooth captures only pure nonlinear deviations with no intercept or linear component. Under the no-interaction assumption, these components are identified from the data without bounding assumptions (SI Section 12).

### Age–Period–Cohort bounding analysis

Because calendar year equals age plus birth year, linear age, period, and cohort drifts cannot be simultaneously identified from observational data. We addressed this using the bounding framework of Fosse and Winship (2019) [31]. The identified composites *A* = bA + bP and C* = bC + bP*, where bA, bP, and bC denote the linear drift per unit increase in age, period, and birth year respectively, were estimated from weighted regression of the person-level outcome on age, birth year, and study fixed effects. The canonical solution line, the one-dimensional family of (bA, bP, bC) triples all fitting the data equally well, is then parameterised by the assumed period drift *bP: bA = A\** − *bP and bC = C\** − *bP* Rather than selecting a single point on this line, we report cohort effects across an explicit grid of assumed period drifts, expressed as fractions of C* — the coefficient on birth year from a weighted regression that includes age and study fixed effects, and therefore the birth-year trend after accounting for age differences. Because the exact dependency P = A + C means period cannot be simultaneously controlled, C* bundles the true cohort effect together with any co-occurring period trend (C* = bC + bP); it represents the maximum attributable cohort effect under the sign constraints, and equivalently the period drift that would be required to eliminate the cohort finding entirely, hence the tipping point (SI Section 12).

To empirically inform which portion of this grid is most plausible, we additionally estimated the long-run linear period drift implied by penalised cubic regression splines (bs = ‘cr’ in mgcv) of varying flexibility (k = 3, 6, 12), fitted within GAMs of the form: outcome ∼ age + birth_year + s(period, bs=‘cr’, k=K) + study, using mgcv::bam() with fREML estimation. The linear drift implied by each fitted spline was extracted by predicting across the observed period range at median covariate values and regressing predictions on calendar year, yielding a model-implied estimate of bP for each k. Specifications implying bA > 0 were excluded as violating Constraint (i). The GAM-implied values serve as empirical calibration points on the grid, indicating where within the admissible range the data place the period effect under flexible but not unconstrained modelling, rather than as identifying assumptions. Robustness was assessed by a tipping-point calculation and bootstrap resampling (B = 200, within-study) (SI Section 12).

### Decomposition and clinical calibration

The total 20-year cohort advantage at age 80 was decomposed into a level component (cohort difference already present at age 60) and a slope component (additional advantage accumulated through slower decline between ages 60 and 80). Clinical calibration used ADNI data (n = 813; 229 CN, 389 MCI, 195 AD) with recall scores rescaled to the 0–20 metric, providing empirical benchmarks for the CN–MCI and CN–AD gaps against which cohort advantages were expressed. Details are in SI Section 9.

### Clinical validation in HRS

To validate the clinical relevance of memory slope estimates within the primary data, we linked person-level Theil–Sen slopes from HRS (n = 22,905 with ≥4 waves) to the Langa– Weir cognitive classification at each participant’s final observed wave, defining impairment as CIND or dementia combined. The association between slope category and impairment was quantified by logistic regression adjusting for age and birth decade, with mediation estimated by comparing cohort coefficients with and without slope adjustment (SI Section 17).

### Distributional analyses and quantile regression

Cohort differences across the full slope distribution were estimated by quantile regression at five quantiles (τ = 0.10, 0.25, 0.50, 0.75, 0.90), adjusting for mid-age and study: *Q_τ(slope) = α_τ + β_τ · birth_decade + γ_τ · age_mid_c + Σ δ_τk · study_k*. Standard errors were obtained by bootstrap resampling of observation pairs (R = 500). The heatmap analyses of decline category prevalence by cohort and age used the same person-level slope estimates discretized into four categories (flat ≥ −0.05; mild −0.05 to −0.30; moderate −0.30 to −1.0; severe ≤ −1.0 words/year). Details are in SI Section 10.

### Bout analyses of sustained stability and steep decline

To characterise the temporal organisation of individual trajectories, we applied a windowed bout classification. For each participant, every consecutive window of W assessments was evaluated: a window was classified as a stable bout if the Theil–Sen slope was ≥ −0.05 words/year and the probability of the true slope exceeding this threshold (under a normal approximation using the jackknife SD) was ≥ 0.50. Steep-decline bouts used the analogous criterion with a ≤ −1.0 words/year threshold. Bout prevalence as a function of age was modelled by binomial GAMs with a shared age smooth and cohort fixed effects, adjusting for follow-up structure. Sensitivity was assessed by varying W (4–8), the stability threshold (−0.05 to −0.50 words/year), and a complementary overall-slope threshold classification. Full specification details are in SI Section 11.

### Neurobiological validation

Bilateral hippocampal volume change rates were estimated in an independent subsample with longitudinal MRI (n = 1,925; ≥2 scans, ≥4 memory assessments). Site-adjusted Theil– Sen slopes of bilateral hippocampal volume (left + right FreeSurfer segmentation) were computed per participant and expressed as percent change per year relative to estimated volume at median scan age. Outliers exceeding ±5%/year or ±4 SD were excluded. Memory slope categories were assigned using percentile-based thresholds matching the main-sample category proportions. Hippocampal atrophy rates were compared across categories by linear regression adjusting for age, sex, and intracranial volume, weighted by MRI follow-up span (SI Section 19).

### Software

All analyses were conducted in R. GAMs were fitted using mgcv::bam() with fREML estimation. Quantile regression used the quantreg package. Theil–Sen slopes were computed using a custom implementation of the pairwise-median estimator. Visualizations used ggplot2.

## Data Availability

All data used in the study were available before the initiation of the study and can be obtained through applications to data owners specified below.
https://share-eric.eu/
https://hrs.isr.umich.edu/about
https://www.elsa-project.ac.uk/
https://www.ucd.ie/issda/
https://lasa-vu.nl/en/

https://share-eric.eu/

https://hrs.isr.umich.edu/about

https://www.elsa-project.ac.uk/

https://www.ucd.ie/issda/

https://lasa-vu.nl/en/

## Acknowledgement

LCBC is supported by the European Research Council under grant agreements no. 283634 and no. 725025 (to A.M.F.) and no. 313440 (to K.B.W.), as well as the Norwegian Research Council (325878, 262453 to A.M.F.; 325001, 301395, 239889 to K.B.W.; 249931 to A.M.F & K.B.W.; 324882 to DVP; 325415 to HG), the National Association for Public Health’s dementia research program, Norway (to A.M.F.), and the University of Oslo through the UiO:Life Science convergence environment (to A.M.F). See SI Section 16 and 22 for funding of the different data sources.

