## Supplemental Information for "Generational gains in memory capacity and stability may account for declining dementia incidence rates in Europe and the United States"

| Section | Title | Page |
| --- | --- | --- |
| 1 | Data assembly and harmonization across longitudinal studies | 2 |
|  | Table S1. Sample characteristics by study on native recall scales | 4 |
| 2 | Practice correction and baseline restriction | 4 |
|  | Figure S1. Practice correction diagnostics by study | 6 |
| 3 | Person-level slope estimation and support diagnostics | 6 |
|  | Figure S2. Distributions of robust person-level memory slopes by study | 8 |
| 4 | Person-level robust level and change estimates, and uncertainty | 8 |
|  | Figure S3. Precision and qualitative robustness of person-level Theil–Sen | 10 |
|  | Figure S4: Testing Theil-Sen slope robustness in synthetic data | 11 |
| 5 | Sex-stratified analyses | 13 |
|  | Figure S5. Sex-stratified cohort effects on episodic memory level and slope | 13 |
| 6 | Growth-curve model for trajectory visualization | 14 |
|  | Figure S6. Predicted memory trajectories by birth cohort from the growth curve | 15 |
| 7 | Distributional diagnostics of memory-change slopes | 16 |
|  | Figure S7. Median-centered distributions of individual memory-change slopes | 16 |
| 8 | Sensitivity analyses | 17 |
|  | Figure S8. Robust cohort profiles in memory level and change | 17 |
|  | Figure S9. Sensitivity to selective inclusion in the ≥4-wave sample | 18 |
| 9 | Calibration of the generational memory advantage & clinical benchmarks | 18 |
|  | Table S2. Sample descriptives for the ADNI subsample | 19 |
|  | Table S3. Memory scores in the ADNI subsample | 19 |
|  | Table S4. Diagnostic accuracy of episodic memory for clinical groups | 19 |
| 10 | Quantile regression of birth cohort effects across the slope distribution | 19 |
| 11 | Bout analyses of sustained stability and steep decline | 20 |
|  | Figure S10: Prevalence of sustained stable memory bouts | 22 |
|  | Figure S11. Prevalence of sustained steep decline in episodic memory | 23 |
| 12 | Age–Period–Cohort bounding analysis: full technical details | 24 |
|  | Table S6: Cohort effect on memory level under a grid of assumed period drifts | 26 |
|  | Table S7: Cohort effect on memory slope under a grid of assumed period drifts | 26 |
|  | Figure S12: Age–period–cohort decomposition for memory level | 28 |
|  | Figure S13: Age–period–cohort decomposition for memory slope | 29 |
|  | Figure S14. Bootstrap distribution of bounded cohort drift in memory level | 30 |
|  | Figure S15. Bootstrap distribution of bounded cohort drift in memory slope | 31 |
| 13 | Mortality sensitivity analysis for quantile regression | 32 |
|  | Table S5: Quantile regression results: full vs mortality-restricted sample | 33 |
|  | Figure S16: Quantile regression cohort coefficients | 34 |
|  | Figure S17: Change in cohort coefficient (restricted minus full) at each quantile | 35 |
| 14 | Effect size comparisons with previous studies | 35 |
| 15 | Consortia authors and cohort information | 36 |
| 16 | Acknowledgement and funding SHARE, HRS, ELSA, TILDA, LASA | 37 |
| 17 | Clinical validation: Langa–Weir cognitive status in HRS | 39 |
| 18 | Identifiable nonlinear APC components | 41 |
|  | Table S8. Nonlinear APC smooth terms - memory level | 42 |
|  | Table S9. Nonlinear APC smooth terms - memory slope | 43 |
|  | Figure S18. Identifiable nonlinear APC components | 43 |
| 19 | Neurobiological validation - hippocampal atrophy | 43 |
| 20 | Replication under an independent Bayesian hierarchical framework | 44 |
| 21 | MRI subsample – sample and test characteristics | 46 |
|  | Table S10. Sample characteristics for the MRI subsample | 46 |
|  | Table S11. Memory tests and MRI scanner acquisition parameters | 47 |
| 22 | Sub-sample funding - MRI | 51 |

**Section 1: Data assembly and harmonization across longitudinal studies**

We pooled episodic memory data from five large longitudinal studies and harmonized them into a common long-format dataset with one row per person per assessment. The goal of this preprocessing stage was to extract a minimally sufficient and transparently defined set of variables for subsequent age–period–cohort and longitudinal change analyses while minimizing study-specific assumptions, particularly for interview timing and birth cohort assignment. All preprocessing was implemented in R in a single stand-alone script using data.table-based operations to read the original study files, perform study-specific transformations, and output a combined "super-dataset" for downstream analyses.

Across all studies we assembled a core set of variables including a study indicator (study), a harmonized person identifier (id), a wave/assessment indicator (wave), interview year (int_year), age at assessment (age), birth year (birth_year), and sex (sex). Episodic memory outcomes were represented as immediate recall (recall_immediate), delayed recall (recall_delayed), and total recall (recall_total), where recall_total was computed as recall_immediate plus recall_delayed. Native study identifiers were retained alongside the harmonized id to ensure reproducibility and facilitate merging of study-specific supplements; these included mergeid for SHARE, hhidpn for HRS, idauniq for ELSA, and id_raw for TILDA. Unless otherwise noted, negative-coded values and other special missing codes were converted to NA. At this stage we did not apply an analytic age-range restriction (e.g., ≥50 years), because this is applied later in the modeling pipeline. Instead, we retained observations only when age, birth_year, and recall_total were all finite after study-specific cleaning and harmonization.

For SHARE, we used the easySHARE release 9.0.0 file easySHARE_rel9_0_0.rda as the primary input. From this file we extracted mergeid, wave, int_year, age, dn003_mod (birth year), female (sex), and episodic memory variables recall_1 (immediate) and recall_2 (delayed). The harmonized SHARE id was formed as "SHARE_" concatenated with mergeid, and mergeid itself was retained for subsequent merges. A key design choice for SHARE was to avoid deriving interview timing or age when study-provided variables were available; therefore int_year and age were taken directly from easySHARE rather than recomputed. To support sensitivity analyses related to mortality, end-of-life (EOL) interviewing, and attrition, we additionally merged two supplementary files stored in the SHARE directory, df.eol.rds and df.eolbywave.rds. The df.eol file is person-level and was merged to SHARE by mergeid; df.eolbywave is person-wave and was merged by mergeid and wave. Prior to merging, the script enforced key uniqueness (one row per mergeid in df.eol; one row per mergeid–wave in df.eolbywave) and performed left joins to preserve all SHARE observations. The merged EOL fields retained include, where present, year of EOL interview, death year and month, last core interview wave and year, proxy and interview-mode indicators, and wave-level death/attrition indicators.

For HRS, we used the RAND HRS Longitudinal File randhrs1992_2022v1.dta. The harmonized HRS id was formed as "HRS_" concatenated with hhidpn, and hhidpn was retained. Birth year was taken from rabyear and sex from ragender. A central design choice for HRS was to ensure interview timing was taken from an explicit interview-year variable rather than left missing or inferred indirectly. Accordingly, int_year was derived from wave-specific interview end year variables r#iwendy, which were reshaped from wide to long and merged by hhidpn and wave. Age at assessment was taken from wave-specific r#agey_e. Episodic memory was extracted from wave-specific recall variables, reshaped to long format. For earlier waves immediate and delayed recall were taken from r#imrc and r#dlrc. For later waves with alternative variable naming conventions (including imputed and word-list variants), immediate and delayed recall were taken from the first non-missing value among the available alternatives (e.g., r#imrcp or r#imrcw for immediate recall and r#dlrcp or r#dlrcw for delayed recall), with negative-coded values set to missing.

For ELSA, we used a hybrid approach to maximize the number of waves while keeping variable extraction explicit and minimally assumption-dependent. Waves 1–10 were extracted from the Gateway/harmonized "fat file" gh_elsa_h.dta, while wave 11 was appended from the raw wave 11 file wave_11_elsa_data_eul_v1.dta. The harmonized ELSA id was formed as "ELSA_" concatenated with idauniq, and idauniq was retained. In gh_elsa_h.dta, interview year was taken from rWiwy (wave-specific r1iwy through r10iwy), age from rWagey (r1agey through r10agey), immediate recall from rWimrc, and delayed recall from rWdlrc, each reshaped to long format. Birth year was taken from rabyear and sex from ragender. For wave 11, the raw wave_11_elsa_data_eul_v1.dta file was used with verified variable names: idauniq as the identifier, iintdaty as interview year, indager as age, indobyr as birth year, indsex as sex, and cflisen and cflisd as immediate and delayed recall. Note: the age variable in the harmonized file contained recording errors for 298 participants in which age stopped updating across later waves. For 294 of these participants, age was recomputed as int_year − birth_year where the discrepancy exceeded 2 years. Four participants with inconsistent birth_year values across waves were excluded entirely.

For TILDA, we used wave-specific public microdata (.dta) files in the downloaded TILDA directory and extracted waves 1–6 where file naming allowed wave identification. The harmonized TILDA id was formed as "TILDA_" concatenated with the native identifier id (retained as id_raw). Episodic memory was taken from derived cognition variables COGimmediatedrecall1 and COGdelayedrecall. We intentionally used the first immediate recall trial (COGimmediatedrecall1) together with delayed recall to maximize comparability with other studies and did not include the second immediate trial. Sex was taken from sex when present, otherwise gd002 if available. A key design choice for TILDA was the treatment of interview timing and birth cohort assignment. Because per-person interview dates/years and birth-year variables were not consistently available across public microdata in a manner comparable to the other studies, we used a transparent wave-level interview-year proxy, int_year_proxy, based on the documented fieldwork timing of each wave. A corresponding integer interview year was then defined as int_year = round(int_year_proxy). Birth year was derived at each observation as birth_year_row = int_year_proxy minus age. We then stabilized birth_year for each person as the rounded median of birth_year_row across that person's observations. To avoid implausible inferred birth-year instability, we excluded individuals with three or more observations whose within-person range of birth_year_row exceeded three years, and then recomputed the stabilized birth_year among retained individuals.

For LASA, we used a set of cycle-specific SPSS (.SAV) files from the LASA data release (LASAfiles_20260226). The harmonized LASA id was formed as "LASA_" concatenated with the native participant identifier respnr, which was retained throughout. Demographic information was extracted from the person-level file LASAZ004.SAV, and medical age variables were extracted from LASAZ008.SAV. Episodic memory was extracted from cycle-specific files following the naming convention LASA{cycle}356.SAV (or LAS{cycle}356.SAV for new-cohort baseline cycles 2B and 3B). LASA uses a three-trial word-list learning paradigm; immediate recall was computed as the mean across the three learning trials ({cycle}mtotal / 3) and delayed recall was derived as the retention percentage ({cycle}mret2pc / 100) multiplied by the maximum words recalled across learning trials ({cycle}mtmax). Total recall was then computed as immediate plus delayed recall. LASA covers twelve assessment cycles (B through K, plus new-cohort baselines 2B and 3B) spanning 1992 to 2021. Age at assessment was taken from the cycle-specific medical age variables (e.g., bmage, cmage) in LASAZ008.SAV for regular cycles; for the new-cohort baseline cycles 2B and 3B, age was computed from the interview date variable t2m_dat and birth year (byear) from the demographic file, expressed in decimal years. For cycle K, where the medical age variable was occasionally missing, age was computed analogously from t11_dat and byear. Birth year was taken from byear in LASAZ004.SAV, converted to integer, and used directly without further stabilization. Interview year was assigned using a cycle-level lookup table based on documented LASA fieldwork timing: 1992 (B), 1995 (C), 1998 (D), 2001 (E), 2002 (2B), 2005 (F), 2008 (G), 2011 (H), 2012 (3B), 2015 (I), 2018 (J), and 2021 (K).

The final output of this stage is a combined long-format file super_dat_ELSA_SHARE_HRS_TILDA_LASA_raw.rds, saved to the project's derived data directory, along with a study-level QC summary table. The combined dataset retains native IDs for traceability and for later merges of additional study-specific supplements.

| **Study** | **Participants** | **Observations** | **Interview years** | **Mean recall total (SD)** |
| --- | --- | --- | --- | --- |
| ELSA | 22,215 | 99,785 | 2002–2024 | 10.41 (3.69) |
| HRS | 38,554 | 230,163 | 1995–2021 | 9.84 (3.62) |
| SHARE | 155,801 | 444,192 | 2004–2022 | 8.97 (3.74) |
| TILDA | 8,116 | 34,879 | 2010–2022 | 12.10 (3.89) |
| LASA | 4,641 | 16,718 | 1992–2021 | 12.84 (5.01) |

***Table S1****. Sample characteristics by study on native recall scales, prior to analytic exclusions.*

*Each row summarises the raw analytic extract for one study prior to age-range restriction (≥50 years), practice-effect correction, and the minimum four-wave requirement applied in the longitudinal analyses. Participants are unique individuals; observations are person-wave rows with non-missing age, birth year, and total recall. Interview years reflect the range of assessment waves included. Mean recall total is reported on each study's native scale and is not directly comparable across studies (see note below); all analyses in the main text and SI use scores harmonized to a common 0–20 scale. Study membership is included as a fixed covariate in all primary analyses, absorbing mean-level differences between studies in both memory level and rate of change. Education was retained in the dataset for traceability but does not enter any primary analysis; its role as a candidate driver of cohort differences is discussed in the main text. Note that mean recall scores are not directly comparable across studies for two reasons. First, studies differ in the number of words presented and the number of learning trials administered. TILDA uses a two-trial immediate recall paradigm, participants hear the word list twice and recall it twice before the delayed test, which both inflates the immediate recall score relative to single-trial studies and improves delayed recall performance through the additional encoding opportunity. LASA uses a three-trial paradigm with the same consequence: the immediate recall component is averaged across three learning trials, and the additional exposures further boost delayed recall relative to single-trial designs. ELSA, HRS, and SHARE each use a single immediate recall trial followed by a single delayed recall trial. Second, studies differ in list length (ranging from 10 to 15 words depending on study and wave). These differences motivate the harmonization of all scores to a common 0–20 scale prior to analysis, which uses study-specific rescaling to render scores comparable across cohorts and studies.*

*Score harmonization to a common 0–20 scale*. Because studies differ in word list length and number of learning trials (see Table S1 note), raw recall totals are not directly comparable across studies. Prior to all analyses, total recall scores were harmonized to a common 0–20 metric. For ELSA, HRS, and SHARE, total recall is already on a 0–20 scale (10-word list, single immediate plus single delayed trial) and no rescaling was applied. For TILDA, the second immediate recall trial is discarded and only the first trial is used, so the effective score (0–10 immediate + 0–10 delayed) is also already on the 0–20 scale. For LASA, the 15-word list and three-trial learning paradigm require rescaling. Immediate recall was computed as the mean score across the three learning trials (range 0–15) and delayed recall as the retention percentage multiplied by the maximum words recalled across learning trials (range 0–15). The sum of these two components (range 0–30) was multiplied by 2/3 to map it onto the 0–20 scale, which is equivalent to rescaling each component proportionally to a 0–10 range before summing. The resulting variable recall_total_20 was used in all downstream analyses. Study membership is included as a fixed covariate in all models, absorbing any residual mean differences between studies that remain after harmonization.

**Section 2: Practice correction and baseline restriction**

To reduce retest-related inflation of episodic memory scores, we applied a baseline restriction and a study-specific practice correction to the pooled long-format dataset. Within each study and individual, observations were ordered chronologically using interview year (int_year), then wave, then age. Baseline age was defined as the age at the first observation in this ordered sequence. We then retained only individuals whose baseline age fell between 50 and 95 years, while keeping all subsequent observations for those individuals. Within the retained sample, we computed the number of prior tests for each observation as the count of earlier assessments for the same individual (prior_tests, starting at 0 at baseline).

Practice effects were estimated separately for each study using all eligible observations with non-missing total recall, age, and prior_tests. We fit a linear model in which total recall was regressed on a flexible age function (a natural spline of age with 8 degrees of freedom) and a practice term defined as the logarithm of one plus the number of prior tests, with the number of prior tests capped at 12 to prevent undue influence of very high retest counts. From each study-specific model we extracted the coefficient for the practice term. For every observation in that study, the expected practice-related gain was computed as the product of this coefficient and the log-transformed capped prior_tests. Practice-corrected recall was then defined as the raw total recall minus the estimated gain. The corrected dataset, including baseline age, prior_tests, and estimated gain, was saved for downstream analyses. Diagnostic plots were produced for each study showing the distribution of practice-corrected recall and the estimated practice correction as a function of prior_tests; these were additionally combined into a 2×3 panel for visual comparison across studies.

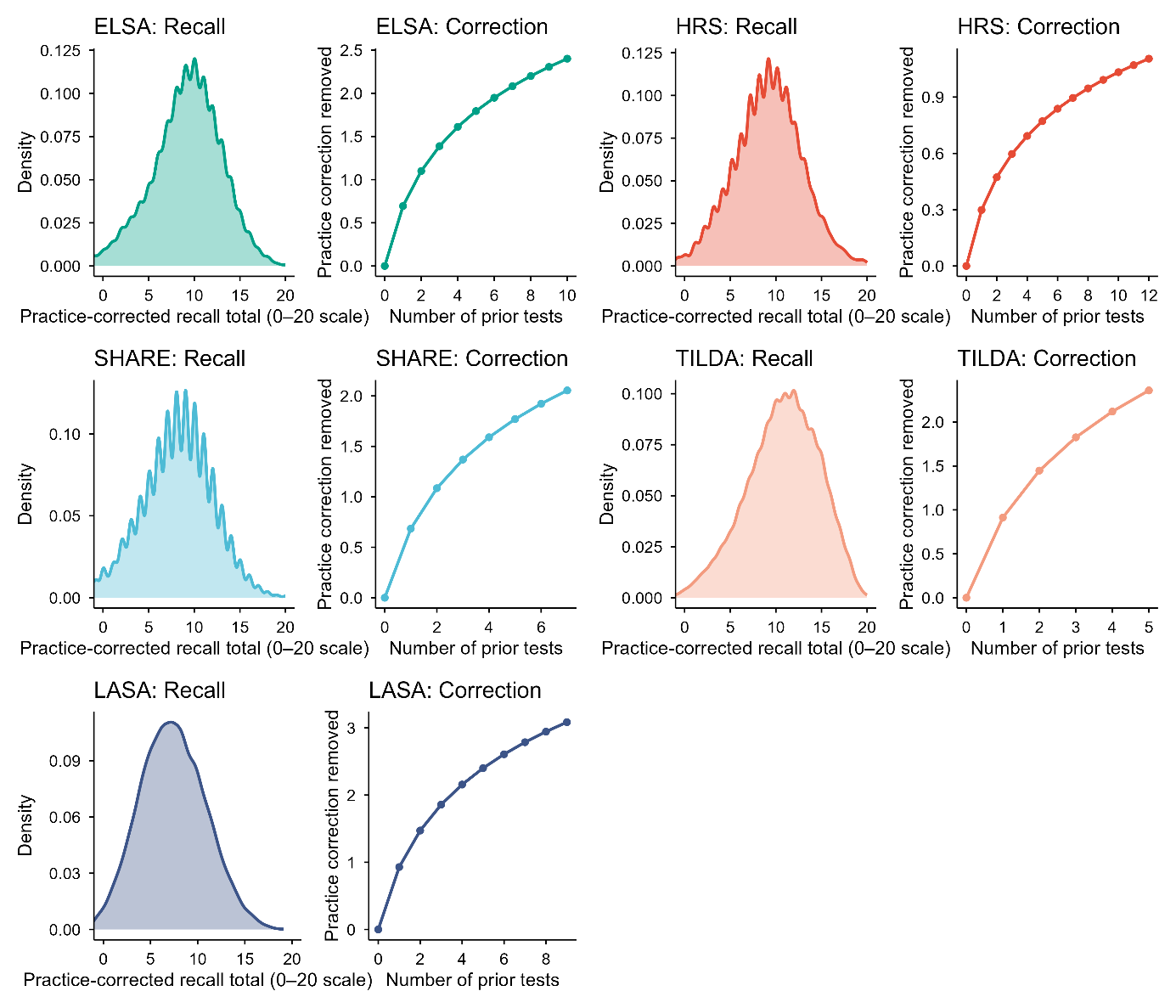

***Figure S1****. Practice correction diagnostics by study.* For each study, the left panel shows the density of practice-corrected total recall (immediate plus delayed recall) after subtracting the estimated retest-related gain. The right panel shows the mean estimated practice correction removed (in words) as a function of the number of prior tests, with shaded bands indicating 95% confidence intervals based on the standard error of the mean at each prior-test count. Panels are arranged in a 2×2 layout to facilitate comparison across ELSA, HRS, SHARE, TILDA and LASA.

**Section 3: Person-level slope estimation and support diagnostics**

To characterize within-person memory change in a way that is robust to measurement noise and irregular spacing of assessments, we estimated person-specific slopes using the practice-corrected long-format dataset produced in the preprocessing stage. Analyses used the practice-corrected total recall score (recall_corr) when available; otherwise total recall (recall_total) was used. Observations were restricted to those with non-missing age, birth year, and the selected memory score. Within each study, individuals were retained if they had at least four eligible assessments (MIN_OBS = 4). For each retained individual, we estimated a robust slope of memory versus age using a Theil–Sen approach based on the median of all pairwise slopes across that person’s observations. This yielded a person-level rate of change in memory in units of words per year of age (slope_ts). To quantify the stability of the individual slope estimate, we also computed a leave-one-out jackknife standard deviation (sd_jk) of the Theil–Sen slope by repeatedly re-estimating the slope after removing each observation in turn (available for individuals with at least four observations). For descriptive purposes and downstream support checks, we additionally computed each person’s mid-observed age (age_mid; the midpoint of the individual’s observed age range) and the span of observed ages (span_years).

Birth cohorts were grouped into 5-year bins based on birth_year by rounding down to the nearest multiple of five, and labeling the bin as a 5-year interval (by_label; e.g., 1930–1934). To support later visualization steps and avoid extrapolation into poorly supported age–cohort regions, we constructed a support table that counts the number of individuals contributing slope estimates within each birth-cohort bin and mid-age bin combination. Mid-age was binned in one-year increments (age_mid_bin; AGE_BIN = 1). The support table records n_people for each by_label × age_mid_bin cell. For diagnostic visualization of overlap, we summarized this support table as the number of cohort bins represented at each age_mid_bin above a minimum support threshold (N_MIN_PER_AGEBIN = 50).

We produced study-specific density plots of the distribution of person-level slopes (slope_ts) to summarize heterogeneity in within-person change rates and to facilitate cross-study comparison. To ensure comparability across studies, all density plots used a common x-axis range from −2 to +2 words per year and included a vertical reference line at zero. The four study-specific density plots were also combined into a 2×2 panel layout to enable direct visual comparison across studies. The person-level slope table (including slope_ts, sd_jk, age_mid, span_years, birth_year and cohort bin labels) and the support table were saved for downstream modeling and plotting.

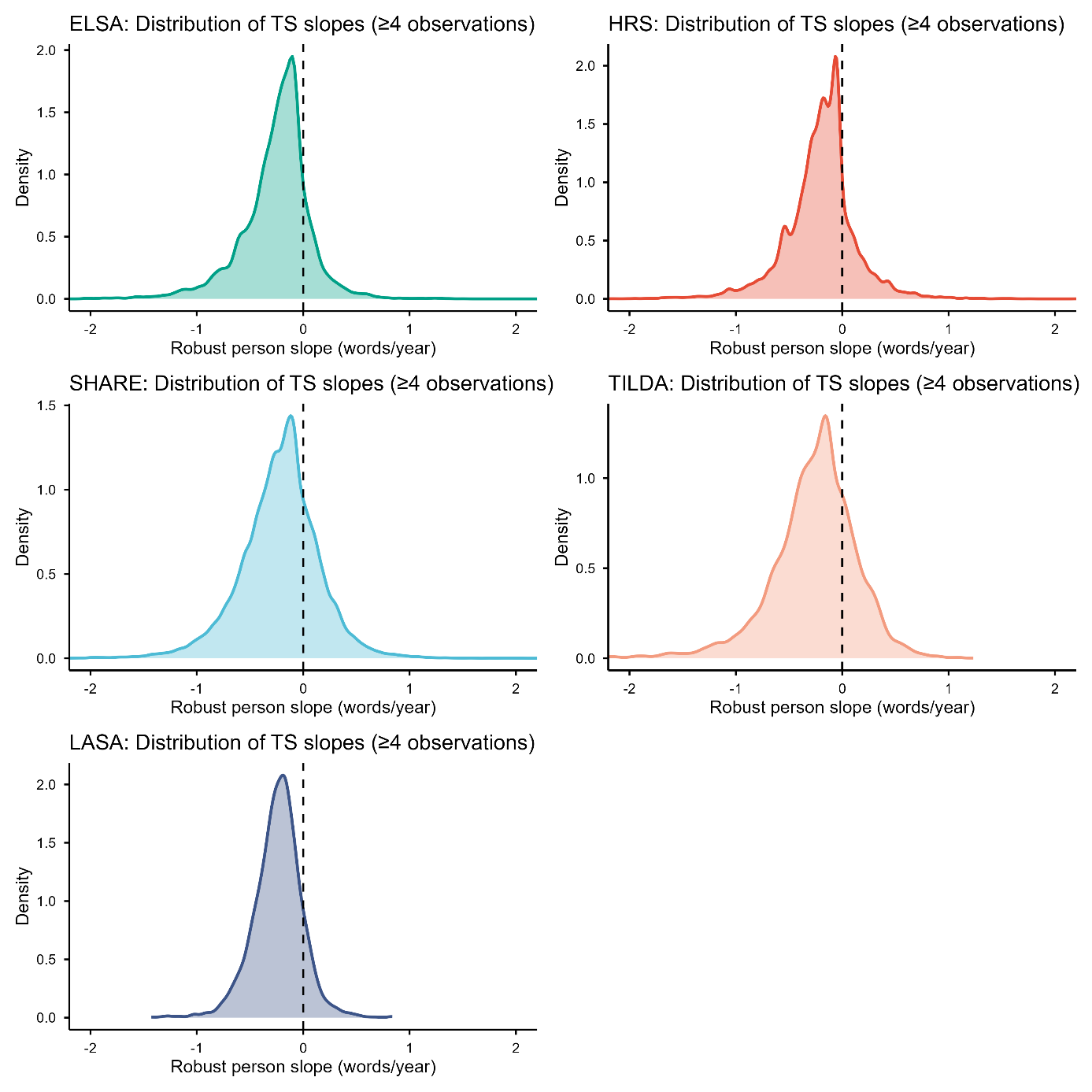

***Figure S2****. Distributions of robust person-level memory slopes by study. Each panel shows the density of person-specific rates of change in episodic memory (slope_ts; words per year of age) estimated using a Theil–Sen procedure applied to practice-corrected total recall scores across repeated assessments. The vertical dashed line indicates zero change. All panels share the same x-axis limits (−2 to +2 words/year) to enable direct comparison of the distributional shape and spread across ELSA, HRS, SHARE, TILDA and LASA.*

**Section 4: Person-level robust level and change estimates, and uncertainty quantification**

Overview and motivation Repeated episodic memory test scores are noisy at the individual level due to imperfect test reliability, day-to-day fluctuations, and occasional outlying responses. In addition, participants contribute uneven numbers of assessments and have unequal follow-up durations. Because the aims of the study require person-level estimates of both (i) memory level and (ii) memory change—together with uncertainty estimates that can be propagated into downstream analyses—we used robust person-level summaries rather than relying exclusively on population-level smooth trajectories.

Practice correction To reduce bias from retesting, we first derived practice-corrected memory scores. Let y_ij denote the observed memory score for participant i at assessment j, and let p_ij denote the number of prior memory assessments completed by participant i before assessment j. We fit an ordinary least squares model to the full dataset, adjusting for age with a flexible function and practice exposure with a monotonic diminishing-returns term. The fitted model yields an expected practice component, which was subtracted from y_ij to obtain practice-corrected scores y_ij_corr. These practice-corrected scores were used for all person-level summaries.

Theil–Sen slope (robust change estimate) For participant i with n_i repeated observations at ages a_i1, …, a_in and practice-corrected scores y_i1_corr, …, y_in_corr, we estimated change using the Theil–Sen slope. Define all pairwise slopes:
b_i(j,k) = (y_ik_corr − y_ij_corr) / (a_ik − a_ij), for 1 ≤ j < k ≤ n_i and a_ik ≠ a_ij.
The Theil–Sen slope is the median of these pairwise slopes:
b_i_TS = median{ b_i(j,k) }.
This estimator is robust to outliers and does not rely on normally distributed errors.

Robust memory level at a common within-person reference age To summarise memory level independently of change, we evaluated each participant’s robust level at the midpoint of their observed age range:
a_i_mid = (min_j a_ij + max_j a_ij) / 2.
We then defined a participant-specific level estimate as the robust fitted value at a_i_mid based on the Theil–Sen line, i.e., using the estimated slope together with a robust intercept (equivalently, the median of y_ij_corr − b_i_TS * a_ij). This yields a person-level level estimate that is comparable across individuals even when measurement schedules differ.

Jackknife uncertainty for person-level slopes To quantify uncertainty in b_i_TS given each participant’s observation pattern, we used a leave-one-out jackknife. For participants with n_i ≥ 3, we recomputed Theil–Sen slopes leaving out one observation at a time:
b_i(-j) = Theil–Sen slope computed from { (a_ik, y_ik_corr) : k ≠ j } for j = 1,…,n_i.
The jackknife standard error was computed as:
SE_i_JK = sqrt( (n_i − 1) * var{ b_i(-j) } ),
where var is the sample variance across the leave-one-out slopes. (For n_i = 2, a jackknife distribution of slopes is not defined because leaving out one observation leaves a single point.)

Empirical precision and sign-stability as a function of number of observations We evaluated how slope precision and qualitative robustness depend on the number of observations n_i. Two diagnostics were computed:

1. Precision (median jackknife SE by n_i). Among participants with n_i = 3, the median SE_i_JK was 1.05 words/year (IQR 0.48–1.93). With n_i = 4, the median SE_i_JK was 0.54 (IQR 0.32–0.85), representing an approximately 49% reduction in median uncertainty from 3 to 4 observations. Precision improved more gradually with additional observations (e.g., median SE_i_JK 0.38 at n_i = 5; 0.30 at n_i = 6; 0.18 at n_i = 8).
2. Sign stability (leave-one-out consistency). For each participant with n_i ≥ 3, we computed the fraction of leave-one-out slopes whose sign matched the sign of the full-sample slope:
   p_i_same = mean_j [ sign(b_i(-j)) = sign(b_i_TS) ].
   With n_i = 3, the median p_i_same was 2/3. With n_i = 4, median p_i_same was 1.0 (IQR 0.75–1.0), indicating that with four assessments the direction of estimated change is typically stable under leave-one-out perturbations. For higher n_i, median p_i_same remained 1.0 and the lower quartile increased further.

Together, these diagnostics show a clear improvement in both quantitative precision and qualitative robustness from 3 to 4 observations. We therefore required n_i ≥ 4 for analyses focused on person-level change and downstream cohort comparisons based on individual slopes.

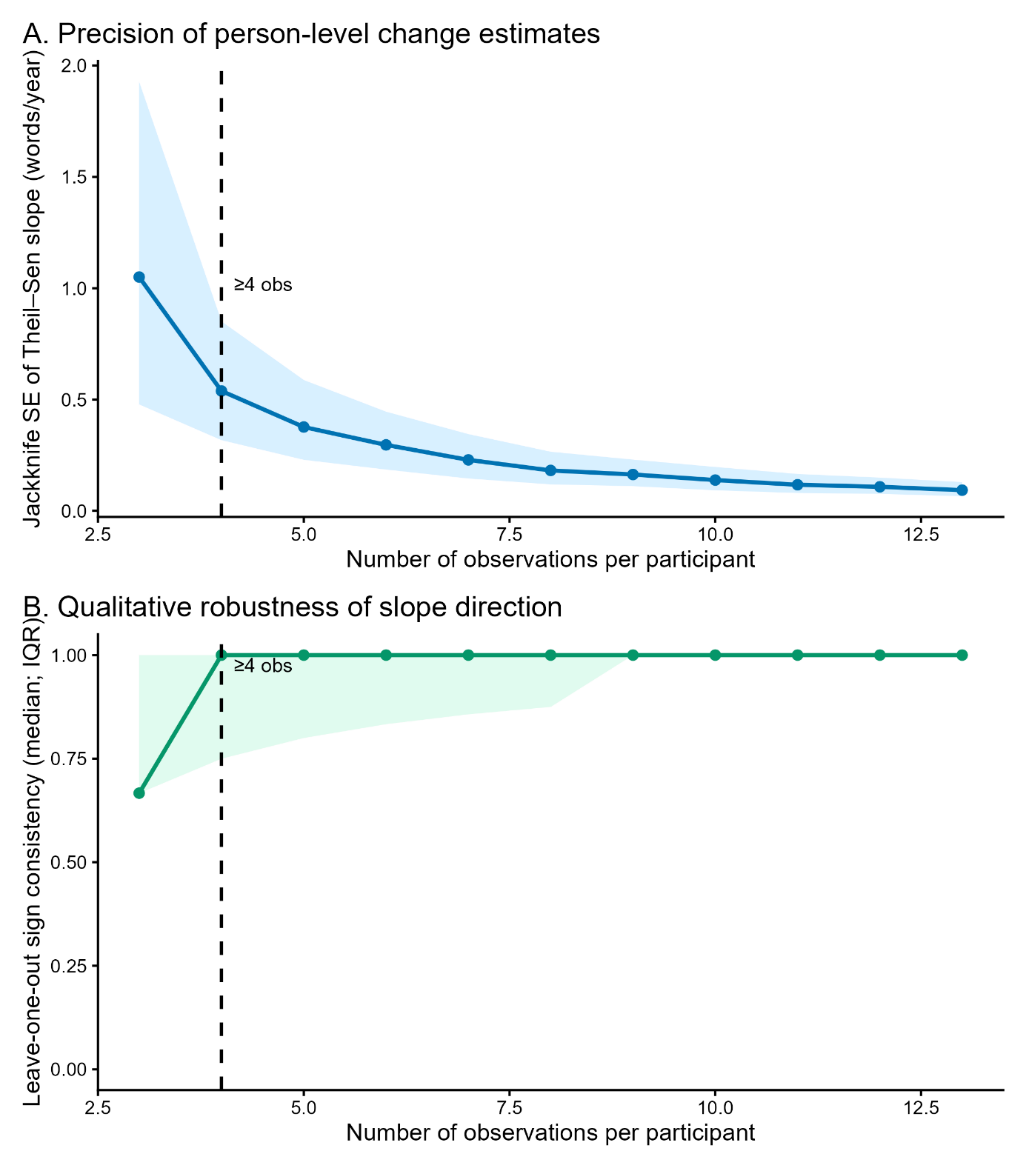

***Figure S3****. Precision and qualitative robustness of person-level Theil–Sen change estimates as a function of number of observations. We evaluated how the reliability of individual change estimates depends on the number of repeated episodic-memory assessments per participant (n_obs). For each participant with ages a_i1,…,a_in and practice-corrected scores y_i1_corr,…,y_in_corr, we estimated a robust Theil–Sen slope (words/year) as the median of all pairwise slopes: b_TS = median{ (y_ik_corr − y_ij_corr) / (a_ik − a_ij) } for all j<k. Uncertainty in b_TS was quantified using a leave-one-out jackknife: for each j, we recomputed the Theil–Sen slope leaving out observation j to obtain b_(-j), and then computed the jackknife standard error as SE_JK = sqrt( (n_obs − 1) * var(b_(-j)) ). These analyses did not include LASA data to avoid risk that the different memory test format could bias the estimates.*

*Panel A (Precision): The solid line shows the median SE_JK across participants within each n_obs; the shaded band shows the interquartile range (25th–75th percentiles). Precision improves sharply from three to four observations (median SE_JK decreases from 1.05 to 0.54 words/year; IQR 0.48–1.93 vs 0.32–0.85), with more gradual gains thereafter.*

*Panel B (Qualitative robustness): For each participant, we computed leave-one-out sign consistency p_same = mean_j[ sign(b_(-j)) = sign(b_TS) ], i.e., the fraction of jackknife slopes whose sign matched the full-sample slope. The solid line shows the median p_same by n_obs and the shaded band the interquartile range. Slope direction is only moderately stable with three observations (median p_same = 2/3) but becomes markedly more stable with four observations (median p_same = 1.0; IQR 0.75–1.0).*

*The vertical dashed line marks the pre-specified inclusion threshold (n_obs ≥ 4) used for person-level change analyses in the main text.*

*Evaluation of Theil–Sen regression and jackknife uncertainty estimation using a simulation experiment*

We evaluated the performance of robust linear trend estimation using the Theil–Sen estimator under varying signal-to-noise ratios (SNRs) and sampling densities. Synthetic time series were generated for n = 1000 samples with randomly assigned intercepts and slopes over 10 discrete time points. Gaussian noise was added with variance scaled by predefined SNR levels (0.0625–2), and 10% of observations were randomly selected as outliers and the noise was amplified fivefold to simulate heavy-tailed noise contamination.

For each sample and SNR level, subsets of t time points (t = 2–10) were randomly selected without replacement, and slopes and intercepts were estimated using Theil–Sen regression. Estimation accuracy was quantified by computing the root-mean-square error (RMSE)  between estimated and true parameters across simulations. Uncertainty of slope estimates was assessed using a leave-one-out jackknife procedure. For each subset of time points, regression was repeated t times with one observation omitted at each iteration, and the standard deviation of the resulting slope estimates was used as an estimate of the standard error.

This framework enabled systematic evaluation of estimator robustness as a function of sampling density and noise level, and assessment of whether jackknife-derived uncertainty reflects improvements in slope estimation accuracy.

We found that the jackknife-derived standard error scaled proportionally with the empirical standard deviation of the slope estimates across simulations, expect for the case with 2 samples as expected, suggesting that the jackknife procedure provides a useful proxy for estimator uncertainty. In addition, we observed a pronounced reduction in RMSE when increasing the number of timesteps from three to four, corresponding to an approximate 35% decrease in estimation error. This improvement was consistent across signal-to-noise ratios, highlighting the importance of at least four temporal observations for stable Theil–Sen slope estimation.

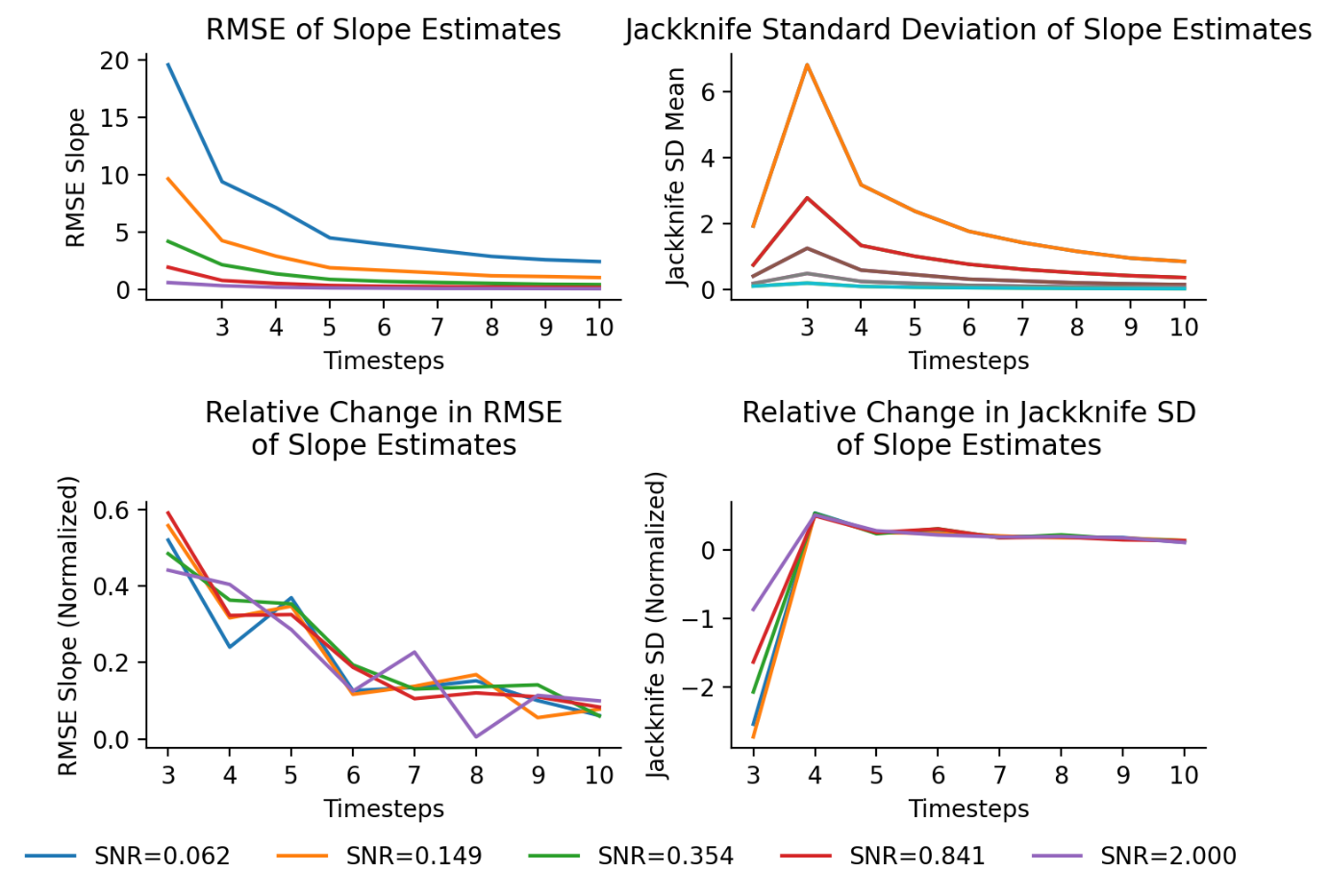

***Figure S4:*** *Testing Theil-Sen slope robustness in synthetic data*

**Section 5: Sex-stratified analyses**

The primary analyses pool across sex, treating study membership as the sole between-sample fixed covariate. To verify that this pooling does not obscure divergent cohort patterns in women and men, we repeated the Figure 1 analysis separately by sex using 10-year birth cohort bins to ensure adequate cell sizes after stratification. The goal was not to quantify sex differences in cohort effects, which would require a full sex × cohort interaction analysis across the 30+ countries represented and is deferred to future work, but to confirm that the cohort ordering in both memory level and slope is preserved within each sex, providing empirical justification for the pooled approach taken throughout.

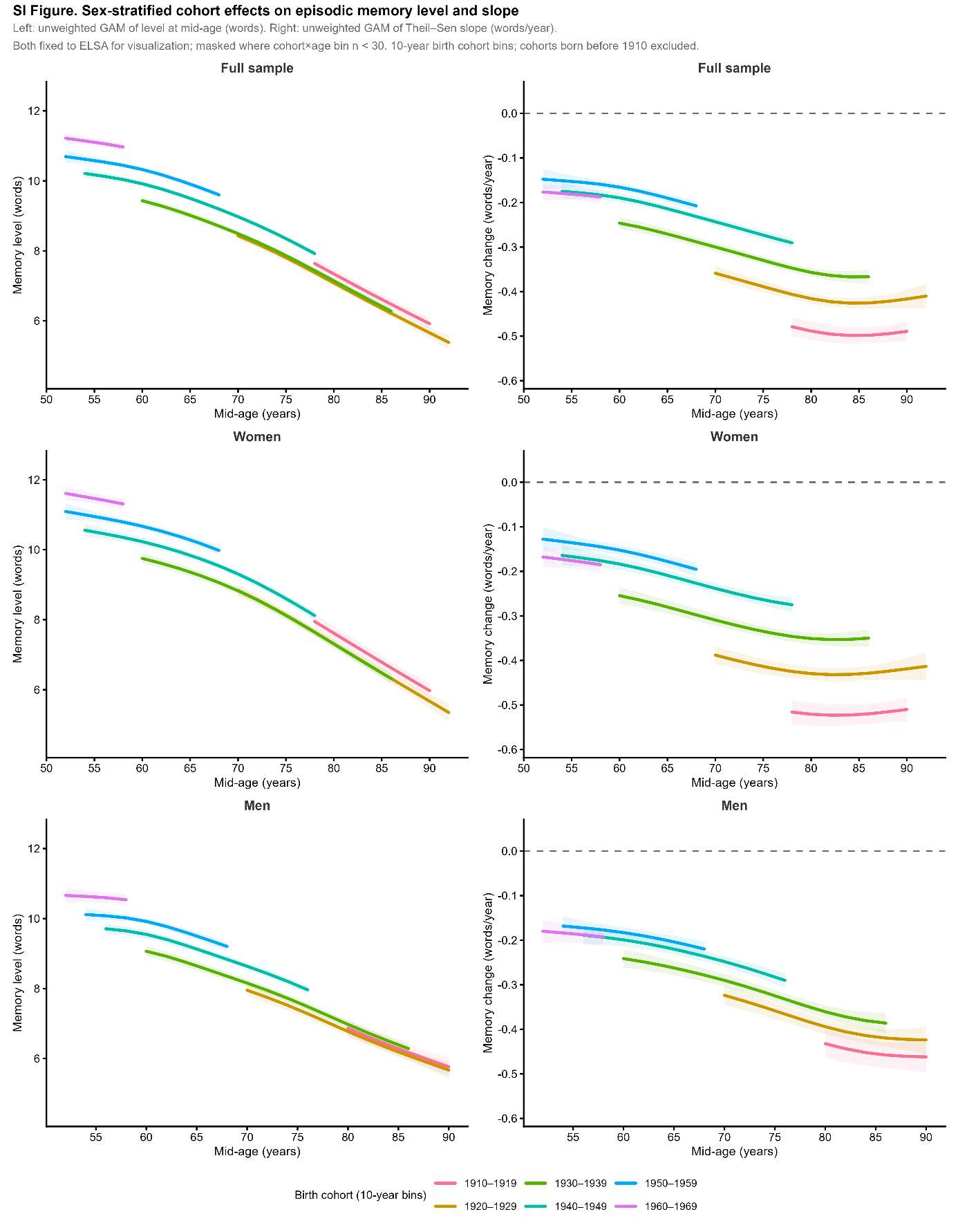

***Figure S5.*** *Sex-stratified cohort effects on episodic memory level and slope across birth cohorts.* *Each row shows results for the full sample (top), women (middle), and men (bottom). The left column shows memory level (words recalled on the 0–20 scale) and the right column shows the rate of memory change (words/year), both plotted as a function of mid-age across 10-year birth cohorts spanning 1910–1969. Cohorts born before 1910 are excluded due to insufficient sample sizes in sex-stratified analyses. The figure is the sex-stratified analogue of Figure 1 in the main text, using identical analytical procedures but 10-year rather than 5-year birth cohort bins to ensure adequate cell sizes after stratification.*

*Level panels (left column). For each participant, a person-level memory level estimate was computed as the predicted score at the individual's mid-age, derived from a Theil–Sen robust regression of recalled words on age using all available observations (minimum 4 waves). A GAM was then fitted to these person-level estimates: level ~ s(age_mid) + birth_cohort + study. Predictions are shown for ELSA as the reference study. Birth cohort enters as a factor, so each curve represents the age trajectory of one cohort after adjusting for a shared smooth of age.*

*Slope panels (right column). Person-level Theil–Sen slopes (words/year) were estimated from each participant's longitudinal record (minimum 4 waves). A GAM was fitted to these slopes: slope ~ s(age_mid) + birth_cohort + study, again with ELSA as the reference study. The dashed horizontal line marks zero change.*

*Masking. Curves are displayed only where the cohort × 2-year age bin contains at least 30 participants. This threshold is relaxed relative to the n ≥ 50 criterion used in the main figures to accommodate the smaller cell sizes resulting from sex stratification, particularly for men at older ages and for the earliest cohorts.*

*Sample. The total analytic sample contributing to this figure comprises 119,229 women and 99,959 men across five studies (SHARE, HRS, ELSA, TILDA, LASA), representing 219,188 individuals — effectively the full analytic sample, as sex information was available for virtually all participants. The female majority (approximately 54% of individuals) reflects the well-documented survival advantage of women in aging cohort studies.*

*Interpretation. The cohort ordering, later-born cohorts showing higher memory levels and less steep decline, is preserved in both sexes across all panels, with no reversals or crossovers that would undermine the main-effects approach used in the primary analyses. The cohort gradient is visible in both the level and slope dimensions for women and men alike, though the separation between cohorts is somewhat more pronounced among women, consistent with historically larger gains in educational attainment and labour-force participation for women across the cohorts studied. These patterns support the conclusion that the generational double dividend is not an artefact of sex composition differences across cohorts, and that a full sex × cohort interaction analysis, while potentially informative, would not alter the primary findings.*

**Section 6: Growth-curve model for trajectory visualization**

To provide an empirically grounded age-trajectory shape for Figure 4, we fitted a mixed-effects generalized additive model (gamm4) to the full longitudinal dataset (except LASA) (all cohort effect estimates used in the decomposition and public health calculations derive from the bounding analysis). The outcome was practice-corrected total recall. Fixed effects included study indicators, a log-transformed practice term (log(1 + prior tests), capped at 12), a penalized spline of centred age (s(age_c), k=10, edf=7.73), and a penalized spline of centred birth year (s(coh_c), k=6, edf=4.87). Random effects included a per-person intercept and the random (penalized) components of both splines. The model was fitted with period omitted, meaning the cohort smooth captures a mixture of cohort and period variance and should not be interpreted as an APC-corrected cohort effect. The model intercept (ELSA, mean age, mean cohort, no prior tests) was 9.24 words, and the between-person variance accounted for 33% of total variance (ICC = 0.33). The marginal R² = 0.20 (fixed effects only) is consistent with expectations for a population-level model of a highly variable cognitive outcome.

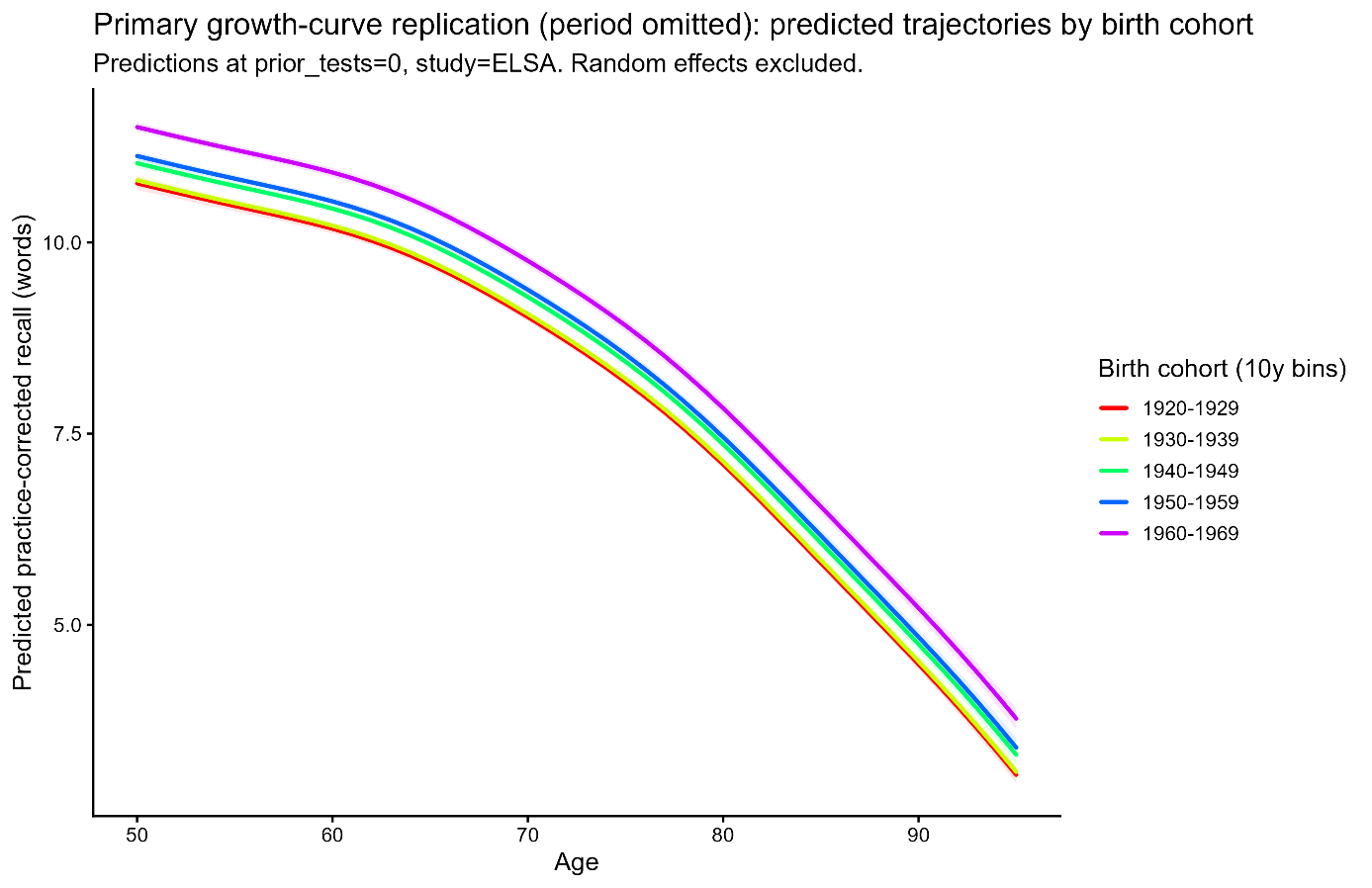

***Figure S6****. Predicted memory trajectories by birth cohort from the growth curve model (period omitted). Predicted practice-corrected total recall (words, 0–20 scale) as a function of age, shown separately for five 10-year birth cohorts (1920–1969). Predictions are evaluated at prior tests = 0 and study fixed to ELSA, with random effects excluded (population-average trajectories). The cohort separation visible here reflects a mixture of cohort and period variance, as period was omitted from the model; these trajectories are used solely to provide an empirically grounded age-trajectory shape for Figure 4 and should not be interpreted as APC-corrected cohort effects.*

**Section 7: Distributional diagnostics of memory-change slopes**

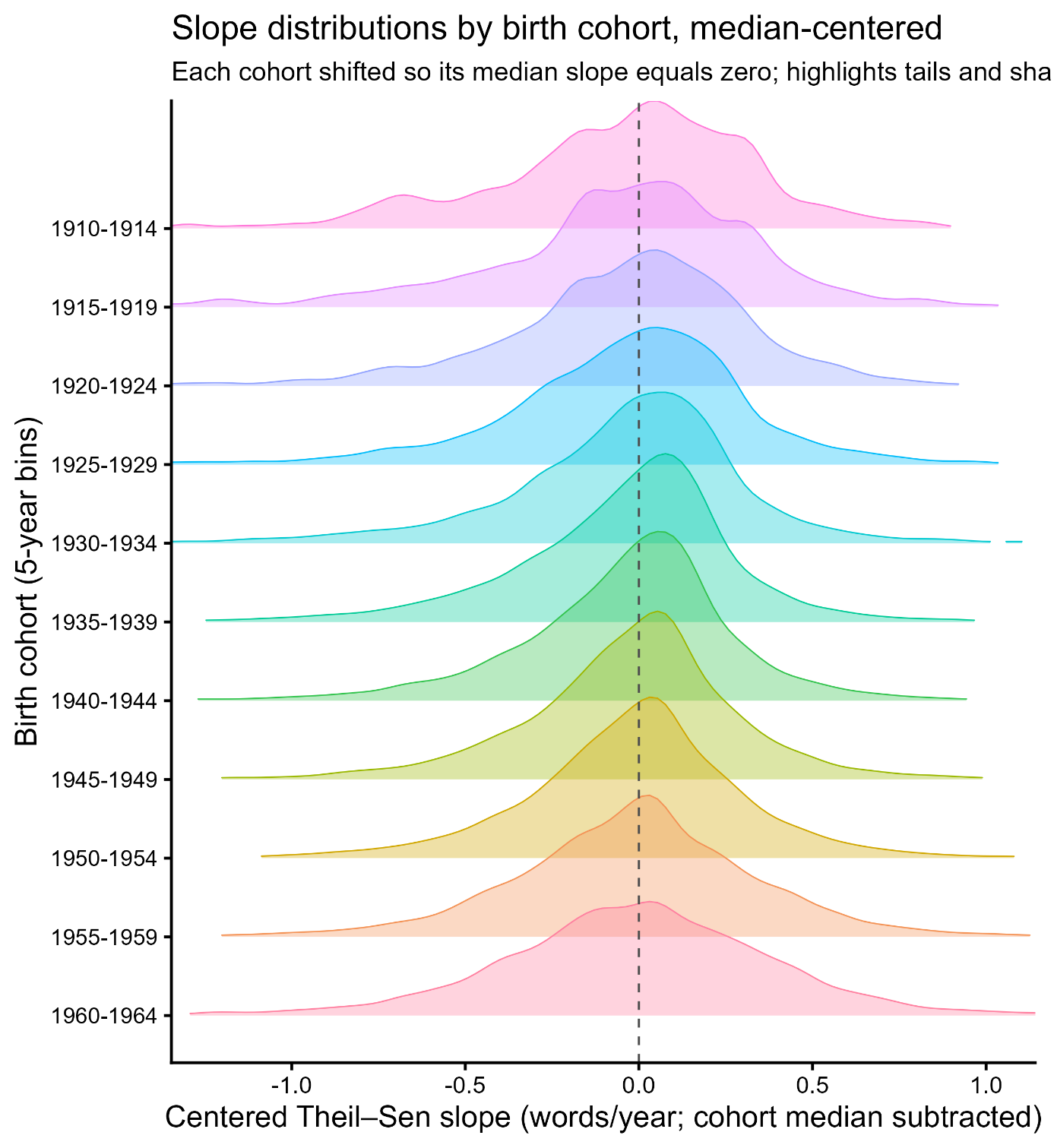

***Figure S7.*** *Median-centered distributions of individual memory-change slopes by birth cohort. Kernel density ridgelines of person-level Theil–Sen slopes (words/year) shown separately for 5-year birth cohorts (cohorts displayed only when n ≥ 200). To reduce age–cohort confounding from diagonal coverage, we restricted to mid-age 65–85 years and summarized slopes within 2-year mid-age bins (age_bin). We retained cohort-by-age cells with at least 50 participants; cohorts were 5-year birth bins. For each cohort, slopes were centered by subtracting the cohort median (vertical dashed line at 0), highlighting differences in distributional shape independent of location shifts. After median-centering, cohorts show broadly similar distribution shapes, with extreme negative slopes present in all cohorts and no strong evidence that cohort differences are driven primarily by a markedly heavier far-left tail in the oldest cohorts.*

**Section 8: Sensitivity analyses**

≥ 2 waves

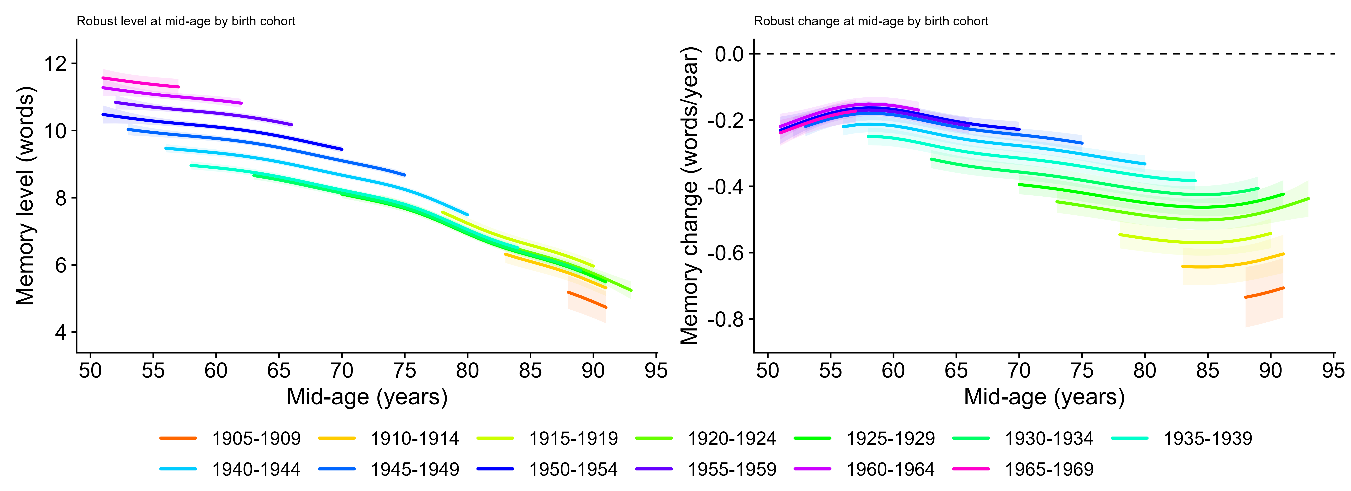

≥ 3 waves

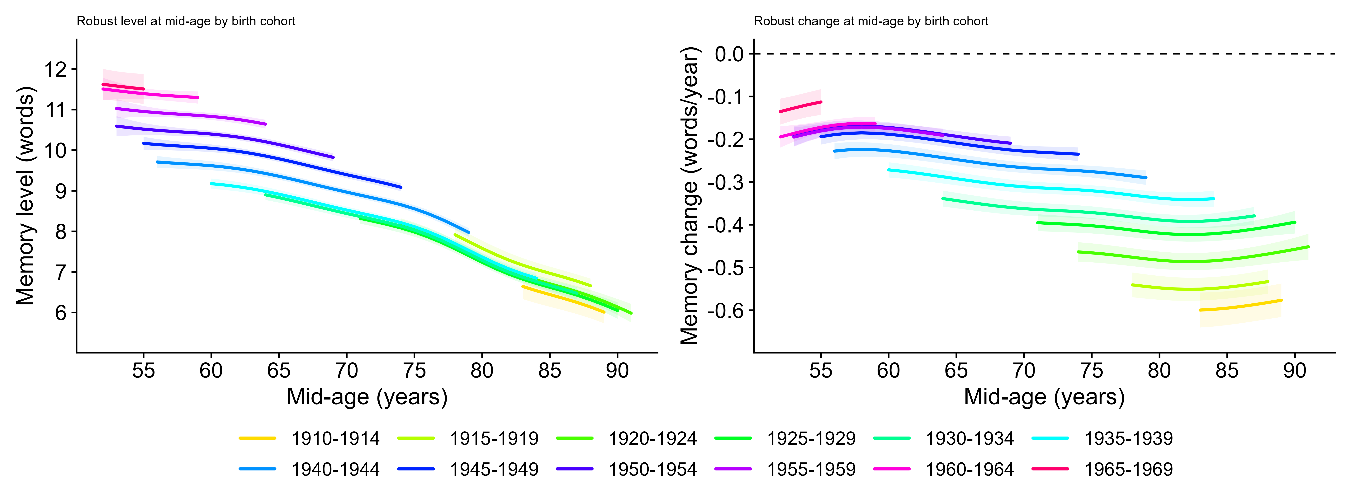

≥ 4 waves

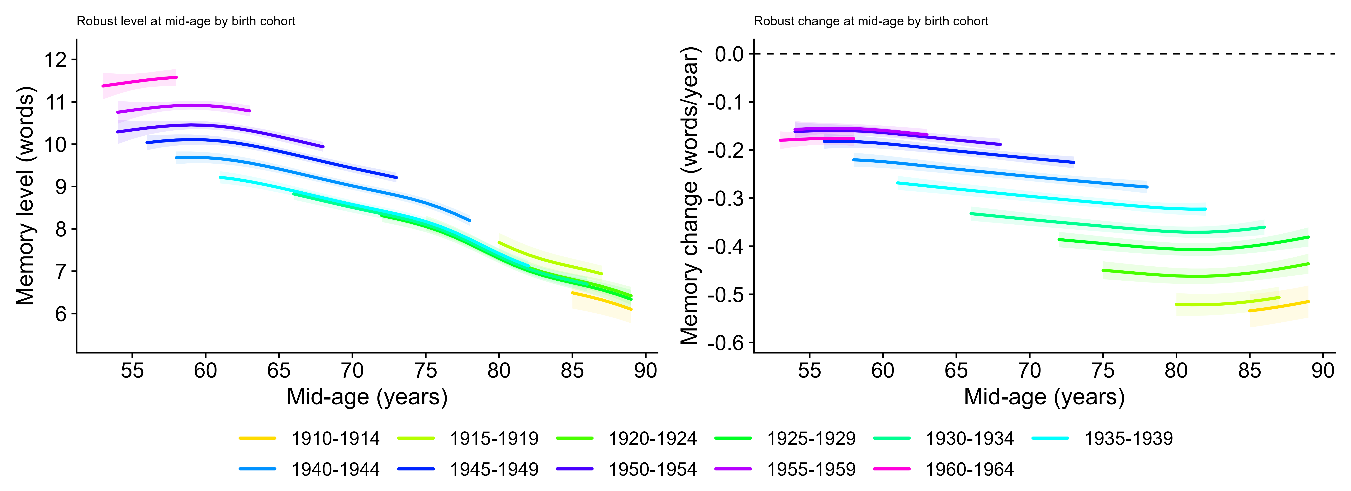

***Figure S8.*** *Robust cohort profiles in memory level and change under different inclusion thresholds.* *We re-estimated the Fig. 2 models after requiring ≥2 (row 1), ≥3 (row 2), or ≥4 (row 3) observations per participant (not including LASA). Left: robust mid-age memory level; right: robust mid-age memory change (Theil–Sen slope, words/year). Across rows, relaxing the inclusion threshold increases the spread of estimated slopes (as expected with fewer observations), while the qualitative cohort ordering is preserved.*

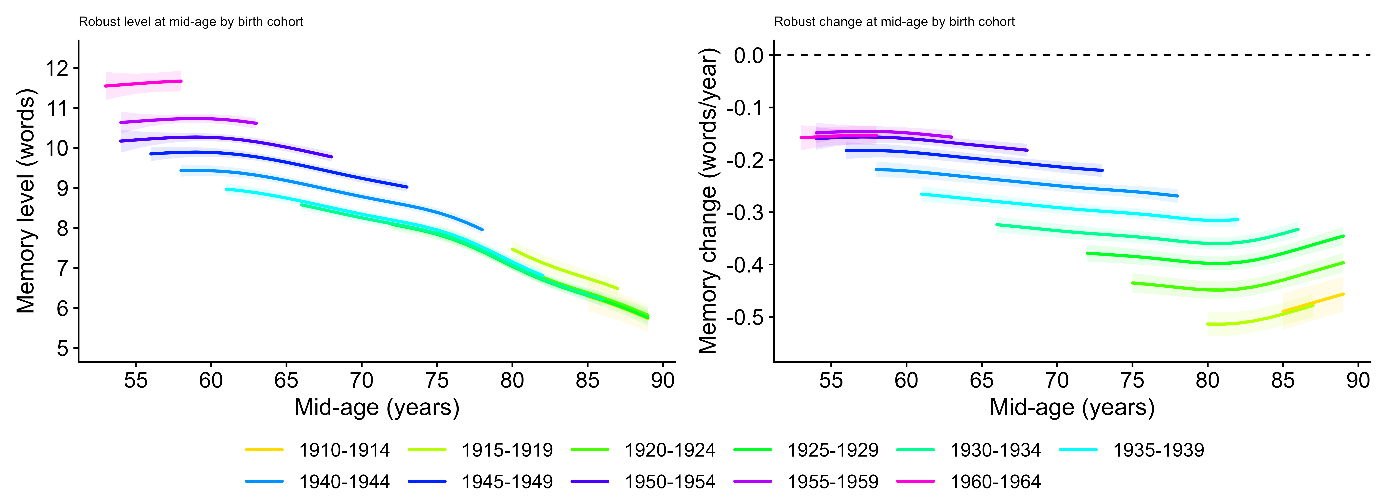

***Figure S9.*** *Sensitivity to selective inclusion in the ≥4-wave sample (inverse-probability weighting).* *Figure 2 re-estimated in the ≥4-wave analytic sample using inverse-probability weights for inclusion (ge4=1) (not including LASA). Inclusion probabilities were modeled by logistic regression as a function of baseline age, baseline episodic-memory performance, baseline interview year, sex, study, and 5-year birth-cohort bin (natural cubic splines for continuous predictors). The resulting weights (inverse predicted inclusion probability; trimmed) were applied to the level and slope GAMs. Reweighted cohort trajectories closely matched the unweighted ≥4-wave results, indicating that the cohort ordering in memory change is not driven by selective inclusion into the ≥4-wave sample under observed baseline covariates.*

**Section 9: Calibration of the generational memory advantage against clinical benchmarks**

To assess the clinical magnitude of the generational memory advantage, we anchored the bounding analysis results against diagnostic memory benchmarks from the Alzheimer's Disease Neuroimaging Initiative (ADNI).

ADNI clinical benchmarks. We used a subsample of 813 ADNI participants (229 cognitively normal [CN], 389 mild cognitive impairment [MCI], 195 Alzheimer's disease [AD]; Table SX) who completed a verbal recall task rescaled to the 0–20 metric used across the five longitudinal studies. Memory scores differed substantially across diagnostic groups (Table SX): CN mean = 14.2 (SD = 2.4), MCI mean = 10.8 (SD = 2.8), AD mean = 7.8 (SD = 3.0). Logistic regression with memory score as the sole predictor yielded strong discrimination for both CN vs. MCI-or-AD (AUC = 0.821) and CN-or-MCI vs. AD (AUC = 0.959), confirming that the recall measure captures clinically meaningful variance across the full spectrum from normal aging to dementia (Table SX). The clinical gap between CN and AD was 6.4 words (14.2 − 7.8); the gap between CN and the pooled MCI+AD group, the point at which clinical ascertainment begins, was 4.4 words (14.2 − 9.8, where 9.8 is the weighted mean of MCI and AD groups).

Cohort advantage from the bounding analysis. The APC bounding analysis estimated a total 20-year cohort advantage at age 80 of 5.0 words under the primary period specification (k=6 for both level and slope) and 4.0 words under the most conservative valid specification, with level and slope contributing approximately 47% and 53% respectively under the primary specification.

Gap-bridging calculation. The 4.5–5.0 word advantage bridges 62–78% of the 6.4-word CN–AD gap, and equals or exceeds the full 4.4-word CN vs. MCI+AD gap, the clinically relevant threshold at which dementia risk becomes elevated and ascertainment begins. These calculations indicate that the generational memory improvement is of sufficient magnitude to account for the reported decline in dementia incidence of approximately 13% per decade documented by the Alzheimer Cohorts Consortium. The gap-bridging fractions should be interpreted as an order-of-magnitude calibration rather than a precise mechanistic estimate: they establish that the generational advantage is large enough to matter clinically, not that memory change is the sole driver of incidence trends.

| Diagnosis | n | Age (mean) | Age (SD) |
| --- | --- | --- | --- |
| CN | 229 | 75.89 | 5.1 |
| MCI | 389 | 74.89 | 7.4 |
| AD | 195 | 75.4 | 7.4 |

***Table S2****. Sample descriptives for the ADNI subsample. CN = cognitively normal; MCI = mild cognitive impairment; AD = Alzheimer's disease.*

| **Diagnosis** | **Mean (native)** | **SD (native)** | **Mean (0–20 scale)** | **SD (0–20 scale)** |
| --- | --- | --- | --- | --- |
| CN | 7.1 | 1.2 | 14.2 | 2.4 |
| MCI | 5.4 | 1.4 | 10.8 | 2.8 |
| AD | 3.9 | 1.5 | 7.8 | 3.0 |

***Table S3****. Memory scores in the ADNI subsample. Memory scores in the ADNI subsample on the native ADNI scale and rescaled to the 0–20 metric used in the main analyses.*

| Comparison | Odds ratio | 95% CI | Accuracy | AUC |
| --- | --- | --- | --- | --- |
| CN vs. MCI or AD | 3.73 | 3.06–4.53 | 0.752 | 0.821 |
| CN or MCI vs. AD | 10.14 | 6.45–15.95 | 0.894 | 0.959 |

***Table S4.*** *Diagnostic accuracy of episodic memory for clinical group discrimination in ADNI. Odds ratios reflect the increase in odds of the more impaired group per unit decrease in episodic memory score. Accuracy and AUC are from logistic regression with episodic memory as the sole predictor. These estimates are used in the main text to calibrate the clinical magnitude of the generational memory advantage.*

**Section 10: Quantile regression of birth cohort effects across the slope distribution**

The main analyses establish that later-born cohorts have higher mean memory levels and slower mean rates of decline. A natural follow-up question is whether the cohort improvement is distributed uniformly across the full slope distribution, reflecting a broad shift of all trajectories, or is concentrated in specific parts of the distribution, particularly the left tail of rapid decliners. We addressed this using quantile regression, which estimates the association between a predictor and the outcome at any specified quantile of the conditional outcome distribution, rather than at the conditional mean as in ordinary least squares regression.

Model specification**.** We fitted the following quantile regression model at each of five quantiles τ ∈ {0.10, 0.25, 0.50, 0.75, 0.90}:

*Q*_τ(slope) = α_τ + β_τ · birth_decade + γ_τ · age_mid_c + Σ δ_τk · study_k

where *Q*_τ(slope) denotes the τ-th conditional quantile of the person-level Theil–Sen slope; birth_decade is birth year expressed in decades and centred at 1950 (so that the coefficient β_τ represents the cohort effect per 10-year birth cohort gap); age_mid_c is the mid-age of each person's observation window centred at 70 years; and study_k are indicator variables for each study (with one study as reference), included to absorb between-study differences in mean slope. The model was fitted using the rq function from the quantreg package in R, with the same inverse-probability-of-follow-up weights used in the main slope analyses.

Age adjustment**.** The inclusion of age_mid_c as a covariate means that the birth cohort coefficient β_τ is estimated holding age at observation constant, it reflects the cohort difference at the same age, not the raw association between birth year and slope which is confounded by the fact that earlier-born cohorts are observed at older ages on average. The age term enters linearly; the heatmap analyses (Figure 5, middle panel) suggest that the age effect on severe decline accelerates at older ages, so the linear approximation may not fully absorb age-related variation at extreme quantiles. However, the bias this would introduce would need to be implausibly large and systematically structured to produce the observed 2:1 ratio of the 10th to 50th percentile cohort coefficient as an artefact, and we regard the linear age adjustment as adequate for the purpose of characterising the distributional shape of the cohort effect.

Standard errors and inference**.** Standard errors were obtained by bootstrap resampling of (x, y) pairs (bsmethod = "xy", R = 500 resamples), which is appropriate for heteroscedastic data and does not assume a specific error distribution. This approach resamples entire observations rather than residuals, making it robust to the correlation structure introduced by study membership. Ninety-five percent confidence intervals were constructed as estimate ± 1.96 × bootstrap SE. All five quantile coefficients were statistically significant at p < 0.001.

Interpretation of results**.** The birth cohort coefficient at each quantile represents the expected difference in Theil–Sen slope between two individuals observed at the same age and from the same study, but born 10 years apart. A positive coefficient (less negative slope) indicates that the later-born individual is expected to have a less steep decline trajectory at that quantile. The pattern of coefficients across quantiles, decreasing from the 10th to the 75th percentile, with a modest uptick at the 90^th^, indicates that the cohort improvement is disproportionately large among those with the most negative slopes (fastest decliners), while individuals near the flat end of the distribution show a smaller but still positive cohort advantage. This pattern is distinct from a location shift, in which all quantile coefficients would be equal, and indicates that generational progress has selectively reduced the prevalence of the worst decline trajectories rather than shifting all trajectories by a uniform amount.

**Section 11: Bout analyses of sustained stability and steep decline**

*Overview and motivation*

The population-level slope distributions and quantile regression analyses (Sections 3, 11) characterise between-person variation in overall decline rates but cannot reveal how change is organised within individual trajectories. To address this, we asked whether the generational improvements documented in Sections 3–5 are expressed not only as a distributional shift in overall slopes but as a difference in the prevalence of sustained trajectory states at equivalent ages — specifically, whether later-born cohorts spend more time in extended plateaus of cognitive stability and less time in episodes of steep accelerated loss. We operationalised this using a windowed bout classification applied to each individual's longitudinal practice-corrected recall record.

*Window construction*

For each participant with at least W consecutive assessments, we extracted every possible consecutive window of length W from the ordered (by age) sequence of observations. Within each window of W assessments at ages a_1 < a_2 < ... < a_W with practice-corrected scores y_1, ..., y_W, we computed:

(i) A Theil–Sen slope (slope_ts), defined as the median of all pairwise slopes (y_k − y_j) / (a_k − a_j) for j < k, as described in Section 3. This is the same robust slope estimator used for person-level change throughout the main analyses.

(ii) A jackknife uncertainty estimate (sd_jk), defined as the standard deviation of leave-one-observation-out Theil–Sen slopes across the W observations, quantifying how sensitive the window slope is to the removal of any single assessment.

(iii) A probabilistic criterion (p_ok), computed as the normal-tail probability that the true slope satisfies the bout criterion, using slope_ts as the mean and sd_jk as the standard deviation:

* For stable bouts: p_ok = P(slope ≥ slope_cut) = 1 − Φ((slope_cut − slope_ts) / sd_jk)

* For steep-decline bouts: p_ok = P(slope ≤ slope_cut) = Φ((slope_cut − slope_ts) / sd_jk)

where Φ is the standard normal CDF. When sd_jk was not finite or was zero (fewer than 4 observations or perfectly collinear), the criterion defaulted to a hard threshold on slope_ts alone.

(iv) A binary flag indicating whether the window met the bout criterion: slope_ts satisfying the directional inequality and p_ok ≥ 0.50 (P_CUT). The 0.50 threshold was chosen as the median-probability criterion: a window is classified as a qualifying bout only if the point estimate and the balance of probabilistic evidence both support the classification.

Additional window-level covariates were recorded for use as nuisance adjustments in the prevalence model: the age at the midpoint of the window (age_mid), the span of ages within the window (win_span_years), and the mean interval between consecutive assessments (mean_interval).

*Person × age-bin collapse*

For each participant and each 1-year mid-age bin (age_mid_bin = floor(age_mid)), the binary outcome was defined as 1 if the participant had at least one qualifying window whose mid-age fell in that bin, and 0 otherwise. This gives one row per person per age bin, with a binary outcome indicating whether a qualifying bout was observed at that age. Participants contributed to multiple age bins if their follow-up span covered multiple bins, and the same window could contribute to the bin corresponding to its mid-age only.

*Prevalence model*

To estimate the cohort-specific probability of a qualifying bout as a function of age, we fitted a binomial generalised additive model (GAM) to the person × age-bin table:

logit(P(outcome = 1)) ~ by_label + study + s(age_mid, k=8) + s(span_total, k=6) + s(win_span_med, k=6) + s(mean_int_med, k=6)

where by_label is a factor for the 5-year birth cohort bin (with all cohorts as levels, estimating a separate intercept per cohort after the shared age smooth), study is a fixed factor absorbing between-study differences, s(age_mid, k=8) is a penalised regression spline of the window mid-age capturing the shared nonlinear age effect on bout prevalence, s(span_total, k=6) adjusts for each person's total follow-up span (longer-followed participants have more windows and a higher chance of qualifying), s(win_span_med, k=6) adjusts for the median window span in years (windows spanning longer age intervals may have higher or lower slope estimates due to the follow-up structure), and s(mean_int_med, k=6) adjusts for the median interval between assessments within the person's windows. Spline basis dimensions were reduced adaptively where the number of unique covariate values was insufficient (k_safe function: k = max(3, min(k_desired, n_unique − 1))). All GAMs were fitted using bam() in the R package mgcv with fREML estimation and discrete = TRUE for computational efficiency.

Predictions were obtained on the probability scale (type = "response") across a fine age grid (0.25-year steps) for each cohort, holding nuisance covariates at their sample medians and fixing study to the first level (alphabetically). Ninety-five percent confidence intervals were derived from the fitted linear predictor and its standard error, back-transformed via the logistic function. Predicted curves were masked in cohort × age-bin cells where fewer than 50 individuals contributed observations, to avoid displaying estimates from sparse regions.

*Stable bout analyses*

A window of W consecutive assessments was classified as a stable bout if:

slope_ts ≥ slope_cut AND p_ok = P(slope ≥ slope_cut) ≥ 0.50

The primary stability threshold was slope_cut = −0.05 words/year, representing near-flat or improving trajectories. Two sensitivity analyses were conducted:

Trail 1 (bout length sensitivity): W varied from 4 to 8 consecutive assessments at fixed slope_cut = −0.05 words/year. Increasing W requires longer uninterrupted stability, reducing overall prevalence but preserving the cohort gradient if the effect is genuine.

Trail 2 (threshold sensitivity): W fixed at 4, slope_cut varied from −0.05 to −0.50 words/year in steps of −0.05. More negative thresholds classify a broader range of declining trajectories as "stable" (lenient criterion), raising overall prevalence substantially. The figure (right column) shows three selected thresholds (−0.05, −0.25, −0.50 words/year) to span the range from strict to lenient.

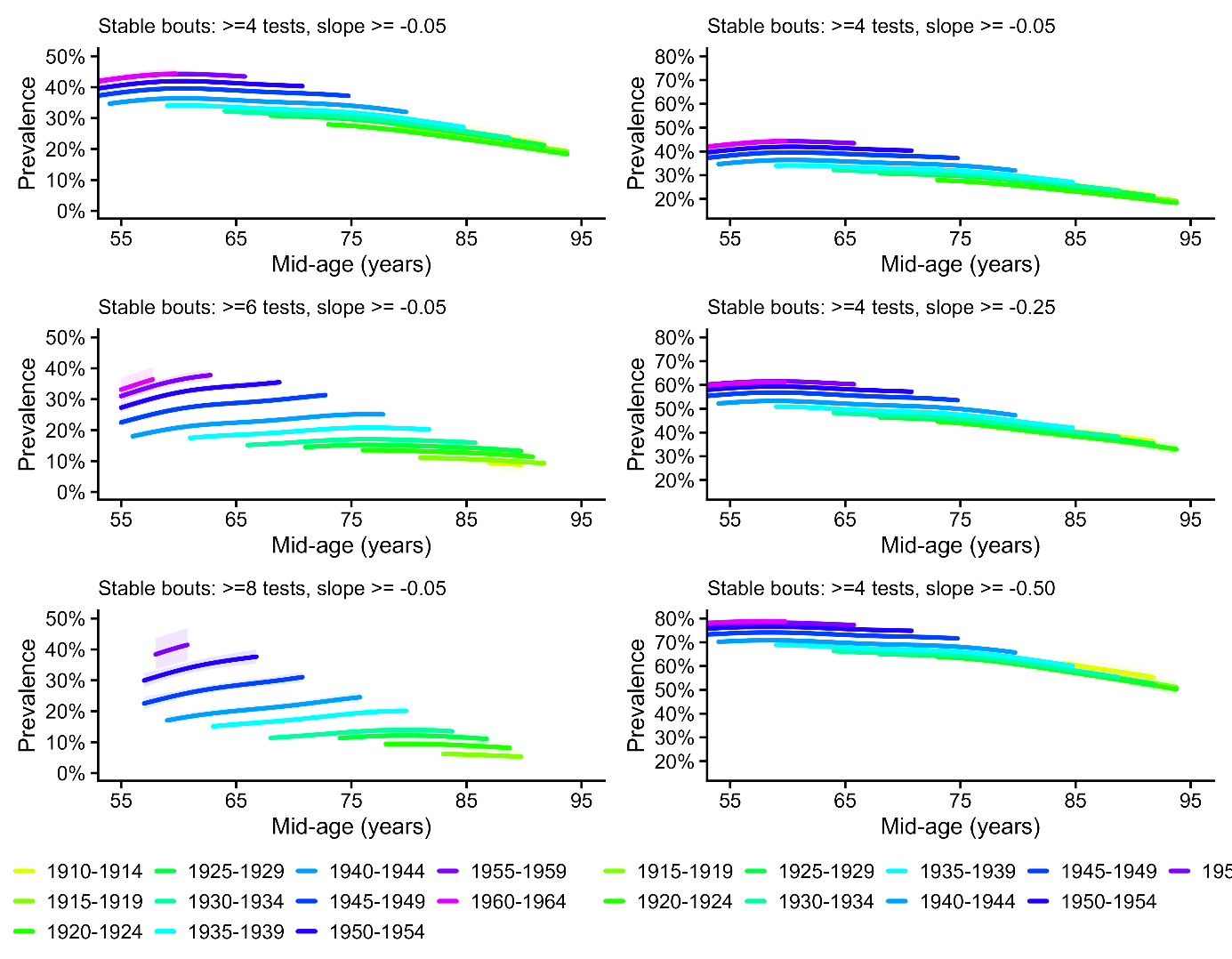

***Figure S10****. Prevalence of sustained stable memory bouts across age and birth cohorts.*

*For each cohort-by-age bin, prevalence is the proportion of participants with at least one qualifying stable window, estimated from a binomial GAM with a shared smooth of age and adjustment for follow-up structure (total follow-up span and within-window spacing). Curves are shown only where the cohort-by-age bin includes ≥50 participants. Left column: sensitivity to bout length (W=4,6,8) at a fixed stability threshold (slope ≥ −0.05 words/year). Right column: sensitivity to stability threshold (slope ≥ −0.05, −0.25, −0.50) at a fixed bout length (W=4).*

*Steep-decline bout analyses*

A window of W consecutive assessments was classified as a steep-decline bout if:

slope_ts ≤ −1.0 words/year AND p_ok = P(slope ≤ −1.0) ≥ 0.50

W was varied from 4 to 6 consecutive assessments at the fixed threshold of −1.0 words/year (Figure 7, left column). The −1.0 threshold corresponds to the "severe decline" category used in the heatmap analyses (Figure 5, middle panel) and is approximately five times the median rate of decline in the sample, selecting only individuals in the extreme left tail of the slope distribution.

*Overall slope threshold classification:* As a complementary approach that does not depend on the windowing procedure, we also classified participants using their full-follow-up Theil–Sen slope (computed across all ≥4 observations, as in Section 3). A participant was classified as a steep decliner at threshold thr if:

slope_ts ≤ thr AND P(slope ≤ thr) ≥ 0.50

where the probability was again computed under a normal approximation using slope_ts and sd_jk. Three thresholds were applied: thr ∈ {−0.75, −1.0, −1.15} words/year. The resulting binary classification (one value per person) was modelled using the same binomial GAM framework, with the person's overall mid-age (midpoint of observed age range) as the age variable and total follow-up span substituted for win_span_med. This approach classifies each person as a decliner or non-decliner based on their entire trajectory rather than any particular window, providing a conceptually distinct operationalisation that avoids the sensitivity of windowed methods to window length.

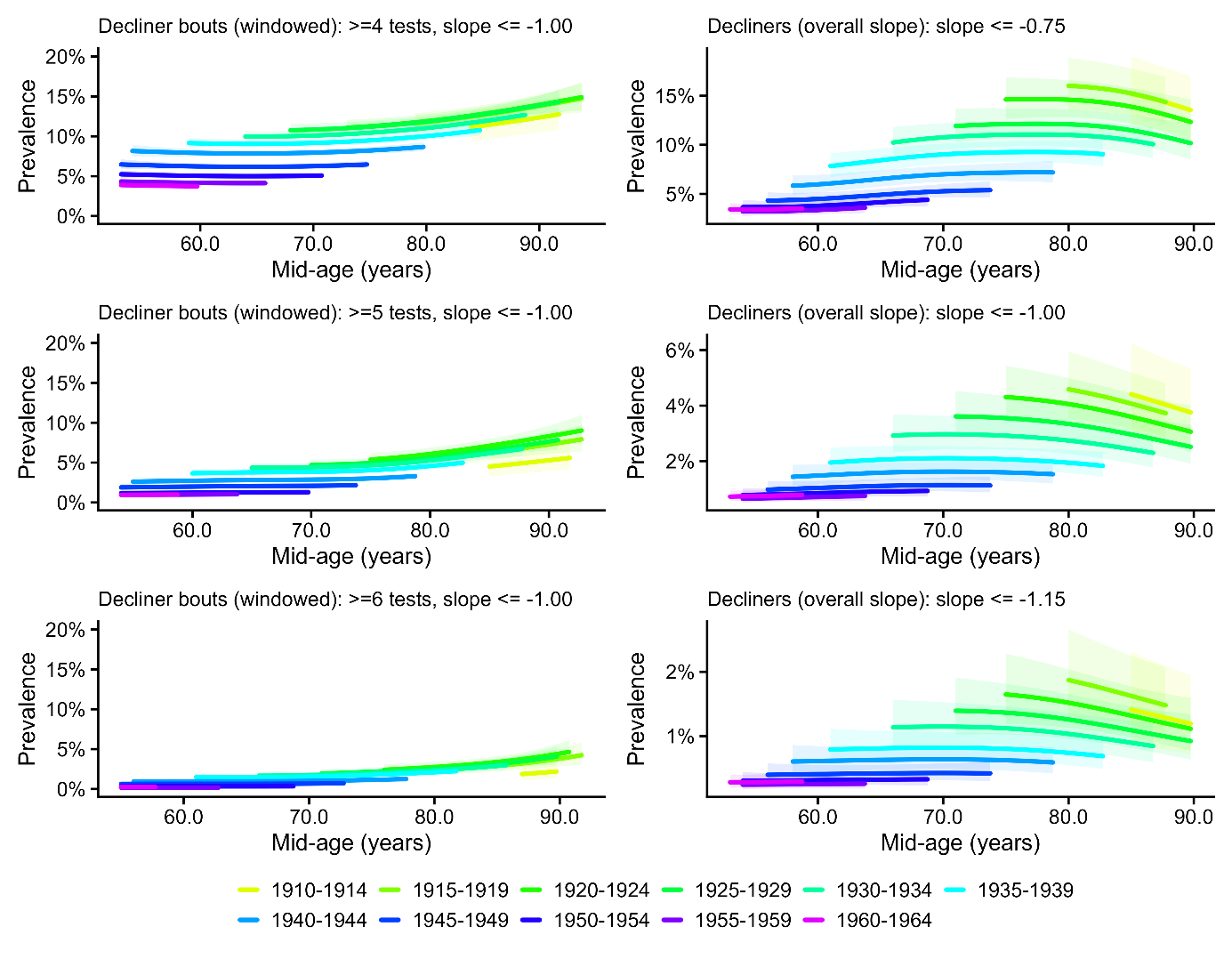

***SI Figure S11****. Prevalence of sustained steep decline in episodic memory across age and birth cohorts*

*Left: participants were classified as having a steep-decline bout if any window of W consecutive assessments (W=4,5,6) met the steep-decline criterion (slope ≤ −1.0 words/year with P(β≤−1.0)≥0.50). Right: participants were classified as decliners based on their full person-level slope (≥4 observations), using increasingly stringent thresholds (≤ −0.75, ≤ −1.0, ≤ −1.15). Modeling and masking rules were identical to Figure S14.*

*Convergence across operationalisations*

The consistency of cohort ordering across the windowed bout approach (varying W and slope_cut) and the overall slope threshold approach (varying thr) in both the stable and steep-decline analyses supports the interpretation that the observed cohort differences reflect a genuine generational shift in trajectory architecture rather than an artefact of any specific analytical choice.

**Section 12: Age–Period–Cohort bounding analysis: full technical details**

This section provides a complete technical account of the APC bounding framework used to estimate cohort effects on memory level (Section 5 of the main text) and memory change (Section 6). The brief description in the main text Methods covers the key logic; the present section documents the mathematical foundations, the derivation of the canonical solution line, the specification and rationale for each constraint, the empirical period-drift estimation procedure, and the bootstrap resampling approach. All analyses were implemented in R using the mgcv package for GAM fitting and base R for the canonical solution line calculations.

### *The APC identification problem*

In observational data, calendar year (period, P) is an exact linear function of birth year (cohort, C) and age (A): P = A + C. This collinearity means that in any regression of an outcome on age, period, and cohort simultaneously, the three linear drift parameters — the rate of change per unit increase in age, period, or cohort, holding the other two constant — cannot be separately identified. Any set of three drift values (bA, bP, bC) that reproduces the observed data can be transformed into an infinite family of equally fitting solutions by adding a constant to bA and bC while subtracting it from bP. Formally, if (bA*, bP*, bC*) fits the data, then so does (bA* + d, bP* − d, bC* + d) for any scalar d. This is the APC identification problem, and it cannot be resolved by collecting more data or fitting more flexible models: it is a mathematical property of the collinearity, not a statistical limitation.

Standard approaches either omit one dimension (introducing bias if that dimension is non-zero), impose an equality constraint (e.g. bA = bC, which is arbitrary), or use instrumental variables that are rarely available in cognitive aging research. We follow Fosse and Winship (2019) in treating the identification problem directly, characterising the full set of equally fitting solutions and then using substantive knowledge to restrict the admissible range.

### *The canonical solution line*

Although the three individual drifts cannot be identified, certain linear combinations of them can. Specifically, a weighted regression of the person-level outcome on age, birth year, and study fixed effects yields two identified composites:

*A* = bA + bP (the age-plus-period composite drift)*

*C* = bC + bP (the cohort-plus-period composite drift)*

These two quantities are identified directly from the data. Every admissible linear decomposition must satisfy bA + bP = A* and bC + bP = C*, so once bP is specified, both bA and bC are determined:

*bA = A* − bP, bC = C* − bP*

The one-dimensional family of triples (bA, bP, bC) that all fit the data equally well is the canonical solution line, parameterised by bP. Rather than selecting a single point on this line — which requires an identifying assumption — the bounding analysis characterises the range of bP values consistent with substantive constraints, yielding a corresponding range of bC values that are all compatible with both the data and the assumed constraints.

For the level analysis, A* and C* were estimated by weighted regression of the person-level robust memory level estimate on centred age, centred birth year, and study fixed effects, with weights proportional to follow-up span and inversely proportional to jackknife estimation variance (matching the weighting used in Figure 1). For the slope analysis, the same regression was applied to person-level Theil–Sen slopes as the outcome, unweighted (matching Figure 1).

### *Sign constraints*

Two weak sign constraints restrict the admissible range of bP without requiring a precise numerical assumption:

Constraint 1: Memory does not improve with age (bA ≤ 0)

Large-scale longitudinal evidence consistently shows that episodic memory declines with advancing age in adulthood, with no credible evidence of net improvement at the population level across the age range studied here (50+). This constraint excludes values of bP for which the implied age drift would be positive, i.e. it excludes bP > A*. For the level analysis, A* = 0.11 words/year; for the slope analysis, A* ≈ 0.00 words/year/year.

Constraint 2: Later-born cohorts are not disadvantaged (bC ≥ 0)

The Flynn effect and the broader secular trends literature provide consistent evidence that later-born cohorts perform at least as well as earlier-born cohorts on cognitive measures in early and mid-adulthood. A negative cohort drift — implying that being born later is associated with lower memory performance at any given age and period — would contradict this evidence and is excluded as implausible. This constraint excludes bP < C* − bP being negative, i.e. it excludes bP > C*. Together, the two constraints restrict bP to the interval [C*, A*] — that is, period drift must lie between the cohort composite and the age composite — defining the light-grey admissible band in Figures S18 and S19.

Empirical estimates of bP from GAM splines of varying flexibility are reported below, after the grid-based analysis that provides the interpretive scale

*Grid-based sensitivity analysis*

The canonical solution line maps every possible linear decomposition (bA, bP,bC) as a function of the assumed period drift bP. Rather than selecting a single point or relying solely on model-based estimates, we characterise the cohort effect across an explicit grid of assumed bP values, expressed as fractions of C*, the tipping point at which bC = 0. This presentation has two advantages. First, the assumptions are stated directly: saying "we assume the period drift

is 25% of C*" is more transparent than saying "we use a GAM with k = 6 to anchor the lower bound." Second, it connects the bounding results to a scale every reader can calibrate: 100% of C* is the worst-case scenario for the cohort interpretation; anything below 50% means the majority of the observed trend is attributable to cohort rather than period.

For memory level, C* = +0.96 words/decade. The admissible grid spans bP from 0% to 100% of C*, with implied cohort effects from 0.96 (at 0%) down to 0.00 (at 100%). For memory slopes, C* = +0.065 words/year/decade. The corresponding grid spans 0.065 (at 0%) to 0.000 (at 100%). Full grids are reported in Tables S6 and S7.

| Assumed period drift | % of C* | bC (words/decade) |
| --- | --- | --- |
| k=6 GAM (empirical) | −24% | 1.19 |
| 0 — period omitted | 0% | 0.96 |
| 25% of tipping point | 25% | 0.72 |
| 50% of tipping point | 50% | 0.48 |
| 75% of tipping point | 75% | 0.24 |
| 100% — tipping point | 100% | 0.00 |

***Table S6****. Cohort effect on memory level under a grid of assumed period drifts. bC is the implied cohort drift in words recalled per decade of later birth, at any given age and period. The tipping-point period drift (C* = +0.96 words/decade) would require a sustained calendar-year improvement in memory performance of approximately one word per decade, comparable in magnitude to the cohort effect itself and contradicted by the empirical spline estimates.*

| Assumed period drift | % of C* | bC (words/yr/decade) |
| --- | --- | --- |
| k=6 GAM (empirical) | −2% | 0.066 |
| 0 — period omitted | 0% | 0.065 |
| k=12 GAM (empirical) | +20% | 0.052 |
| 25% of tipping point | 25% | 0.049 |
| 50% of tipping point | 50% | 0.033 |
| 75% of tipping point | 75% | 0.016 |
| 100% — tipping point | 100% | 0.000 |

***Table S7****. Cohort effect on memory slope under a grid of assumed period drifts. bC is the implied cohort drift in words/year per decade of later birth. The tipping-point period drift (C* = +0.065 words/year/decade) would represent a secular improvement in memory decline rates equivalent to ~33% of the typical annual rate of decline in this sample (~0.20 words/year).*

*Empirical period-drift estimation*

Within the constraint-defined admissible range, we additionally estimated the

long-run linear period drift implied by the data when the period component is

modelled flexibly. We fitted GAMs of the form:

*outcome ~ age + birth_year + s(period, bs='cr', k=K) + study*

where s(period, bs='cr', k=K) is a penalised cubic regression spline for the calendar-year component, fitted with mgcv::bam() using fREML estimation with a smoothness penalty on the integrated squared second derivative. The long-run linear drift implied by each fitted spline was extracted by predicting across the observed period range at median covariate values and regressing the predictions on calendar year, yielding a model-implied estimate of bP, denoted bP_k, for each spline flexibility k. Four specifications were considered: k = 3 (very smooth), k = 6 (moderately smooth), k = 12 (flexible), and k = 0 (period omitted, implying bP = 0).

For the level analysis:

- k = 3 implies bA = +0.20 words/decade, violating Constraint 1. Excluded.

- k = 6 implies bP = −0.23 words/decade (−24% of C*). Valid. bC = 1.19 words/decade.

- k = 12 implies bA = +0.88 words/decade, violating Constraint 1. Excluded.

- k = 0 implies bP = 0. Valid. bC = 0.96 words/decade.

Both valid specifications imply bP ≤ 0, meaning calendar-year trends worked at most neutrally and possibly modestly against memory performance over this window. The period-omitted estimate (bC = 0.96) is therefore conservative: the empirical data, under flexible-but-valid modelling, support a slightly larger cohort effect (bC = 1.19 under k = 6). The GAM-implied values correspond to −24% and 0% of C* respectively, far below the tipping point at which the cohort finding would be challenged.

For the slope analysis:

- k = 3 implies bA > 0, violating Constraint 1. Excluded.

- k = 6 implies bP = −0.0013 words/year/decade (−2% of C*). Valid. bC = 0.066.

- k = 12 implies bP = +0.013 words/year/decade (+20% of C*). Valid. bC = 0.052.

- k = 0 implies bP = 0. Valid. bC = 0.065.

The three valid slope specifications cluster tightly between −2% and +20% of C*, spanning a narrow range of cohort effects (0.052–0.066 words/year/decade). This clustering provides empirical evidence that the true period drift is small relative to the cohort composite. The empirically supported band (dark grey in Figures S12 and S13) spans [min(bP_valid) − ε, max(bP_valid) + ε], where ε = 0.00025 is a small numerical pad, and the corresponding cohort drift bounds are bC_lo = C* − bP_hi and bC_hi = C* − bP_lo.

The GAM-implied values serve as empirical calibration points on the grid rather than as identifying assumptions: they indicate where the data place the period component under flexible modelling, but cannot resolve the identification problem. GAM splines with any finite k cannot distinguish a genuine slowly-drifting period effect from a confounded cohort trend. The grid framing is therefore primary; the GAM values are supplementary evidence that the true period drift is likely near the lower end of the grid.

### *Tipping-point analysis*

To assess the robustness of the positive cohort effects, we identified the period drift required to render each cohort advantage exactly null (bC = 0). Setting bC = 0 implies bP = C*, so the tipping-point period drift is C* itself. For memory level, C* = +0.96 words/decade. This would require a sustained secular improvement in memory test performance of approximately one additional word recalled per decade, comparable in magnitude to the cohort effect itself

and in the opposite direction from what the empirical spline estimates imply.

For memory slopes, C* = +0.065 words/year/decade — approximately one-third of the average annual rate of memory decline in this sample (~0.20 words/year). The k = 6 spline implies bP ≈ −0.001 (~2% of C* in the opposite direction); the k = 12 spline implies bP ≈ +0.013 (~20% of C*). Accepting a null cohort slope effect would therefore require a period improvement five times larger than the most generous valid empirical estimate, consistent across decades but undetectable by flexible spline models.

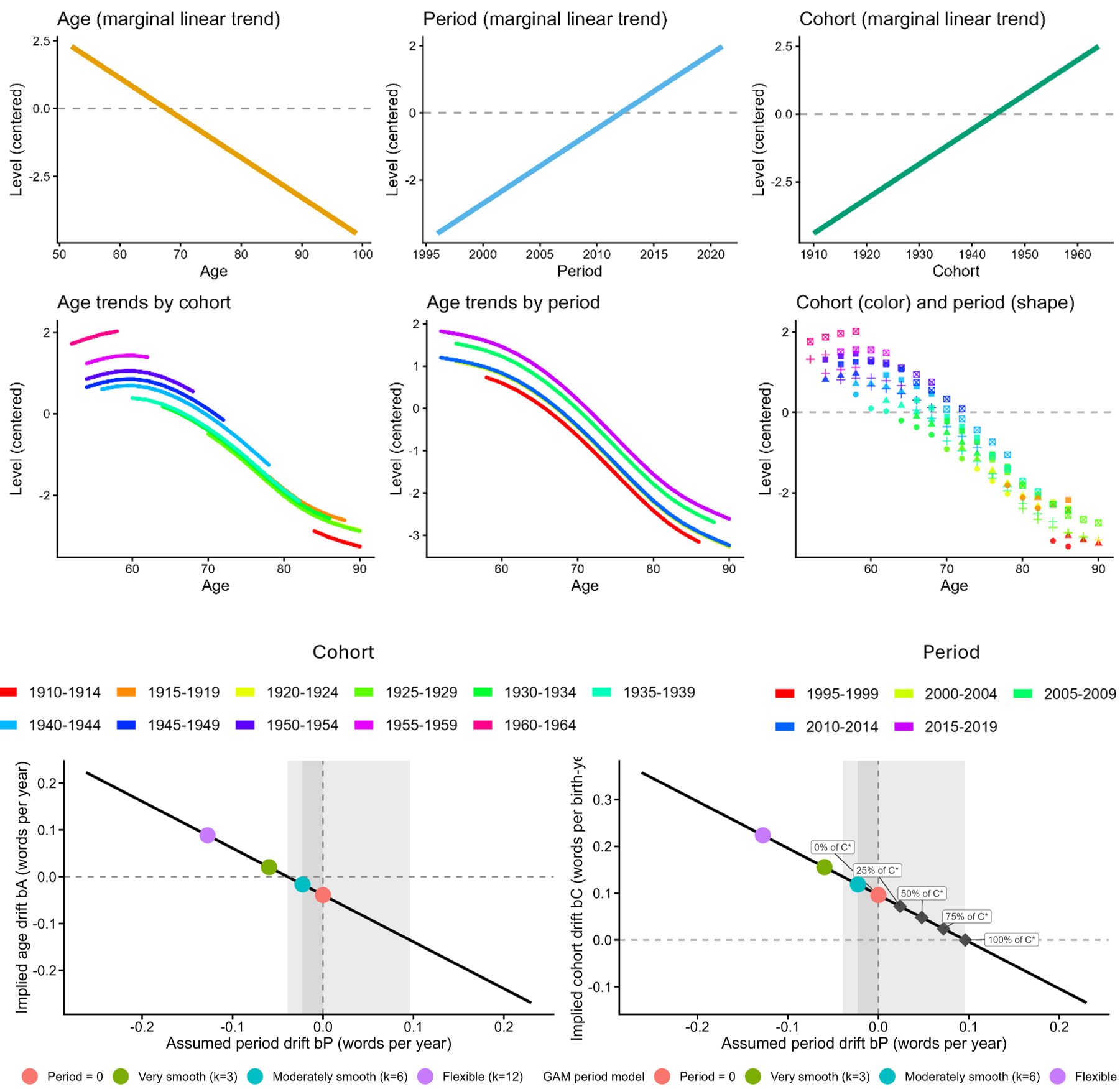

***Figure S12****: Age–period–cohort decomposition for memory level. Top row: marginal linear trends of the memory level outcome over age (left), calendar year (middle), and birth cohort (right), adjusted for study. Middle row: cohort-stratified trajectories across age (left), period-stratified trajectories across age (middle), and binned means by cohort and period (right); data shown only where cohort n ≥ 200 and cohort × age-bin n ≥ 50. Bottom row: canonical solution line showing all linear APC decompositions consistent with the observed data, plotted as implied age drift bA (left) and implied cohort drift bC (right) as a function of assumed period drift bP. The light-grey band marks the admissible range under both sign constraints (memory does not improve with age; later cohorts are not disadvantaged). The dark-grey band spans the two valid period specifications, period omitted (k=0, pink) and moderately smooth (k=6, teal), which imply period drifts at or below zero; specifications k=3 (green) and k=12 (purple) are excluded because both imply positive age drift (bA > 0), visible as both points falling above the bA = 0 dashed line in the left panel. Diamonds show the implied cohort effect at 0%, 25%, 50%, 75%, and 100% of C*, where C* is the period drift that would be required to eliminate the cohort effect entirely (tipping point).*

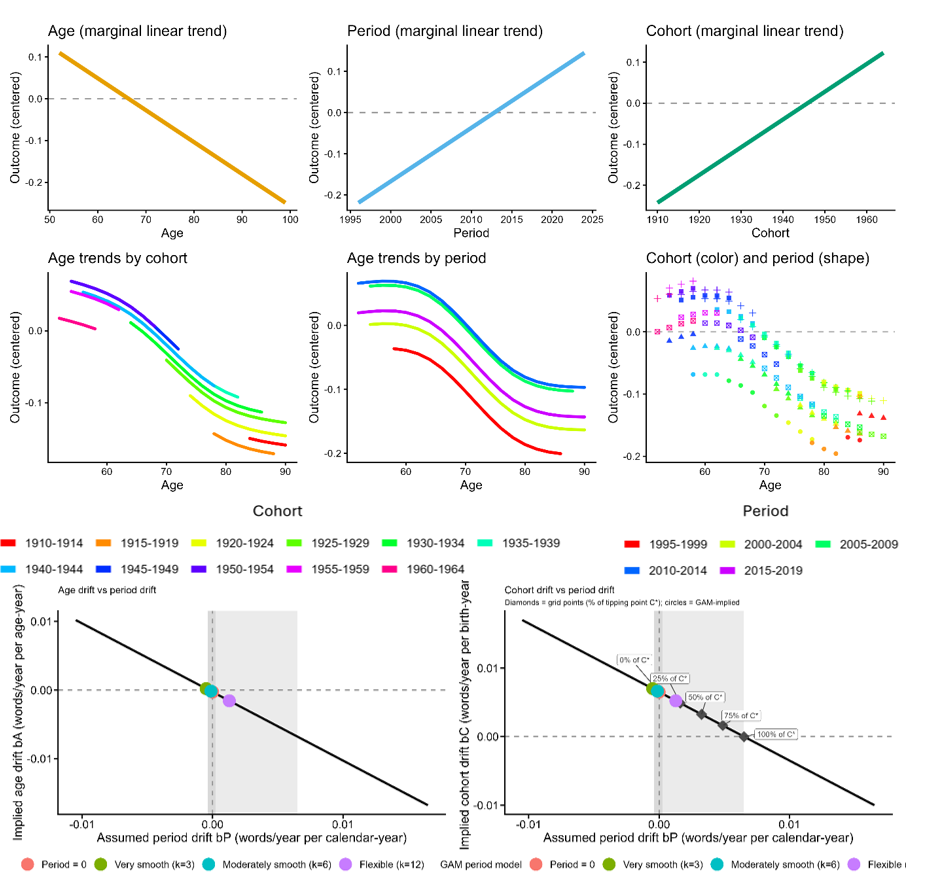

***Figure S13****. Age–period–cohort decomposition for memory slope. Top and middle rows follow the same structure as Figure 6, with person-level Theil–Sen rates of change (words/year) substituted for memory level. Bottom row: canonical solution line for memory slope, plotted as implied age drift bA (left) and implied cohort drift bC (right) as a function of assumed period drift bP. The light-grey band marks the admissible range under both sign constraints (see Figure 6). Unlike the level analysis, the valid specifications for slopes are moderately smooth (k=6, teal) and flexible (k=12, purple), as the period-omitted specification and k=3 are excluded; the dark-grey band spans this range, straddling zero slightly in the left panel, reflecting greater uncertainty in within-person slope estimates than in level estimates. The very smooth k=3 specification is excluded because it implies positive age drift. Diamonds show the implied cohort slope advantage at 0%, 25%, 50%, 75%, and 100% of C, where C* is the tipping point (see Figure 6); both valid specifications (filled circles) fall below 25% of C*, well short of the period drift that would be required to eliminate the cohort finding.*

*Bootstrap stability of level bounds.*

The bounding analysis for memory level retains two valid period specifications satisfying both sign constraints: period omitted (bP = 0, lower bound) and moderately smooth spline k=6 (upper bound). The period-omitted lower bound is defined by assumption rather than estimation; its bootstrap variability reflects only resampling noise in the C_star estimate (SD = 0.08 words/decade) and was positive in all 200 resamples, with a median of 0.96 words/decade (5th–95th percentile: 0.83–1.10). The k=6 upper bound was positive in all 200 resamples, with a median of 1.15 words/decade (5th–95th percentile: 0.99–1.48). One resample produced an extreme upper bound (36.5 words/decade) attributable to k=6 spline overfitting in a bootstrap sample with thin tails in the period distribution; excluding this resample the 95th percentile was 1.44 words/decade. The bootstrap density plot in Figure S5 is trimmed to the central 98% of the distribution for legibility.

The more flexible k=12 specification, which defined the upper bound in an earlier version of this analysis, was excluded because it implies positive age drift (bA = +0.09 words/year) in violation of the sign constraint that memory does not improve with age. Its bootstrap instability — producing implausible bP12 values between +2.6 and +5.3 words/year in five resamples of the original analysis — further supports this exclusion: a specification that is well-identified should not produce numerically unstable estimates under resampling. The conservatively bounded range of 0.96–1.19 words/decade is both analytically justified and empirically stable across all valid bootstrap resamples.

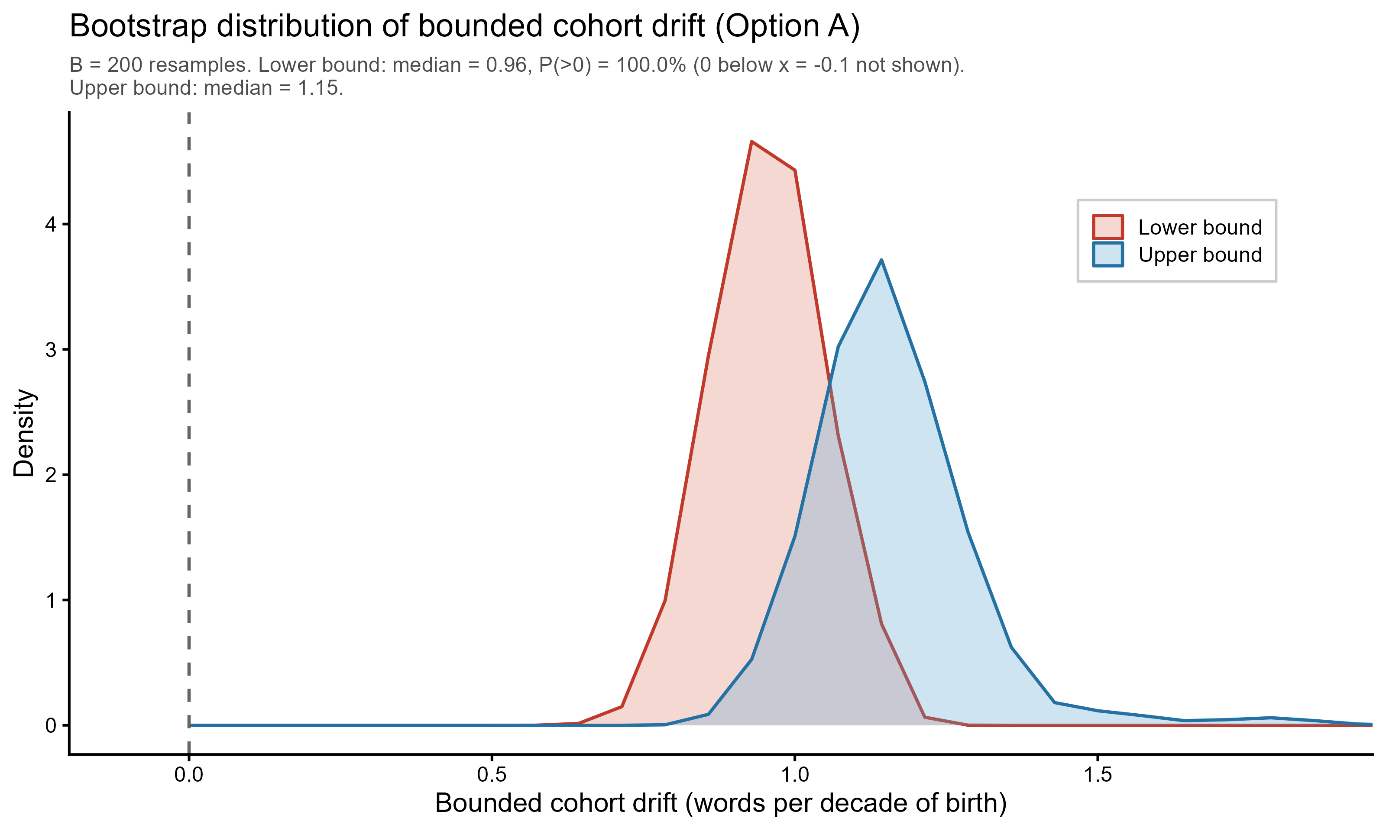

***Figure S14****. Bootstrap distribution of bounded cohort drift in memory level (200 resamples, within-study resampling). Each resample draws participants with replacement separately within each study and re-estimates the full bounding model. The lower bound (red) corresponds to the period-omitted specification (bP = 0), whose variability reflects resampling noise in the cohort coefficient alone. The upper bound (blue) corresponds to the moderately smooth period spline (k = 6). Both distributions are trimmed to the central 98% for legibility; one extreme upper-bound resample (36.5 words/decade, attributable to spline overfitting in a bootstrap sample with thin tails in the period distribution) falls outside the plot window. Dashed vertical line marks zero. Both bounds were positive in all 200 resamples.*

*Bootstrap stability of slope bounds.* The slope bounding analysis retains three valid period specifications: period omitted (bC = 0.065 words/year/decade), moderately smooth k=6 (bC = 0.066), and more flexible k=12 (bC = 0.052). The very smooth k=3 specification was excluded because it implies positive age drift (bA = +0.0002 words/year/age-year), violating the constraint that memory does not improve with age. The three valid specifications cluster tightly between 0.052 and 0.066 words/year/decade, indicating that the slope bounding result is relatively insensitive to period model specification within the valid range. Bootstrap resampling (B=200, within-study) using k=6 as the lower bound and k=12 as the upper bound confirmed that the lower bound was positive in 93.5% of resamples (median 0.052 words/year/decade; 5th–95th percentile: −0.015 to 0.069) and the upper bound positive in 99.5% of resamples (median 0.067; 5th–95th percentile: 0.052 to 0.084). Two resamples produced extreme upper bounds (2.75 and 3.10 words/year/decade) attributable to k=12 overfitting; excluding these, the 95th percentile of the upper bound was 0.084. The bootstrap density plot in SI Figure S6 is trimmed to the central 98% of the distribution for legibility. Unlike the levels analysis, the lower bound for slopes shows modest instability (6.5% negative resamples), reflecting the smaller absolute magnitude of the slope cohort effect relative to measurement noise, though the median and point estimate remain robustly positive.

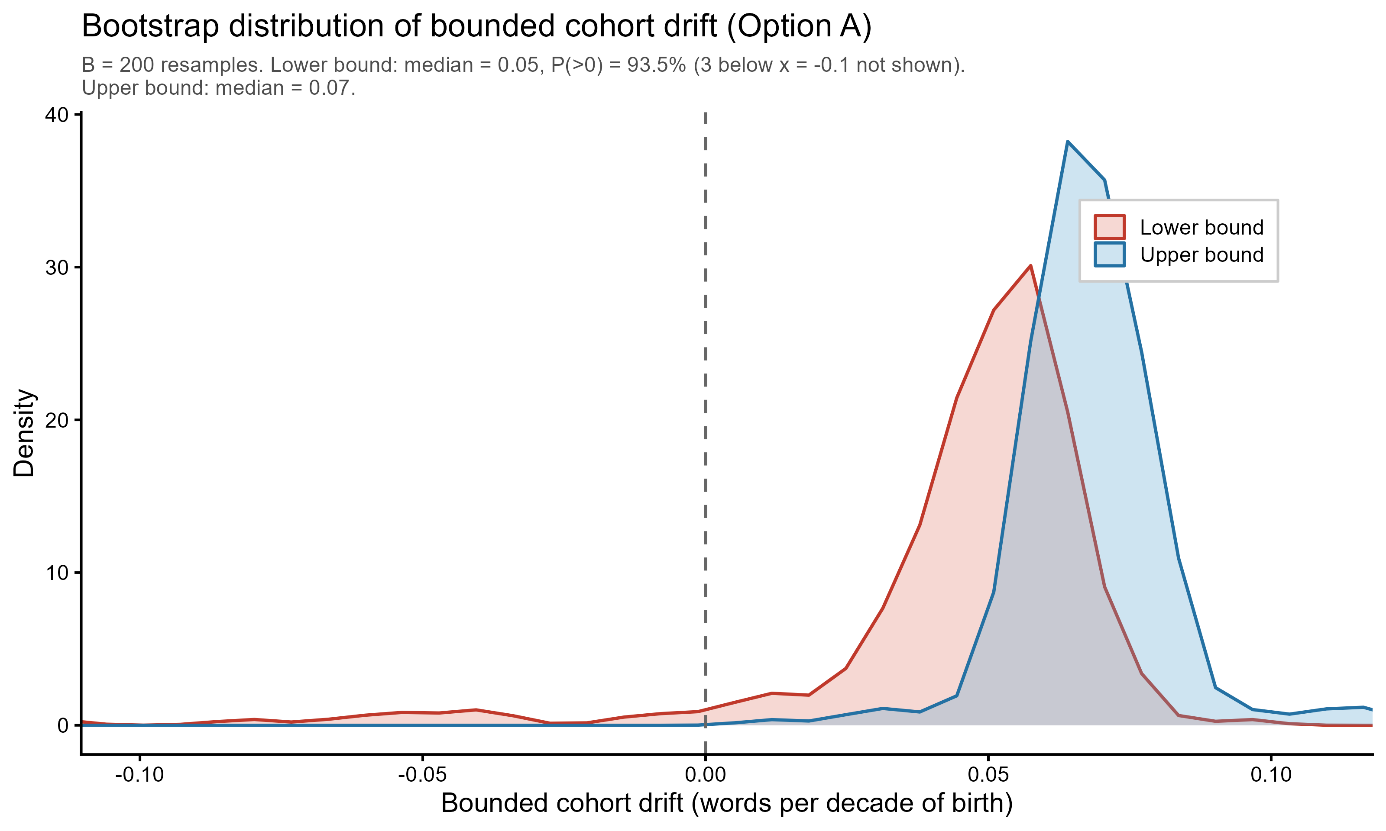

***Figure S15****. Bootstrap distribution of bounded cohort drift in memory slope (200 resamples, within-study resampling). Each resample draws participants with replacement separately within each study and re-estimates the full bounding model. The lower bound (red) corresponds to the moderately smooth period spline (k = 6) and the upper bound (blue) to the more flexible spline (k = 12). Both distributions are trimmed to the central 98% for legibility; two extreme upper-bound resamples (2.75 and 3.10 words/year/decade, attributable to k = 12 overfitting) fall outside the plot window. Dashed vertical line marks zero. The upper bound was positive in 99.5% of resamples; the lower bound in 93.5%, reflecting the smaller absolute magnitude of the slope cohort effect relative to measurement noise.*

### *Decomposition into level and slope components*

To translate the bounding results into a concrete picture of late-life cognitive trajectories (Figure 4), the total 20-year cohort advantage at age 80 was decomposed into two additive components. The level component is the cohort difference in memory score already present at age 60, equal to bC_level × 20 years. The slope component is the additional advantage accumulated through differential decline between ages 60 and 80, equal to bC_slope × 20 years × 20 years of follow-up. Under the primary period specification (k = 6 for both level and slope, the most empirically supported valid specification), these components are approximately 2.37 words (level: 1.187 × 2) and 2.66 words (slope: 0.066 × 2 × 20), summing to 5.03 words. Under the most conservative valid specification (period omitted for level, k = 12 for slope), the components are 1.92 words (level: 0.960 × 2) and 2.09 words (slope: 0.052 × 2 × 20), summing to 4.01 words. The full 20-year advantage therefore ranges from approximately 4.0 to 5.0 words across all valid period specifications, with the level and slope components contributing roughly comparable shares in both scenarios.

The level component is the cohort difference in memory score already present at age 60, equal to bC_level × 20 years (where 20 is the cohort gap in years and bC_level is in words per birth year). The slope component is the additional advantage accumulated through differential decline between ages 60 and 80, equal to bC_slope × 20 years × 20 years of follow-up (where bC_slope is in words per year of age per birth year). The age trajectory shape used to construct Figure 4 Panel A was taken from the growth-curve model described in Section 7, fitted with period omitted, for visualisation purposes only; all numerical estimates derive from the bounding analysis.

**Section 13: Mortality sensitivity analysis for quantile regression**

*Overview and motivation*

The quantile regression analysis (Section 11) revealed that the generational improvement in memory slope is disproportionately concentrated in the left tail of the slope distribution: the cohort effect at the 10th percentile is approximately 1.7 times the size of the median effect. A potential artefactual explanation is terminal decline, the well-documented acceleration of cognitive decline in the months and years preceding death. If earlier-born cohorts contain a higher proportion of participants in terminal decline at any given age (because they are older on average within the overlapping observation window), this could artificially inflate the left tail of earlier cohorts' slope distributions, mimicking a cohort difference in steep decline. We conducted a direct empirical test of this hypothesis.

*Study selection*

The sensitivity analysis was restricted to SHARE and HRS, the two studies for which mortality information is available with near-complete ascertainment. Together these studies contributed 70,859 participants (SHARE: 47,954; HRS: 22,905), representing approximately 88% of the full analytic sample. ELSA, TILDA, and LASA were excluded from this analysis rather than retained with unknown mortality status, which would have diluted the comparison.

*Mortality data sources*

For SHARE, year of death was obtained from the end-of-life interview data (raxyear, harmonised to death_year), which is collected via proxy interview following a respondent's death and covers all waves. For HRS, year of death (RADYEAR) was extracted from the RAND HRS Longitudinal File (Version 2022 V1), which ascertains mortality through the National Death Index, exit interviews, and wave-to-wave tracking. Both sources provide person-level death year that is independent of the cognitive assessments.

*Exclusion criterion*

A participant was excluded from the restricted sample if a death year was recorded and that death occurred within 2 years of their last observation in the slope table. Last observation year was approximated as birth year plus the maximum age at observation (birth_year + age_max). Participants with no recorded death — whether still living at the time of data extraction or with unknown vital status — were retained. The 2-year threshold was chosen to be study-design neutral: it is the minimum interval that is structurally meaningful for HRS, which operates on a biennial interview schedule, and is conservative for SHARE, which has annual follow-up.

The numbers excluded under this criterion were: SHARE 1,197 of 47,954 (2.5%); HRS 3,207 of 22,905 (14.0%). The higher exclusion rate in HRS reflects the older age composition of that sample rather than differences in mortality ascertainment. The restricted analytic sample comprised 66,455 participants.

*Quantile regression specification*

The quantile regression was fitted identically to the main analysis (SI Section 11):

*slope_ts_∼birth_decade+age_mid_c+study*

with quantile levels τ ∈ {0.10, 0.25, 0.50, 0.75, 0.90}, participant-level precision weights, and bootstrap standard errors (R = 500, xy-pair resampling). Both the full SHARE+HRS sample and the mortality-restricted sample were fitted under this specification.

*Results*

Full sample and restricted sample coefficient estimates for the birth_decade term are presented in Table S5 and illustrated in Figure S12. The overall pattern was highly stable across the two samples. All five quantile-specific confidence intervals overlapped between the full and restricted samples, and the U-shaped profile of cohort effects across the distribution, with the largest effects at the 10th and 90th percentiles and the minimum at the 75^th^, was preserved in the restricted sample.

The cohort effect at the 10th percentile attenuated from 0.102 (95% CI: 0.074–0.130) in the full sample to 0.082 (95% CI: 0.054–0.111) in the restricted sample, a reduction of 19.2%. The median effect attenuated from 0.060 (95% CI: 0.046–0.073) to 0.054 (95% CI: 0.041–0.067), a reduction of 9.3%. The 10th-to-50th percentile ratio consequently declined from 1.71 to 1.52, indicating that proximity to death accounts for a modest portion of the left-tail concentration but leaves the majority of it intact. At the upper percentiles, the restricted sample showed slightly larger cohort effects than the full sample (+27.4% at the 75th percentile, +24.3% at the 90th percentile), consistent with the removal of imminent decedents producing a modest upward shift in upper-tail trajectories.

The pattern of changes across quantiles is shown in SI Figure S[X+1]: the delta plot illustrates a systematic gradient from modest negative values at lower percentiles (reflecting partial mediation by terminal decline) to positive values at upper percentiles, but with all changes small in absolute terms relative to the baseline cohort effect. These results indicate that terminal decline artefacts cannot account for the left-tail concentration of the generational improvement in memory trajectories. The finding that later-born cohorts show disproportionately large improvements among those with the steepest decline profiles reflects a genuine cohort-level shift rather than a mortality-selection artefact.

| **τ** | **Full estimate** | **Full 95% CI** | **Restricted estimate** | **Restricted 95% CI** | **Δ (%)** | **CIs overlap** |
| --- | --- | --- | --- | --- | --- | --- |
| 0.10 | 0.102 | [0.074, 0.130] | 0.082 | [0.054, 0.111] | −19.2% | Yes |
| 0.25 | 0.074 | [0.057, 0.092] | 0.064 | [0.044, 0.085] | −13.5% | Yes |
| 0.50 | 0.060 | [0.046, 0.073] | 0.054 | [0.041, 0.067] | −9.3% | Yes |
| 0.75 | 0.031 | [0.016, 0.047] | 0.040 | [0.023, 0.057] | +27.4% | Yes |
| 0.90 | 0.075 | [0.058, 0.092] | 0.093 | [0.078, 0.108] | +24.3% | Yes |

***Table S5****: Quantile regression results: full vs mortality-restricted sample*

Estimates are words/year per decade of birth, age-adjusted, with study fixed effects. Restricted sample excludes SHARE and HRS participants who died within 2 years of their last observation (SHARE: n = 1,197 excluded; HRS: n = 3,207 excluded).

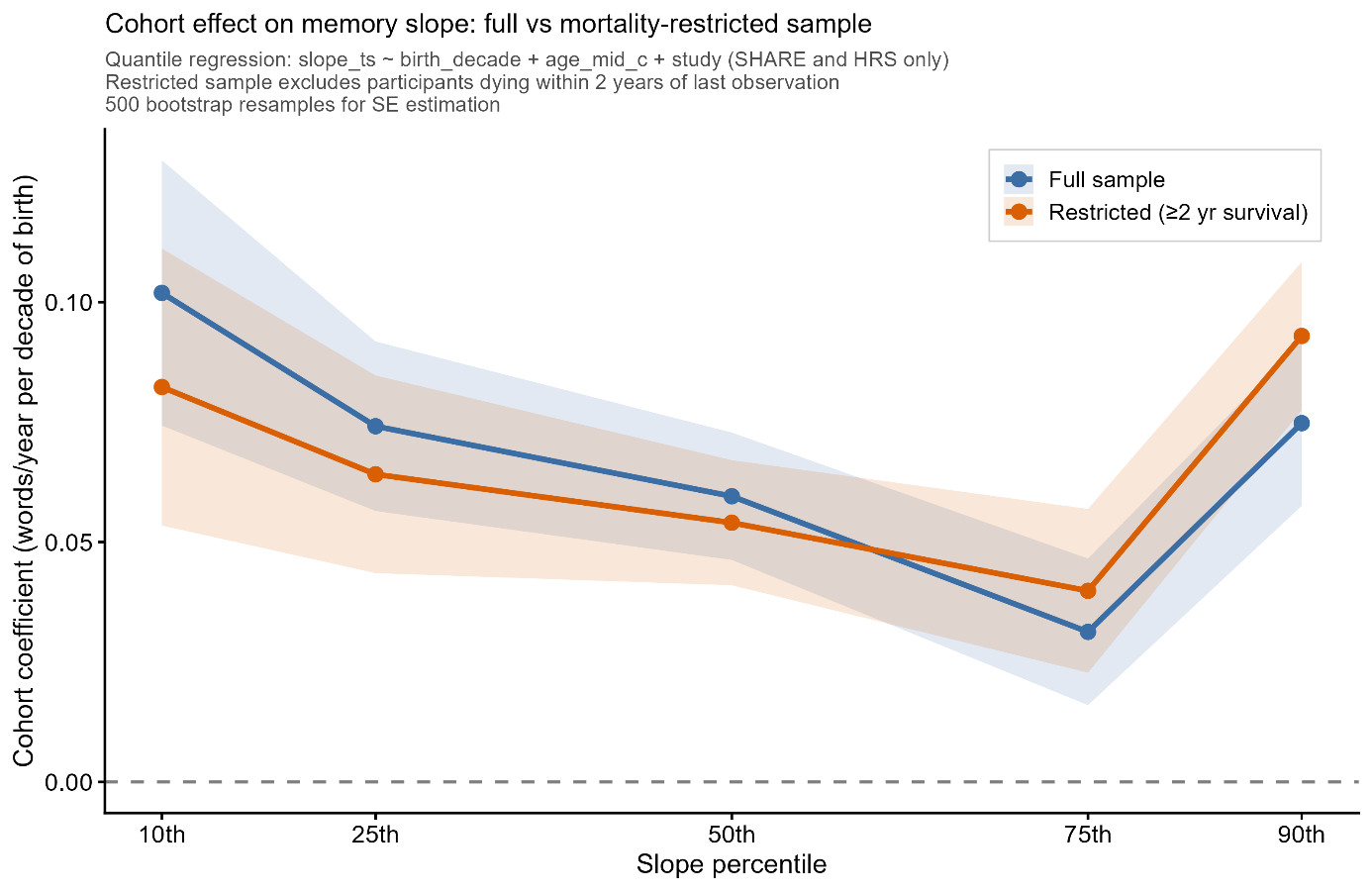

***Figure S16****: Quantile regression cohort coefficients across the slope distribution for the full SHARE+HRS sample (blue) and the mortality-restricted sample (orange), with 95% bootstrap confidence intervals. The two curves follow the same U-shaped profile and have overlapping confidence intervals at all five quantiles, indicating robustness of the distributional pattern to proximity-to-death exclusions.*

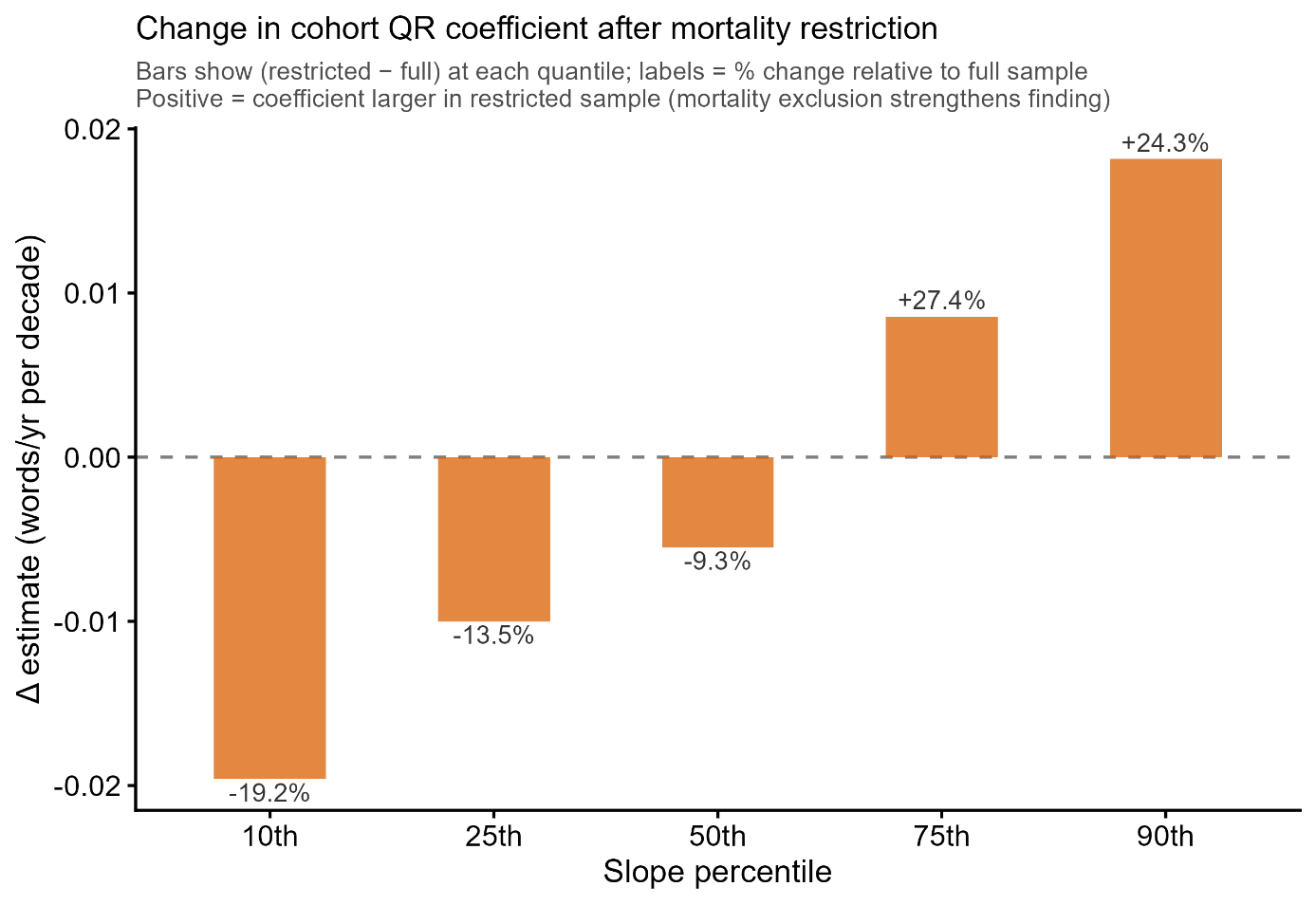

***Figure S17:*** *Change in cohort coefficient (restricted minus full) at each quantile, expressed as absolute difference (bars) and percentage change relative to the full-sample estimate (labels). Negative values at lower quantiles indicate modest attenuation attributable to terminal decline; positive values at upper quantiles indicate slight amplification after removal of imminent decedents.*

**Section 14: Effect size comparisons with previous studies**

Several methodological features help explain both where the current findings converge with previous studies and where they differ or extend prior results. Here we perform some quantitative comparisons with a selection of influential previous studies. Studies using single cohort comparisons separated by roughly 10–25 years, such as Brailean et al. (2018) using LASA (N=1,351; 6-year follow-up) and Gerstorf et al. (2022) using the Berlin Aging Studies (N=811 matched participants), have been well-powered to detect level differences, which are large and consistent across the literature. Gerstorf et al. (2022) estimated a cohort level effect of d=1.20 at age 78 (equivalent to d≈0.48 per decade), and Badham's (2024) meta-analysis across 51 studies yielded a pooled d=0.25 [95% CI 0.17–0.32], increasing approximately 0.02 standard deviations per year of cohort separation (d≈0.20 per decade). The present study is fully consistent with these findings: being born one decade later is associated with approximately 0.96 additional words recalled at any given age under the conservative period-omitted specification, corresponding to d = 0.32 per decade, falling between the two prior estimates as expected given the broader and less selected sample. Under the primary specification (k=6), the level advantage is approximately 1.2 words per decade (d ≈ 0.40), which also falls between the two prior estimates.

The studies diverge regarding conclusions about birth cohort effects on the slope, and the divergence is quantitatively consistent with a power explanation. The cohort slope advantage we estimate, approximately 0.065 words/year per decade of birth, or roughly 33% of the average annual rate of decline, corresponding to d=0.18 per decade, is a small effect by conventional standards, and detecting it reliably requires both a large sample and extended follow-up. This is directly visible in the uncertainty of prior null results: Gerstorf et al. (2022) reported a non-significant trend in the opposite direction (γ11 = −1.41, SE = 1.56), with a 95% confidence interval spanning 6.1 Digit Symbol units, equivalent in standardized terms to d ≈ [−0.62, +0.23], a range that encompasses the effect we observe (d = 0.18 per decade). That the point estimate falls on the opposite side of zero is itself consistent with sampling noise around a small true effect: their minimum detectable effect at 80% power corresponds to approximately 35% of their average annual decline per decade, placing our estimated slope effect just at the lower boundary of what their sample could have reliably detected. Brailean et al. (2018) faced a further constraint: with only three measurement occasions over six years, individual slope estimates carry substantial noise, limiting sensitivity to small trajectory differences regardless of sample size. Consistent with the present findings, a prior study with a larger sample and follow-up up to 17 years, Vonk et al. (2019; N=2,840), did find slope advantages for later-born cohorts in unadjusted models across all racial/ethnic groups, though effects were attenuated and inconsistent across groups after full covariate adjustment. The null slope findings in Gerstorf et al. (2022) and Brailean et al. (2018) should therefore be read as absence of evidence rather than evidence of absence: their confidence intervals are consistent with the effect we report, and the present sample of 88,550 participants with a median follow-up of 12.5 years provides the resolution needed to estimate a small but consequential slope effect with precision.

**Section 15: Consortia authors and cohort information**

SHARE – study description

The Survey of Health, Ageing and Retirement in Europe is a research infrastructure for studying the effects of health, social, economic and environmental policies over the life-course of European citizens and beyond (https://share-eric.eu/). We used the SHARE easySHARE release (rel9_0_0) for the main analyses, restricting to respondents with baseline age 50–90 years. SHARE contains observations of individuals from 50 years of age from 28 countries, recruited to be representative of the population in each country. Data for the present analyses was extracted from easySHARE (release 8.0.0, February 10th 2022, doi:10.6103/SHARE.easy.800), see (2, 3) for methodological details. The easySHARE release 8.8.0 is based on SHARE Waves 1, 2, 3, 4, 5, 6, 7, and 8 (DOIs:10.6103/SHARE.w1.800, 10.6103/SHARE.w2.800, 10.6103/SHARE.w3.800, 10.6103/SHARE.w4.800, 10.6103/SHARE.w5.800, 10.6103/SHARE.w6.800, 10.6103/SHARE.w7.800, 10.6103/SHARE.w8.800).

HRS – study description

The Health and Retirement Study (HRS) is a longitudinal panel study designed to investigate health, cognitive functioning, retirement, and aging in the United States, based on a nationally representative sample of community-dwelling adults aged 50 years and older and their partners, with replenishment cohorts added over time to maintain population representation. Participants are followed at regular intervals with repeated interviews and assessments, providing harmonized measures of sociodemographic factors, health, and cognition across waves and enabling analyses of late-life trajectories and individual differences. For the present analyses, we used the RAND HRS Longitudinal File (1992–2022 release, Stata format), which provides cleaned and consistently coded variables across waves.

ELSA – study description

“The English Longitudinal Study of Ageing (ELSA) is a panel study of a representative cohort of men and women living in England aged ≥50 years. It was designed as a sister study to the Health and Retirement Study in the USA and is multidisciplinary in orientation, involving the collection of economic, social, psychological, cognitive, health, biological and genetic data. The study commenced in 2002, and the sample has been followed up every 2 years. Data are collected using computer-assisted personal interviews and self-completion questionnaires, with additional nurse visits for the assessment of biomarkers every 4 years. The original sample consisted of 11 391 members ranging in age from 50 to 100 years.” (Steptoe et al., 2013)

TILDA – study description

“The Irish Longitudinal Study on Ageing (TILDA) is a nationally representative longitudinal study of community-dwelling adults residing in Ireland. The study has been described in detail previously. The first wave of data collection took place between 2009 and 2011. The sampling frame was drawn from the Irish Geodirectory, a comprehensive listing of all residential addresses in Ireland. Addresses were randomly sampled by using the Random Sample (RANSAM) method, meaning that each address had an equal probability of selection. Individuals at these addresses aged ≥50 years and capable of providing informed consent were invited to take part in the study. Spouses of any age were also invited.” (Scarlett, 2025)

LASA – study description

“The Longitudinal Aging Study Amsterdam (LASA) is an ongoing prospective cohort study of older adults in the Netherlands, with data on multiple domains of functioning available over a period of more than 30 years of follow-up. The study started in 1992 with a nationally representative sample of older adults aged 55–84 years. Over the years, three refresher cohorts (two cohorts aged 55–64 years in 2002 and in 2012, and one cohort aged 60–86 years in 2024) were added. The main aim of LASA was to describe determinants, trajectories and consequences of (changes in) physical, cognitive, emotional and social functioning” (Hoogendijk et al., 2025).

Data from the Longitudinal Aging Study Amsterdam (LASA) are available for use for specific

research questions, provided that an agreement is made up. Research proposals should be

submitted to the LASA Steering Group, using a standard analysis proposal form that can be

obtained from the LASA website: www.lasa-vu.nl. Files with data published in this publication

are freely available for replication purposes and can be obtained using the same analysis

proposal form. The LASA Steering Group will review all requests for data to ensure that

proposals for the use of LASA data do not violate privacy regulations and are in keeping with

informed consent that is provided by all LASA participants.

**Section 16: Acknowledgement**

LCBC is supported by the European Research Council under grant agreements no. 283634 and no. 725025 (to A.M.F.) and no. 313440 (to K.B.W.), as well as the Norwegian Research Council (325878, 262453 to A.M.F.; 325001, 301395, 239889 to K.B.W.; 249931 to A.M.F & K.B.W.; 324882 to DVP; 325415 to HG), the National Association for Public Health’s dementia research program, Norway (to A.M.F.), and the University of Oslo through the UiO:Life Science convergence environment (to A.M.F). Those who carried out the original analysis and collection of the datasets used in this manuscript bear no responsibility for the further analysis or interpretation of it.

SHARE: funded by the European Commission, DG RTD through FP5 (QLK6-CT-2001-00360), FP6 (SHARE-I3: RII-CT-2006-062193, COMPARE: CIT5-CT-2005-028857, SHARELIFE: CIT4-CT-2006-028812), FP7 (SHARE-PREP: GA N°211909, SHARE-LEAP: GA N°227822, SHARE M4: GA N°261982, DASISH: GA N°283646) and Horizon 2020 (SHARE-DEV3: GA N°676536, SHARE-COHESION: GA N°870628, SERISS: GA N°654221, SSHOC: GA N°823782, SHARE-COVID19: GA N°101015924) and by DG Employment, Social Affairs & Inclusion through VS 2015/0195, VS 2016/0135, VS 2018/0285, VS 2019/0332, VS 2020/0313, SHARE-EUCOV: GA N°101052589 and EUCOVII: GA N°101102412. Additional funding from the German Federal Ministry of Education and Research (01UW1301, 01UW1801, 01UW2202), the Max Planck Society for the Advancement of Science, the U.S. National Institute on Aging (U01_AG09740-13S2, P01_AG005842, P01_AG08291, P30_AG12815, R21_AG025169, Y1-AG-4553-01, IAG_BSR06-11, OGHA_04-064, BSR12-04, R01_AG052527-02, R01_AG056329-02, R01_AG063944, HHSN271201300071C, RAG052527A) and from various national funding sources is gratefully acknowledged (see www.share-eric.eu).

HRS: Sponsored by the U.S. National Institute on Aging (grant U01AG009740) and conducted by the University of Michigan, with additional support from the U.S. Social Security Administration. RAND HRS Longitudinal File (1992–2022 release, Stata format) was produced by the RAND Center for the Study of Aging, with funding from the National Institute on Aging (grant numbers NIA U01AG009740 and NIA R01AG073289) and the Social Security Administration. Santa Monica, CA.

ELSA: Research reported in this publication was supported by the National Institute On Aging of the National Institutes of Health under Award Number R01AG017644. The content is solely the responsibility of the authors and does not necessarily represent the official views of the National Institutes of Health. ELSA is funded by the NIHR Policy Research Programme (HEI) 198_1074_03. The views expressed are those of the author(s) and not necessarily those of the NIHR or the Department of Health and Social Care.

TILDA: The Irish Longitudinal Study on Ageing Wave 5 and Wave 6 were funded by the Department of Health (Government of Ireland) with funding administered by the Health Research Board (TILDA-2017–1) and The Atlantic Philanthropies.

LASA: The Longitudinal Aging Study Amsterdam is supported by a grant from the Netherlands

Ministry of Health Welfare and Sports, Directorate of Long-Term Care. The data collection [in 2012-2013 and 2013-2014] was financially supported by the Netherlands Organization for Scientific Research (NWO) in the framework of the project “New Cohorts of young old in the 21st century” (file number 480-10-014).

ADNI: The ADNI was launched in 2003 as a public-private partnership, led by Principal Investigator Michael W. Weiner, MD. The primary goal of ADNI has been to test whether serial magnetic resonance imaging (MRI), positron emission tomography (PET), other biological markers, and clinical and neuropsychological assessment can be combined to measure the progression of mild cognitive impairment (MCI) and early Alzheimer’s disease (AD). For up-to-date information, see https://adni.loni.usc.edu/. As such, the investigators within the ADNI contributed to the design and implementation of ADNI and/or provided data but did not participate in analysis or writing of this report. A complete listing of ADNI investigators can be found at: http://adni.loni.usc.edu/wp-content/uploads/how_to_apply/ADNI_Acknowledgement_List.pdf. Data collection and sharing for this project were funded by the ADNI (NIH Grant U01 AG024904). ADNI is funded by the National Institute on Aging, the National Institute of Biomedical Imaging and Bioengineering, and through generous contributions from the following: AbbVie, Alzheimer's Association; Alzheimer's Drug Discovery Foundation; Araclon Biotech; BioClinica, Inc.; Biogen; Bristol-Myers Squibb Company; CereSpir, Inc.; Cogstate Eisai Inc.; Elan Pharmaceuticals, Inc.; Eli Lilly and Company; EuroImmun; F. Hoffmann-La Roche Ltd and its affiliated company Genentech, Inc.; Fujirebio; GE Healthcare; IXICO Ltd.; Janssen Alzheimer Immunotherapy Research & Development, LLC.; Johnson & Johnson Pharmaceutical Research & Development LLC.; Lumosity; Lundbeck; Merck & Co., Inc.; Meso Scale Diagnostics, LLC.; NeuroRx Research; Neurotrack Technologies; Novartis Pharmaceuticals Corporation; Pfizer Inc.; Piramal Imaging; Servier; Takeda Pharmaceutical Company; and Transition Therapeutics. The Canadian Institutes of Health Research is providing funds to support ADNI clinical sites in Canada. Private sector contributions are facilitated by the Foundation for the National Institutes of Health (http://www.fnih.org). The grantee organization is the Northern California Institute for Research and Education, and the study is coordinated by the Alzheimer's Therapeutic Research Institute at the University of Southern California. ADNI data are disseminated by the Laboratory for Neuro Imaging at the University of Southern California.

**Section 17: Clinical validation: Langa–Weir cognitive status in HRS**

The primary analyses estimate cohort differences in memory trajectories and calibrate their magnitude against diagnostic benchmarks from ADNI, an external clinical sample. This section provides an internal validation using the same HRS data, linking the person-level Theil–Sen slope estimates directly to a prospectively validated clinical classification of cognitive status, the Langa–Weir scheme, derived from a broader cognitive battery that is independent of the word-list recall measure used to estimate slopes. The Langa–Weir classification was developed and validated specifically for HRS and is derived from the HRS cognitive battery, which includes word-list recall (the outcome used for slope estimation) plus additional measures: orientation

items (date, day, month, year, US president, US vice-president), serial arithmetic (counting backwards from 20 and serial 7s), and an immediate counting-backwards item. The total score (cogtot, 0–27 points) is available from wave 3 onward and has been used extensively to classify cognitive status in HRS-based epidemiological research.

Equivalent validated algorithms exist for ELSA (the TICS-based classification) and SHARE (composite score–based schemes), but their psychometric properties and diagnostic thresholds are not directly comparable to Langa–Weir. Including multiple studies with different classification schemes would introduce cross-study heterogeneity in the outcome definition that would complicate interpretation. We therefore restricted this analysis to HRS, which has the additional advantage of the longest individual follow-up among the five studies (mean 14.7 years, up to 25 years for participants with ≥4 waves), maximizing the age range over which both slope estimates and clinical status can be compared within the same participants. The large HRS sample (n = 22,905 with ≥4 waves; n = 17,870 with Langa–Weir status at last wave, 78.0% match rate) provides adequate power for all analyses. Keeping the analysis within one study

also avoids the need to harmonize outcome definitions across countries and study designs, preserving interpretive clarity.

The Langa–Weir classification assigns each HRS respondent to one of three cognitive categories at each wave based on the self-respondent total cognitive score (cogtot) or, for proxy-interviewed participants, the proxy-rated memory quality (pstmem):

Self-respondents (proxy = 0):

cogtot ≥ 12:	Normal cognition

cogtot 7–11:	Cognitive impairment, no dementia (CIND)

cogtot ≤ 6:	Dementia

Proxy respondents (proxy = 1):

pstmem 1–3 (excellent/very good/good):	Normal/CIND

pstmem 4–5 (fair/poor):	Dementia

The cogtot variable is available as r[W]cogtot for waves 3–13 and as r[W]cogtotp for waves 14–15 in the RAND HRS Longitudinal File (1992–2022 release). The pstmem proxy memory rating is available as r[W]pstmem across all waves. Waves 1–2 were excluded because the HRS cognitive battery changed format at wave 3, consistent with the exclusion of these waves from the primary word-list recall analyses. Negative-coded values (RAND missing codes) were set

to NA before classification.

We extracted the Langa–Weir status at the wave most closely matching each participant's age_max (the endpoint of their slope estimation window), within a 2-year age tolerance. The closest match was selected when multiple waves fell within the tolerance window. This concurrent design, using the classification at approximately the same age as the last slope observation, reflects the hypothesis that the slope estimated over the observation window

predicts the cognitive state at its conclusion. It does not require a prospective follow-up wave beyond the slope window, which would be unavailable for the majority of participants since the slope window typically extends to their last observed HRS assessment.

A potential concern is that Langa–Weir uses word-list recall as one of its components, which is also the basis for the Theil–Sen slope predictor. We address this concern in three ways. First, word-list recall contributes only 10 of the 27 available points to cogtot; the remaining 17 points come from orientation and arithmetic items that are entirely independent of recall. Second, the Theil–Sen slope captures the *rate of change* in recall over multiple waves, whereas the Langa–Weir classification uses the *level* of performance at a single wave, these are conceptually and statistically distinct quantities. Third, the strength of the observed gradient (3% to 35% impairment from flat to severe decline) substantially exceeds what would be expected from a purely tautological relationship between decline rate and single-wave score, which would predict much more modest differentiation among those classified as CIND.

Of the 22,905 HRS participants in the ≥4-wave slope table, 17,870 (78.0%) had a Langa–Weir classification at a wave within 2 years of their age_max. The 22.0% without a matched classification primarily reflect participants whose last slope-window wave did not have a valid cogtot or pstmem assessment (e.g., due to proxy interviews without pstmem data, missing cognitive assessments, or waves falling outside the 1992–2022 file range). Among the 17,870 matched participants, the Langa–Weir distribution at last wave was: normal (15,627; 87.4%), CIND (1,570; 8.8%), and dementia (673; 3.8%). This distribution is consistent with published HRS Langa–Weir prevalence estimates adjusted for the age composition of this ≥4-wave sample, which is healthier and slightly younger on average than the full HRS baseline sample.

Participants were assigned to four slope categories matching the heatmap categories used in the main analyses (Figure 5, middle panel):

Flat:	slope ≥ −0.05 words/year	(n = 4,066; 22.8%)

Mild:	−0.05 to −0.30 words/year	(n = 7,842; 43.9%)

Moderate:	−0.30 to −1.0 words/year	(n = 5,530; 30.9%)

Severe:	slope < −1.0 words/year	(n = 432; 2.4%)

Analysis A: Impairment rate by slope category. For each slope category, the proportion of participants with Langa–Weir impairment (CIND or dementia) was computed with 95% confidence intervals based on the normal approximation to the binomial.

Analysis B: Logistic regression. Two models were fitted:

Model 1: *impaired ~ slope_ts + age_mid_c* (slope in words/year; age centred at 70 years)

Model 2: *impaired ~ slope_ts + age_mid_c + birth_decade* (birth decade centred at 1950)

Odds ratios and 95% confidence intervals were obtained by profile likelihood.

Analysis C: Cohort differences in impairment rate. A binomial GAM with a shared age smooth and cohort intercepts was fitted to the subset with birth cohorts 1920–1960:

*impaired ~ s(age_mid, k=8) + cohort_label* using mgcv::bam() with fREML estimation. Predicted impairment probabilities were computed across an age grid of 55–90 years, holding study at HRS, and masked where cohort × 2-year age bin cell counts fell below 30.

Analysis D: Mediation analysis. The proportion of the cohort effect on impairment mediated by the Theil–Sen slope was estimated using the difference-in-coefficients approach. Two logistic models were fitted:

Unadjusted: *impaired ~ birth_decade + age_mid_c*

Adjusted: *impaired ~ birth_decade + age_mid_c + slope_ts*

The proportion mediated was computed as (b_unadj − b_adj) / b_unadj × 100, where b denotes the log-odds coefficient for birth_decade. A value exceeding 100% (complete mediation) indicates that after removing the slope pathway, there is a slight reversal of the direct cohort effect, which is a known property of logistic-scale mediation when the mediating pathway is dominant and the direct and indirect effects operate in consistent directions (inconsistent

mediation is not present here; both unadjusted and mediator-to-outcome effects are in the expected direction).

*Results summary for the clinical validation in HRS*

Analysis A: Impairment rates followed a monotone gradient across slope categories (3.0%, 7.8%, 24.7%, 34.5% for flat, mild, moderate, and severe decline respectively), representing a 12-fold difference between the shallowest and steepest trajectories.

Analysis B: Each word/year of steeper decline was associated with an 86% reduction in the odds of remaining cognitively normal (OR = 0.143, 95% CI: 0.124–0.165, p < 0.001), after adjusting for age. The cohort effect remained significant after adding slope to the model (OR per decade later birth = 1.17, 95% CI: 1.03–1.34, p = 0.017), indicating that birth cohort has a small independent association with impairment in the adjusted model, which is consistent with the level component of the cohort advantage, which operates independently of slope.

Analysis C: At any given age, later-born cohorts had markedly lower Langa–Weir impairment rates, with the cohort gradient particularly pronounced at older ages where impairment rates diverge steeply.

Analysis D: The cohort effect on impairment was entirely mediated by slope. The unadjusted cohort OR was 0.593 (95% CI: 0.519–0.677, p < 0.001), indicating that each decade of later birth was associated with 41% lower odds of impairment at matched ages. After adjusting for the Theil–Sen slope, the cohort effect was completely eliminated (OR = 1.044, 95% CI: 0.906–1.202,

p = 0.55; proportion mediated = 108%). This complete mediation indicates that the generational protection against clinical cognitive impairment operates entirely through the trajectory mechanism documented in the primary analyses: the cohort improvement in memory slopes fully accounts for the cohort advantage in clinical status.

**Section 18: Identifiable nonlinear APC components**

Overview

The bounding analysis in SI Section 13 addresses the linear components of the age, period, and cohort drifts — specifically, how the identified composites A* and C* can be decomposed under varying assumptions about the linear period drift bP. A complementary approach exploits the fact that nonlinear deviations from the underlying linear trends are identifiable from the data without any assumptions about the linear components, under the assumption of no interactions between age, period, and cohort (Fosse & Winship, 2018; Rohrer, 2025). This section describes the estimation of these identifiable nonlinear components and reports results for both memory level and memory slope.

Estimation procedure

Nonlinear APC components were estimated in two steps. In Step 1, the canonical solution model was fitted:

outcome ~ age_mid + birth_year + study

using weighted least squares (weights identical to those used in the main bounding analysis). This model yields the identified composites A* and C* and produces residuals that are free of all linear APC drifts — both the identified composites and any unidentified redistribution between them. In Step 2, three penalised regression splines were fitted simultaneously to these residuals:

resid ~ s(age_mid, bs="tp", m=c(2,0), k=8) + s(birth_year, bs="tp", m=c(2,0), k=8) + s(I(age_mid + birth_year), bs="tp", m=c(2,0), k=8)

using mgcv::gam() with REML estimation. The m=c(2,0) specification imposes a second-order derivative penalty and removes the null space (intercept and linear component) from each smooth, ensuring that each term captures only pure nonlinear curvature with no linear component. Because the three smooths operate on mutually orthogonal nonlinear components after the linear terms have been removed, they cannot absorb each other's variance in the way that standard period splines can in the full APC model. The three smooth terms correspond to the nonlinear age deviation, the nonlinear cohort deviation, and the nonlinear period deviation respectively. Under the assumption of no APC interactions, these three components are identified from the data without bounding assumptions (Fosse & Winship, 2018; Rohrer, 2025). Smooth estimates were extracted using the gratia package and centred to zero mean for visualization.

Results — memory level

All three nonlinear components were highly significant for memory level (Table S19a). The nonlinear cohort component showed an accelerating positive deviation for cohorts born after approximately 1935, indicating that the linear cohort trend underestimates the gains of the most recently born cohorts — the true generational improvement in memory level is super-linear rather than uniform. For cohorts born before approximately 1935, the nonlinear deviation was negative, meaning those cohorts performed slightly below what the linear trend predicts; this pattern is consistent with the Depression-era disadvantage experienced by cohorts born into the worst economic conditions of the 20th century. The nonlinear period component showed a U-shaped pattern with a trough around 2012–2014, consistent with a period-specific disruption following the 2008 financial crisis, though alternative explanations including cohort compositional changes across waves cannot be excluded. The nonlinear age component was modest, showing mild deviation from linearity that is consistent with slight acceleration of level decline at very old ages beyond what the linear trend captures.

Results — memory slope

All three nonlinear components were also highly significant for memory slope (Table S19b). The nonlinear cohort component showed the complementary pattern to level: a positive deviation (less steep decline than the linear trend predicts) for cohorts born before approximately 1930, declining toward zero for post-1950 cohorts. This indicates that the oldest cohorts show a larger slope advantage beyond the linear trend, while the slope improvement for more recently born cohorts is more uniformly captured by the linear component. Together, the level and slope cohort nonlinearities reveal a dissociation in the temporal structure of the double dividend: level gains are disproportionately concentrated in the most recently born cohorts (accelerating), while slope improvements are disproportionately expressed in the oldest cohorts (front-loaded). The nonlinear period component for slopes showed an inverted-U pattern with a peak around 2012–2014 — the mirror image of the level period pattern — consistent with the hypothesis that the mid-2010s period disruption produced a temporary drop in memory scores (level) while simultaneously producing shallower measured decline rates, possibly through selective attrition of the most cognitively vulnerable participants during that period. The nonlinear age component for slopes showed progressive steepening of decline with advancing age beyond the linear trend, consistent with acceleration of memory loss at very old ages.

**Table S8**. Nonlinear APC smooth terms - memory level

|  | edf | F | p |
| --- | --- | --- | --- |
| s(age_mid) | 5.7039 | 38.4847 | 0 |
| s(birth_year) | 4.9475 | 23.9796 | 0 |
| s(I(age_mid + birth_year)) | 5.7593 | 59.9085 | 0 |

**Table S9**. Nonlinear APC smooth terms - memory slope

|  | edf | F | p |
| --- | --- | --- | --- |
| s(age_mid) | 4.5417 | 10.3920 | 0 |
| s(birth_year) | 5.7109 | 12.4920 | 0 |
| s(I(age_mid + birth_year)) | 5.8106 | 181.2629 | 0 |

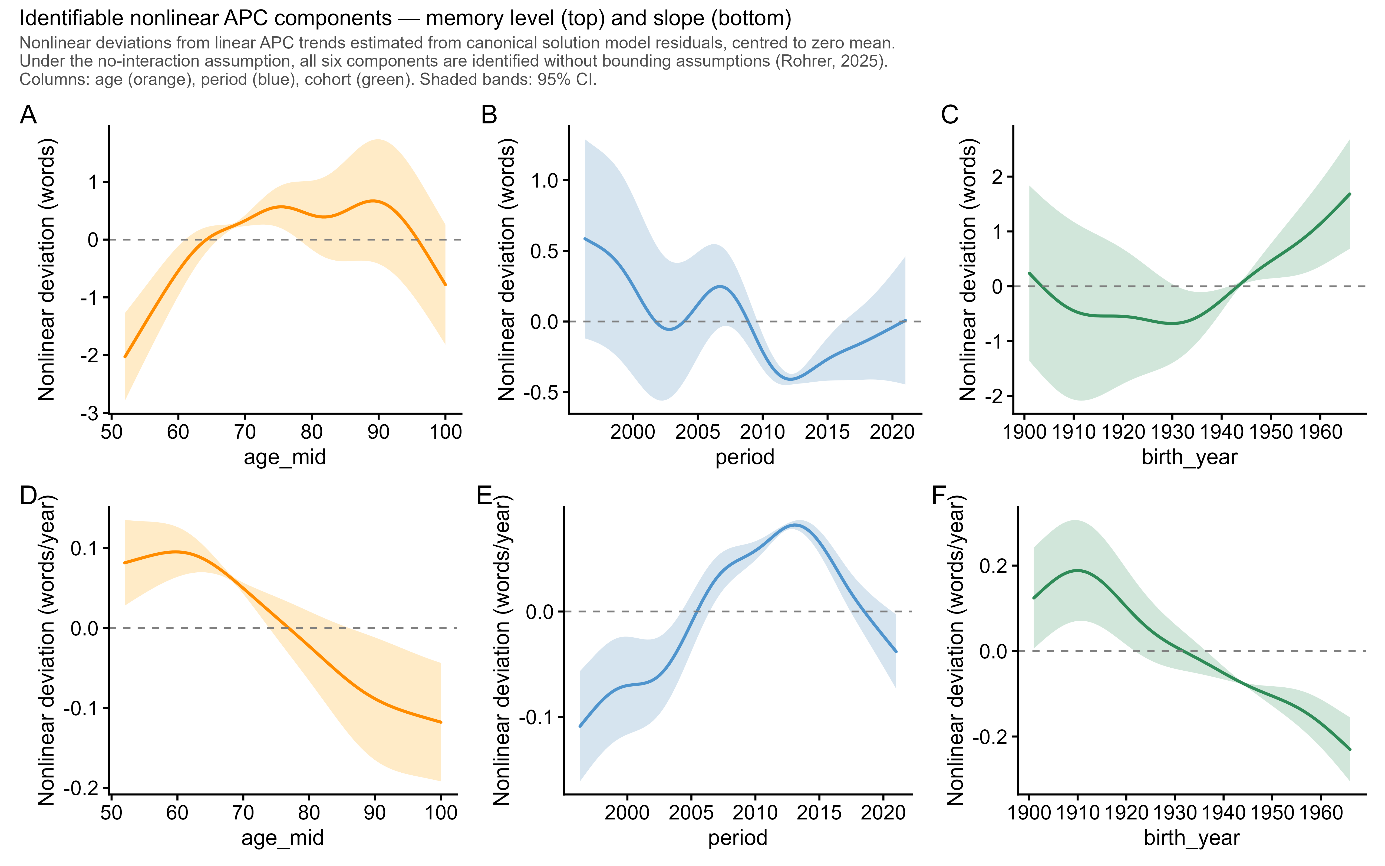

***Figure S18****: Identifiable nonlinear APC components for memory level (top row) and memory slope (bottom row). Columns show the age (orange), period (blue), and cohort (green) nonlinear components respectively. Values are centred to zero mean; positive indicates performance above what the linear trend predicts, negative indicates below. All six components are identified without bounding assumptions under the no-interaction assumption (Rohrer, 2025). Shaded bands: 95% CI.*

**Section 19: Neurobiological validation - hippocampal atrophy by memory slope category**

*Sample*

The neurobiological validation used a subsample of 1,925 participants with both episodic memory assessments and repeated structural MRI, drawn from the longitudinal MRI studies described in a previous paper ([Punctuated memory change: The temporal dynamics and brain basis of memory stability in aging | bioRxiv](https://www.biorxiv.org/content/10.64898/2026.01.20.700557v1)). Inclusion required at least four memory assessments and at least two MRI scans with available hippocampal volume measures. Of 2,749 participants with both memory slopes and hippocampal data, 1,925 had valid bilateral hippocampal atrophy estimates after outlier trimming. The analytic sample had a median MRI follow-up of 4.1 years (range 0.5–18.1 years), median age at median MRI scan of 71.2 years (range 51.5–92.8), and was 53.4% female.

*Memory slope categories*

Memory slopes were estimated using the same Theil–Sen regression framework as the main analyses, applied to the PCA composite memory score used in this subsample - a principal component aggregating multiple memory subtests within each study. Because the PCA composite has different units and variance from the word-list recall measure used in HRS, slope categories were assigned using percentile-based thresholds replicating the approximate category proportions observed in the main HRS sample: Severe (lowest 2%), Moderate (2nd–33rd percentile), Mild (33rd–77th percentile), and Flat (upper 23%). The resulting distribution in the MRI subsample (Flat 22.1%, Mild 43.0%, Moderate 32.9%, Severe 1.9%) closely matched the HRS target proportions, confirming that the percentile bridging preserved the conceptual meaning of the categories across the two memory measures. The Severe group represents the steepest decliners in both samples.

*Hippocampal volume change*

Bilateral hippocampal volume was computed as the sum of left and right hippocampal volumes from FreeSurfer segmentation. For each study, raw volumes were site-adjusted by subtracting the site-specific mean deviation from the global mean, removing systematic scanner and protocol offsets while preserving within-subject longitudinal variation. The Theil–Sen slope of site-adjusted bilateral hippocampal volume over age was computed for each participant, giving an estimate of annual volume change robust to outlying scan pairs. Volume change was expressed as percent per year (%/year) by dividing the slope by the estimated volume at the participant's median scan age. Outliers were removed in two steps: values with |%/year| > 5 were set to missing, followed by removal of values more than 4 SD from the sample mean.

*Statistical model*

The association between memory slope category and hippocampal atrophy rate was estimated by linear regression:

*hippo_pctyr ~ slope_cat + age_c + sex + ICV_z*

where age_c is age at median MRI scan centred at 70 years, sex is a binary factor, and ICV_z is z-scored estimated total intracranial volume (person-level median across available scans). Models were weighted by MRI follow-up span (span / median span across participants), giving longer trajectories proportionally more influence — matching the interval-weighting approach used in the main brain analyses. Reported values are model-adjusted means at reference covariate values (age 70, male, mean ICV). The Flat category served as the reference group. Results are reported as hippocampal volume change (%/year); negative values indicate atrophy.

*Results*

Hippocampal atrophy rates followed a monotone gradient across memory slope categories (Figure SX). Flat decliners showed −0.63%/year (95% CI: −0.73 to −0.53), mild decliners −0.72% (−0.80 to −0.64), moderate decliners −1.04% (−1.13 to −0.95), and severe decliners −2.00% (−2.36 to −1.64). Severe decliners showed approximately three times faster hippocampal atrophy than flat decliners. The confidence intervals for flat and mild categories do not overlap with those for moderate and severe, confirming that the gradient reflects genuine biological differences across the slope categories rather than statistical noise.

**Section 20: Replication under an independent Bayesian hierarchical framework**

The Bayesian hierarchical model was fitted on a combined sample of the three datasets with the most comparable recall measures (SHARE, ELSA, and HRS). In the SHARE data, participants with only one session were omitted due to memory constraints during model fitting. Analyses were restricted to birth cohorts from 1920 to 1965, ages 50 to 100 years, and the period 2000–2025 to ensure sufficient data coverage across all estimates.

Longitudinal recall performance was analyzed using a Bayesian hierarchical regression model designed to separate age-related change from cohort differences while accounting for repeated testing effects and between-dataset variability. Recall scores were modelled as a function of age at assessment and birth year using flexible smooth functions. Age effects were represented using cubic spline terms, and birth year was allowed to influence both baseline performance and the rate of age-related change. This specification allows different cohorts to follow distinct age trajectories rather than assuming parallel age-related decline across generations. The splines used five knots evenly distributed across the age range 50–100 years and the cohort range 1920–1965.

To account for systematic differences between contributing datasets, dataset-specific offsets constrained to sum to zero across samples were included.

Retest effects were modelled using a monotonic spline function of the number of previous test sessions. The first testing occasion was treated as the reference session with no retest effect.

Individual differences in baseline memory level and longitudinal rates of change were captured using subject-specific random intercepts and slopes. These random effects were modelled using skew-normal distributions to allow asymmetric heterogeneity in both baseline performance and ageing trajectories across individuals.

To improve robustness to outliers and non-Gaussian residual variation, observed memory scores were modelled using a Student-t likelihood centred on the latent trajectory for each individual at each observation time point.

The latent memory trajectory at each observation was modelled as the sum of:

–	a smooth cohort-specific ageing trajectory,

–	a monotonic retest effect,

–	a subject-specific intercept,

–	a subject-specific ageing slope,

–	and residual measurement noise.

A schematic of the model structure is shown in Figure S16.

Posterior inference was performed using the No-U-Turn Sampler (NUTS) as implemented in NumPyro.

This model differs from the main analysis in several important respects. First, inference was performed jointly over level, slope, and retest effects, allowing uncertainty in each component to propagate into the others. Second, the slope was defined as the derivative of the level trajectory, ensuring internal consistency of the model. In the main analysis, no interaction between age and period was assumed for the level estimates; however, because a significant slope effect was observed, this assumption does not hold and is therefore relaxed in the present analysis. Third, participants with fewer than four sessions were included, whereas the main analysis required at least four sessions. Because the ability to attend many sessions may reflect selective participation by healthier individuals, including participants with fewer sessions reduces this potential source of bias. Fourth, all sessions for each participant were included in the analysis, and the nonlinear age effect was modelled at each observation rather than reducing individuals to summary statistics at a single reference age. Fifth, whereas the main analysis bins cohorts into 5-year intervals treated as categorical values, the present analysis models the effect of cohort as a continuous smooth function.

*APC Bounding Analysis — Methods*

We performed an age–period–cohort (APC) bounding analysis to estimate upper and lower limits for the age, cohort, and period effects. The posterior estimates of the age and cohort splines can be interpreted as representing their combined effects together with period. Because age, cohort, and period are linearly dependent, a bounding analysis is required to estimate plausible ranges for their separate linear contributions.

We estimated the total linear effects of age and cohort using weighted linear regression applied to the spline estimates while accounting for their uncertainty. We assumed that the linear effect of age on both level and slope should be negative and that the linear effect of cohort should be positive. Using the bounding approach of Fosse and Winship (2019), we estimated ranges for the direct effects of age, cohort, and period.

For the level analysis, we note that the no-interaction assumption between cohort and period is formally violated, because the cohort effect on slope implies an interaction between age and cohort at the level of offset. The analysis is nevertheless included as a first-order approximation. For the slope analysis, the no-interaction assumption holds within the model.

*Results*

Results were consistent with the main analysis. Both the cohort ordering in memory level and the cohort ordering in rate of change were preserved across the full age range. Crucially, the nonlinear cohort pattern was closely replicated: accelerating generational gains in memory level for more recently born cohorts, and a front-loaded slope advantage largest for cohorts born approximately 1930–1955 and diminishing for more recently born cohorts. This qualitative replication across two independent analytical frameworks demonstrates robustness of the core findings. Differences in absolute predicted recall levels between the two analyses are expected given differences in model specification, including the treatment of age trajectories as cohort-specific rather than shared, and sample composition, and are discussed in the main text. The APC bounding analysis was consistent with the main analysis. The results indicate that the period effect on memory slope is likely to be positive, suggesting that the relatively flat slope trajectories observed across cohorts may partly reflect period effects. Apparent linear trends in age and cohort effects reflect the data rather than modelling constraints, as flexible cubic splines without smoothing penalties were used to estimate the trajectories.

**Section 21: MRI subsample – sample characteristics**

MRI samples were identical to a previous paper ([Punctuated memory change: The temporal dynamics and brain basis of memory stability in aging | bioRxiv](https://www.biorxiv.org/content/10.64898/2026.01.20.700557v1)), and we refer to that for details. In brief, we analyzed longitudinal MRI data from 10 independent studies (n =1,978 participants aged > 50 with > 4 episodic-memory test sessions covering 16,342 tests and 7,206 scans). We analyzed longitudinal MRI data from 10 independent studies (n =1,978 participants aged > 50 with > 4 episodic-memory test sessions covering 16,342 tests and 7,206 scans). The datasets include the the Alzheimer’s Disease Neuroimaging Initiative (ADNI) database (<https://adni.loni.usc.edu>) (Mueller, Weiner et al. 2005), AIBL (Ellis, Bush et al. 2009), BLSA (open subset) (Ferrucci 2008), Cognorm (Idland, Sala-Llonch et al. 2020), HABS_HD (Petersen, Zhou et al. 2025), LCBC (Walhovd, Krogsrud et al. 2016), Mayo (Roberts, Geda et al. 2008), PREVENT-AD (Breitner, Poirier et al. 2016, Tremblay-Mercier, Madjar et al. 2021), OASIS3 (LaMontagne, Benzinger et al. 2019), and WRAP (Sager, Hermann et al. 2005) datasets. The initial dataset included individuals with 1 to 14 MRI acquisitions with longitudinal structural MRI data spanning up to 15.8 years. Similarly, memory assessments range from 1 to 30 observations per individual with a follow-up up to 31.6 years.

| Cohort | Sample | n | Obs | Age | Waves | Interval |
| --- | --- | --- | --- | --- | --- | --- |
| ADNI | Full | 821 | 3769 | 76,6 | 4,6 | 3,98 |
| ADNI | >=4 sessions | 447 | 3097 | 77,6 | 6,9 | 6,4 |
| AIBL | Full | 608 | 1260 | 73,7 | 2,1 | 1,74 |
| AIBL | >=4 sessions | 115 | 535 | 73,7 | 4,7 | 5,75 |
| BLSA | Full | 132 | 396 | 82,7 | 3 | 3,27 |
| BLSA | >=4 sessions | 46 | 237 | 83,9 | 5,2 | 5,98 |
| HABS_HD | Full | 2832 | 4576 | 66,4 | 1,6 | 1,44 |
| HABS_HD | >=4 sessions | 131 | 524 | 67,2 | 4 | 6,56 |
| MCSA | Full | 5609 | 20936 | 74,7 | 3,7 | 3,72 |
| MCSA | >=4 sessions | 2742 | 15988 | 75,3 | 5,8 | 6,47 |
| Oasis3 | Full | 923 | 5985 | 74,1 | 6,5 | 7,32 |
| Oasis3 | >=4 sessions | 623 | 5359 | 74,6 | 8,6 | 9,78 |
| COGNORM | Full | 114 | 677 | 75,9 | 5,9 | 5,28 |
| COGNORM | >=4 sessions | 100 | 651 | 75,8 | 6,5 | 5,88 |
| PreventAD | Full | 348 | 1916 | 66,1 | 5,5 | 4,38 |
| PreventAD | >=4 sessions | 265 | 1710 | 66,3 | 6,5 | 5,17 |
| LCBC | Full | 256 | 559 | 65,4 | 2,2 | 4,94 |
| LCBC | >=4 sessions | 53 | 215 | 66 | 4,1 | 12,26 |
| Wrap | Full | 1094 | 5021 | 65,1 | 4,6 | 10,93 |
| Wrap | >=4 sessions | 702 | 4357 | 65,7 | 6,2 | 15,4 |

***Table S10. Sample characteristics for the MRI subsample****. The numbers refer to participant characteristics tied to the memory tests. The essential numbers for the number of MRI scans are provided in the main text. ADNI – Alzheimer’s Disease Neuroimaging Initiative; AIBL – Australian Imaging, Biomarker & Lifestyle Study of Ageing; BLSA – Baltimore Longitudinal Study of Aging; HABS-HD - Health and Aging Brain Studies–Health Disparities; MCSA – Mayo Clinic Study of Aging / Mayo Alzheimer’s Disease Research Center cohorts; OASIS3 – Open Access Series of Imaging Studies (OASIS-3); COGNORM – Oslo University Hospital studies; PREVENT-AD – Pre-symptomatic Evaluation of Experimental or Novel Treatments for Alzheimer Disease; LCBC – Center for Lifespan Changes in Brain and Cognition cohorts; WRAP – Wisconsin Registry for Alzheimer’s Prevention.*

Data was converted to BIDS (Gorgolewski, Auer et al. 2016) and preprocessed using the longitudinal FreeSurfer v.7.1.0 stream (Reuter, Schmansky et al. 2012) for cortical reconstruction and volumetric segmentation of the structural T1w scans (Dale, Fischl et al. 1999, Fischl, Sereno et al. 1999). Data was tabulated based on the *aseg* (subcortical, *N* = 18 regions) atlases (Fischl, Salat et al. 2002).

| Dataset | Memory test | Scanner | Field | Sequence parameters |
| --- | --- | --- | --- | --- |
| ADNI | ADNI-MEM^1^ | Multisite | 1.5/  3.0 | https://adni.loni.usc.edu/methods/documents/mri-protocols/ |
| OUS | CERAD short and long delay recall | Siemens Avanto | 3.0 | MPRAGE. TR: 2400 ms; TE: 3.79 ms, TI: 1000 ms; flip angle 8°, slice thickness: 1.2 mm, FoV 240 x 240, 160 slices. |
|  |  | SiemensPrisma | 3.0 | MPRAGE. TR: 2400 ms; TE: 2.22 ms, TI: 1000 ms; flip angle 8°, slice thickness: 0.8 mm, FoV 240 x 256, 208 slices, iPat = 2. |
| HABS_HD | SEVLT (learning & long delay recall); Logical memory (immediate & delay recall); | SiemensSkyra | 3.0 | MPRAGE. TR: 2300 ms; TE: 2.93 ms, TI: 900 ms; flip angle 9°, slice thickness: 1.2 mm, FoV 240 x 256, 176 slices. |
|  |  | Siemens Vida | 3.0 | MPRAGE. TR: 2300 ms; TE: 2.98 ms, TI: 900 ms; flip angle 9°, slice thickness: 1 mm, FoV 240 x 256, 208 slices. |
| LCBC | CVLT (learning, short & long delay recall) | SiemensAvanto | 1.5 | MPRAGE. TR: 2400 ms; TE: 3.79 ms, TI: 1000 ms; flip angle 8°, slice thickness: 1.2 mm, FoV 240 x 240, 160 slices. |
|  |  | SiemensSkyra | 3.0 | MPRAGE. TR: 2300 ms; TE: 2.98 ms, TI: 850 ms; flip angle 8°, slice thickness: 1 mm, FoV: 256 x 256, 176 slices. |
|  |  | SiemensPrisma | 3.0 | MPRAGE. TR: 2400 ms; TE: 2.22 ms, TI: 1000 ms; flip angle 8°, slice thickness: 0.8 mm, FoV 240 x 256, 208 slices, iPat = 2. |
| OASIS3 | Logical memory immediate | Vision | 1.5 | MPRAGE. TR: 9,7 ms; TE: 4.0 ms, TI: 20 ms; flip angle 10°, slice thickness: 1.25 mm, FoV: 256 x 256, 160 slices. |
|  |  | Siemens Sonata | 1.5 | MPRAGE. TR: 9,7 ms; TE: 3.9 ms, TI: 20 ms; flip angle 15°, slice thickness: 1 mm, FoV: 224 x 256, 160 slices. |
|  |  | Siemens Tim Trio | 3.0 | MPRAGE. TR: 2400 ms; TE: 3.1 ms, TI: 1000 ms; flip angle 8°, slice thickness: 1 mm, FoV 256 x 256, 176 slices. |
|  |  | Siemens Magnetom Vida | 3.0 | MPRAGE. TR: 2300 ms; TE: 2.3 ms, TI: 900 ms; flip angle 9°, slice thickness: 1.2 mm, FoV 240 x 256, 176 slices. |
|  |  | Siemens BioGraph mMR | 3.0 | MPRAGE. TR: 2300 ms; TE: 2.3 ms, TI: 900 ms; flip angle 9°, slice thickness: 1.2 mm, FoV 240 x 256, 176 slices. |
| PreventAD | RBANS list recall and learning, story immediate and delayed recall | Siemens Tim Trio | 3.0 | MPRAGE. TR: 2300 ms; TE: 2.98 ms, TI: 900 ms; flip angle 9°, slice thickness: 1 mm, FoV 240 x 256, 176 slices. |
| Mayo | pzmemory^2^ | GE Discovery MR 750 | 3.0 | SPGR. TR: 7.4 ms; TE: 3.0 ms, TI: 900 ms; flip angle 8°, slice thickness: 1.2 mm, FoV 256 x 256, 166 slices. |
|  |  | GE Signa HDxt | 3.0 | SPGR. TR: 7.0 ms; TE: 2.8 ms, TI: 900 ms; flip angle 8°, slice thickness: 1.2 mm, FoV 256 x 256, 166 slices. |
|  |  | GE Signa HDx | 3.0 | SPGR. TR: 6.9 ms; TE: 3.0 ms, TI: 900 ms; flip angle 8°, slice thickness: 1.2 mm, FoV 256 x 256, 166 slices. |
|  |  | GE Signa Excite | 3.0 | SPGR. TR: 7.2 ms; TE: 3.1 ms, TI: 900 ms; flip angle 8°, slice thickness: 1.2 mm, FoV 256 x 256, 166 slices. |
| WRAP | Logical Memory short & long delay recall; RAVLT learning & long delay recall; BVMT learning & delay recall | GE Signa Premier | 3.0 | SPGR. TR: 8.2 ms; TE: 3.2 ms, TI: 450 ms; flip angle 12°, slice thickness: 1 mm, FoV 256 x 256, 156 slices. |
|  |  | GE Signa Excite | 3.0 | SPGR. TR: 8.4 ms; TE: 1.7 ms, TI: 600 ms; flip angle 10°, slice thickness: 1.2 mm, FoV 256 x 256, 124 slices. |
|  |  | GE Signa HDx | 3.0 | SPGR. TR: 6.6 ms; TE: 2.8 ms, TI: 900 ms; flip angle 8°, slice thickness: 1.2 mm, FoV 256 x 256, 166 slices. |
|  |  | GE Discovery MR 750 | 3.0 | SPGR. TR: 8.2 ms; TE: 3.2 ms, TI: 450 ms; flip angle 12°, slice thickness: 1 mm, FoV 256 x 256, 156 slices. |
| BLSA | CVLT (learning, short & long delay recall) | Philips Achieva | 3.0 | MPRAGE. TR: 6.5 ms; TE: 3.1 ms, TI: 0 ms; flip angle 8°, slice thickness: 1.2 mm, FoV 256 x 256, 170 slices. |
| AIBL | Logical Memory short & long delay recall | Avanto Siemens | 1.5 | MPRAGE. TR: 2300 ms; TE: 2.98 ms, TI: 900 ms; flip angle 9°, slice thickness: 1.25 mm, FoV: 240 x 256, 160 slices. |
|  |  | Verio Siemens | 3.0 | MPRAGE. TR: 2300 ms; TE: 2.98 ms, TI: 900 ms; flip angle 9°, slice thickness: 1.25 mm, FoV: 240 x 256, 160 slices. |
|  |  | Tim Trio Siemens | 3.0 | MPRAGE. TR: 2300 ms; TE: 2.98 ms, TI: 900 ms; flip angle 9°, slice thickness: 1.25 mm, FoV 240 x 256, 160 slices. |

***Table S11.*** *Memory tests and MRI scanner acquisition parameters in each MRI subsample****.*** *Tests related to episodic memory included in the analyses for each sample****.*** *A PC was estimated based on the first time point for which multiple memory measures were available. RAVLT = Rey Auditory Verbal Learning Test; CVLT = California Verbal Learning Test; Logical memory = Memory subtest of the Wechsler Memory Scale. CERAD = Consortium to Establish a Registry for Alzheimer's Disease (CERAD) Word List Memory test. SRT = Buschke Selective Reminding Task. RBANS = Repeatable Battery for Assessment of Neuropsychological Status. Story recall = Story recall and recognition task of episodic memory from Wechsler Neuropsychological Battery. ^1^ADNI-MEM score was computed developed by (Crane, Carle et al. 2012) and consists of a composite score of memory which includes measures from Rey Auditory Verbal Learning Test (learning trials, list, recognition and recalls), Alzheimer Disease Assessment Scale (learning trials, recall, and recognitions), Mini-mental State Examination words, and Logical memory. ^2^pzmemory score was developed by (Knopman et al., 2015) and consists of a composite score of memory tests which includes measures from Logical memory (delayed recall), Weschler’s Visual Reproduction (delayed recall), and RAVLT (delayed recall). For MRI: TR = Repetition Time; TE = Echo Time; TI = inversion time; FoV = Field of View, iPat = in-plane acceleration. ^a^Two matched scanners.*

For datasets not provided in Brain Imaging Data Structure (BIDS) format, data was converted to BIDS(Gorgolewski, Auer et al. 2016). BIDS transformation of ADNI, AIBL, OASIS3, and HABS_HD data was performed with Clinica software (Samper-Gonzalez, Burgos et al. 2018, Routier, Burgos et al. 2021). For sessions with multiple scans, data from the scanners were averaged. Briefly, the images were processed using the cross-sectional stream, which includes the removal of nonbrain tissues, Talairach transformation, intensity correction, tissue and volumetric segmentation, cortical surface reconstruction, and cortical parcellation. Next, an unbiased within-subject template space based on all cross-sectional images was created for each participant, using robust, inverse-consistent registration. The processing of each time point was then reinitialized with common information from the within-subject template to increase reliability and statistical power.

For each sample, we first z-normalized all measures based on the first time point and the different available memory tests. When multiple measures were available, we estimated a main component using Principal Component Analysis (PCA; *prcomp*) with all measures at the first time point as inputs. Missing values were imputed using *imputePCA* from the *missMDA* r-package (Josse and Husson 2016). Only for OASIS3, the imputed number of values was not negligible (> .5%).

*MRI references*

Breitner, J. C. S., J. Poirier, P. E. Etienne and J. M. Leoutsakos (2016). "Rationale and Structure for a New Center for Studies on Prevention of Alzheimer's Disease (StoP-AD)." J Prev Alzheimers Dis **3**(4): 236–242.

Crane, P. K., A. Carle, L. E. Gibbons, P. Insel, R. S. Mackin, A. Gross, R. N. Jones, S. Mukherjee, S. M. Curtis, D. Harvey, M. Weiner, D. Mungas and I. Alzheimer's Disease Neuroimaging (2012). "Development and assessment of a composite score for memory in the Alzheimer's Disease Neuroimaging Initiative (ADNI)." Brain Imaging Behav **6**(4): 502–516.

Dale, A. M., B. Fischl and M. I. Sereno (1999). "Cortical surface-based analysis. I. Segmentation and surface reconstruction." Neuroimage **9**(2): 179–194.

Ellis, K. A., A. I. Bush, D. Darby, D. De Fazio, J. Foster, P. Hudson, N. T. Lautenschlager, N. Lenzo, R. N. Martins, P. Maruff, C. Masters, A. Milner, K. Pike, C. Rowe, G. Savage, C. Szoeke, K. Taddei, V. Villemagne, M. Woodward, D. Ames and A. R. Group (2009). "The Australian Imaging, Biomarkers and Lifestyle (AIBL) study of aging: methodology and baseline characteristics of 1112 individuals recruited for a longitudinal study of Alzheimer's disease." Int Psychogeriatr **21**(4): 672–687.

Ferrucci, L. (2008). "The Baltimore Longitudinal Study of Aging (BLSA): a 50-year-long journey and plans for the future." J Gerontol A Biol Sci Med Sci **63**(12): 1416–1419.

Fischl, B., D. H. Salat, E. Busa, M. Albert, M. Dieterich, C. Haselgrove, A. van der Kouwe, R. Killiany, D. Kennedy, S. Klaveness, A. Montillo, N. Makris, B. Rosen and A. M. Dale (2002). "Whole brain segmentation: automated labeling of neuroanatomical structures in the human brain." Neuron **33**(3): 341–355.

Fischl, B., M. I. Sereno and A. M. Dale (1999). "Cortical surface-based analysis. II: Inflation, flattening, and a surface-based coordinate system." Neuroimage **9**(2): 195–207.

Gorgolewski, K. J., T. Auer, V. D. Calhoun, R. C. Craddock, S. Das, E. P. Duff, G. Flandin, S. S. Ghosh, T. Glatard, Y. O. Halchenko, D. A. Handwerker, M. Hanke, D. Keator, X. Li, Z. Michael, C. Maumet, B. N. Nichols, T. E. Nichols, J. Pellman, J. B. Poline, A. Rokem, G. Schaefer, V. Sochat, W. Triplett, J. A. Turner, G. Varoquaux and R. A. Poldrack (2016). "The brain imaging data structure, a format for organizing and describing outputs of neuroimaging experiments." Sci Data **3**: 160044.

Idland, A. V., R. Sala-Llonch, L. O. Watne, A. Braekhus, O. Hansson, K. Blennow, H. Zetterberg, O. Sorensen, K. B. Walhovd, T. B. Wyller and A. M. Fjell (2020). "Biomarker profiling beyond amyloid and tau: cerebrospinal fluid markers, hippocampal atrophy, and memory change in cognitively unimpaired older adults." Neurobiol Aging **93**: 1–15.

Josse, J. and F. Husson (2016). "missMDA: A Package for Handling Missing Values in Multivariate Data Analysis." Journal of Statistical Software **70**(1): 1–31.

LaMontagne, P. J., T. L. S. Benzinger, J. C. Morris, S. Keefe, R. Hornbeck, C. Xiong, E. Grant, J. Hassenstab, K. Moulder, A. G. Vlassenko, M. E. Raichle, C. Cruchaga and D. Marcus (2019). "OASIS-3: Longitudinal Neuroimaging, Clinical, and Cognitive Dataset for Normal Aging and Alzheimer Disease." medRxiv.

Mueller, S. G., M. W. Weiner, L. J. Thal, R. C. Petersen, C. Jack, W. Jagust, J. Q. Trojanowski, A. W. Toga and L. Beckett (2005). "The Alzheimer's disease neuroimaging initiative." Neuroimaging Clin N Am **15**(4): 869–877, xi–xii.

Petersen, M. E., Z. Zhou, J. R. Hall, N. Phillips, K. L. Meeker, M. T. Borzage, M. N. Braskie, A. L. Clark, Y. Shi, R. A. Rissman, F. Zhang, R. Vintimilla, A. Casas, J. Rhodes, R. C. Barber, L. Johnson, K. Yaffe, A. W. Toga, S. E. O'Bryant and H.-H. S. Team (2025). "Health and Aging Brain Study-Health Disparities (HABS-HD) methods and partner characteristics." Alzheimers Dement (N Y) **11**(3): e70140.

Reuter, M., N. J. Schmansky, H. D. Rosas and B. Fischl (2012). "Within-subject template estimation for unbiased longitudinal image analysis." Neuroimage **61**(4): 1402–1418.

Roberts, R. O., Y. E. Geda, D. S. Knopman, R. H. Cha, V. S. Pankratz, B. F. Boeve, R. J. Ivnik, E. G. Tangalos, R. C. Petersen and W. A. Rocca (2008). "The Mayo Clinic Study of Aging: design and sampling, participation, baseline measures and sample characteristics." Neuroepidemiology **30**(1): 58–69.

Routier, A., N. Burgos, M. Diaz, M. Bacci, S. Bottani, O. El-Rifai, S. Fontanella, P. Gori, J. Guillon, A. Guyot, R. Hassanaly, T. Jacquemont, P. Lu, A. Marcoux, T. Moreau, J. Samper-Gonzalez, M. Teichmann, E. Thibeau-Sutre, G. Vaillant, J. Wen, A. Wild, M. O. Habert, S. Durrleman and O. Colliot (2021). "Clinica: An Open-Source Software Platform for Reproducible Clinical Neuroscience Studies." Front Neuroinform **15**: 689675.

Sager, M. A., B. Hermann and A. La Rue (2005). "Middle-aged children of persons with Alzheimer's disease: APOE genotypes and cognitive function in the Wisconsin Registry for Alzheimer's Prevention." J Geriatr Psychiatry Neurol **18**(4): 245–249.

Samper-Gonzalez, J., N. Burgos, S. Bottani, S. Fontanella, P. Lu, A. Marcoux, A. Routier, J. Guillon, M. Bacci, J. Wen, A. Bertrand, H. Bertin, M. O. Habert, S. Durrleman, T. Evgeniou, O. Colliot, I. Alzheimer's Disease Neuroimaging, B. Australian Imaging and a. Lifestyle flagship study of (2018). "Reproducible evaluation of classification methods in Alzheimer's disease: Framework and application to MRI and PET data." Neuroimage **183**: 504–521.

Tremblay-Mercier, J., C. Madjar, S. Das, A. Pichet Binette, S. O. M. Dyke, P. Etienne, M. E. Lafaille-Magnan, J. Remz, P. Bellec, D. Louis Collins, M. Natasha Rajah, V. Bohbot, J. M. Leoutsakos, Y. Iturria-Medina, J. Kat, R. D. Hoge, S. Gauthier, C. L. Tardif, M. Mallar Chakravarty, J. B. Poline, P. Rosa-Neto, A. C. Evans, S. Villeneuve, J. Poirier, J. C. S. Breitner and P.-A. R. Group (2021). "Open science datasets from PREVENT-AD, a longitudinal cohort of pre-symptomatic Alzheimer's disease." Neuroimage Clin **31**: 102733.

Walhovd, K. B., S. K. Krogsrud, I. K. Amlien, H. Bartsch, A. Bjornerud, P. Due-Tonnessen, H. Grydeland, D. J. Hagler, Jr., A. K. Haberg, W. S. Kremen, L. Ferschmann, L. Nyberg, M. S. Panizzon, D. A. Rohani, J. Skranes, A. B. Storsve, A. E. Solsnes, C. K. Tamnes, W. K. Thompson, C. Reuter, A. M. Dale and A. M. Fjell (2016). "Neurodevelopmental origins of lifespan changes in brain and cognition." Proc Natl Acad Sci U S A **113**(33): 9357–9362.

**Section 22: Sub-sample funding - MRI**

ADNI: The ADNI was launched in 2003 as a public-private partnership, led by Principal Investigator Michael W. Weiner, MD. The primary goal of ADNI has been to test whether serial magnetic resonance imaging (MRI), positron emission tomography (PET), other biological markers, and clinical and neuropsychological assessment can be combined to measure the progression of mild cognitive impairment (MCI) and early Alzheimer’s disease (AD). For up-to-date information, see https://adni.loni.usc.edu/. As such, the investigators within the ADNI contributed to the design and implementation of ADNI and/or provided data but did not participate in analysis or writing of this report. A complete listing of ADNI investigators can be found at: http://adni.loni.usc.edu/wp-content/uploads/how_to_apply/ADNI_Acknowledgement_List.pdf. Data collection and sharing for this project were funded by the ADNI (NIH Grant U01 AG024904). ADNI is funded by the National Institute on Aging, the National Institute of Biomedical Imaging and Bioengineering, and through generous contributions from the following: AbbVie, Alzheimer's Association; Alzheimer's Drug Discovery Foundation; Araclon Biotech; BioClinica, Inc.; Biogen; Bristol-Myers Squibb Company; CereSpir, Inc.; Cogstate Eisai Inc.; Elan Pharmaceuticals, Inc.; Eli Lilly and Company; EuroImmun; F. Hoffmann-La Roche Ltd and its affiliated company Genentech, Inc.; Fujirebio; GE Healthcare; IXICO Ltd.; Janssen Alzheimer Immunotherapy Research & Development, LLC.; Johnson & Johnson Pharmaceutical Research & Development LLC.; Lumosity; Lundbeck; Merck & Co., Inc.; Meso Scale Diagnostics, LLC.; NeuroRx Research; Neurotrack Technologies; Novartis Pharmaceuticals Corporation; Pfizer Inc.; Piramal Imaging; Servier; Takeda Pharmaceutical Company; and Transition Therapeutics. The Canadian Institutes of Health Research is providing funds to support ADNI clinical sites in Canada. Private sector contributions are facilitated by the Foundation for the National Institutes of Health (http://www.fnih.org). The grantee organization is the Northern California Institute for Research and Education, and the study is coordinated by the Alzheimer's Therapeutic Research Institute at the University of Southern California. ADNI data are disseminated by the Laboratory for Neuro Imaging at the University of Southern California.

OUS-COGNORM is funded by the South-Eastern Norway Regional Health Authorities (#2017095) The Norwegian Health Association (#19536) and by Wellcome Leap’s Dynamic Resilience Program (jointly funded by Temasek Trust) #104617).

AIBL: Data used in the preparation of this article was obtained from the Australian Imaging Biomarkers and Lifestyle flagship study of ageing (AIBL) funded by the Commonwealth Scientific and Industrial Research Organisation (CSIRO) which was made available at the ADNI database (www.loni.usc.edu/ADNI). The AIBL researchers contributed data but did not participate in analysis or writing of this report. AIBL researchers are listed at www.aibl.csiro.au.

BLSA: This study uses research data from The Baltimore Longitudinal Study of Aging study (BLSA) that has been made available through the LONI IDA and GAAIN. The BLSA is supported by the Intramural Research Program (IRP) of the National Institute on Aging (NIA). The authors thank the investigators, staff, and participants of the BLSA study and the neuroimaging sub-study for making the data available. BLSA investigators have not contributed to nor approved, and are not in any way responsible for, the contents of this article.

HABS_HD: Research reported on this publication was supported by the National Institute on Aging of the National Institutes of Health under Award Numbers R01AG054073, R01AG058533, R01AG070862, P41EB015922 and U19AG078109. The content is solely the responsibility of the authors and does not necessarily represent the official views of the National Institutes of Health.

PREVENT-AD: PREVENT-AD was funded by the Canadian Institutes of Health Research, McGill University, the Fonds de Recherche du Québec – Santé, Alzheimer’s Association, Brain Canada, the Government of Canada, the Canada Fund for Innovation, the Douglas Hospital Research Centre and Foundation, the Levesque Foundation, an unrestricted research grant from Pfizer Canada. Private sector contributions are facilitated by the Development Office of the McGill University Faculty of Medicine and by the Douglas Hospital Research Centre Foundation (http://www.douglas.qc.ca/).

OASIS3: Longitudinal Multimodal Neuroimaging: Principal Investigators: T. Benzinger, D. Marcus, J. Morris; NIH P30 AG066444, P50 AG00561, P30 NS09857781, P01 AG026276, P01 AG003991, R01 AG043434, UL1 TR000448, R01 EB009352. AV-45 doses were provided by Avid Radiopharmaceuticals, a wholly owned subsidiary of Eli Lilly.

MCSA: Data used in preparation of this article were shared by the Mayo Clinic Study of Aging (MCSA). The MCSA is funded by the following sources: NIH U01 AG006786, R01 AG034676, R37 AG011378, R01 AG041851, R01 NS097495, R01 AG056366, R01 AG068206, P30 AG062677, GHR Foundation, Elsie and Marvin Dekelboum Family Foundation, Liston Award, Schuler Foundation, Alexander Foundation, Mayo Foundation for Medical Education and Research.

WRAP: Data used in preparation of this articles was made kindly available by the University of Wisconsin. The Wisconsin Registry for Alzheimer's Prevention (WRAP) was funded by the following NIH sources: R01AG027161, and R01AG021155, R01AG062285, and R01AG062167 for MRI data collection.

VETSA: Parts of the data are from VETSA, which is funded by National Institute of Aging (NIA) R01 grants AG018384, AG018386, AG050595, AG022381 and AG076838. The content is the responsibility of the authors and does not necessarily represent the official views of the NIA, the NIH, the US Department of Veterans Affairs, the US Department of Defense, the National Personnel Records Center, the National Archives and Records Administration, the Internal Revenue Service, the National Opinion Research Center, the National Research Council, the National Academy of Sciences or the Institute for Survey Research. Temple University provided invaluable assistance in the conduct of the Vietnam Era Twin Registry. The Cooperative Studies Program of the US Department of Veterans Affairs provided financial support for development and maintenance of the Vietnam Era Twin Registry. We would also like to acknowledge the continued cooperation and participation of the members of the Vietnam Era Twin Registry and their families.
